# What matters to families about the healthcare of preterm or low birth weight infants: A qualitative evidence synthesis

**DOI:** 10.1101/2022.06.22.22276770

**Authors:** Lisa Hurt, David Odd, Mala Mann, Hannah Beetham, Emma Dorgeat, Thomas CW Isaac, Annie Ashman, Fiona Wood

## Abstract

**Background:** Preterm and low birth weight (LBW) infants have complex healthcare needs. In this qualitative evidence synthesis, we aimed to understand what matters to families about the healthcare provided from birth to preterm or LBW infants, to inform appropriate planning and delivery of care.

**Methods:** We searched nine databases and the reference lists of included studies to identify eligible studies that used qualitative methods to examine the views of families relating to healthcare for preterm or LBW infants. Studies in all countries (low, middle, and high-income) and healthcare settings (home, community, primary, secondary and tertiary care) were eligible for inclusion. Study quality was assessed using the Critical Appraisal Skills Programme checklist for qualitative studies. The GRADE-CERQual approach was used to assess confidence in each review finding. Thematic synthesis techniques were used for analysis.

**Results:** 203 studies (in 208 papers) were eligible for inclusion. 49 studies (in 51 papers) from 25 countries were sampled for the analysis, based on pre-specified criteria (methodological quality; data richness; and ensuring representation across continents and from settings with varying resources). Eight analytical themes were identified. What mattered to carers was: a positive outcome for the child; active involvement in care; support to cope at home after discharge; emotional support for the family; the healthcare environment; information needs were met; logistical support was available; and positive relationships with staff. Confidence in most results was high to moderate.

**Conclusion:** We found high consistency in what matters to families regardless of location or resources, even though carers reported a variety of experiences in the care of their preterm or LBW infant. Families views on care in neonatal units have been extensively studied. Further research is needed on what matters to fathers and other family members, and to families receiving community-based care especially in low and middle-income countries.

**What is already known on this topic:**

- Preterm and low birthweight births are a major cause of poor health globally.
- Despite advances in neonatal care, preterm and low birthweight infants often require complex and prolonged medical intervention, with survivors at increased risk of lifelong disability and poor quality of life.

**What this study adds:**

- Carers have strong views about what matters in relation to the healthcare provided to preterm and low birthweight infants, and these are remarkably consistent across different countries and healthcare settings.
- What matters to carers is a positive outcome for the child; active involvement in care; support to cope at home after discharge; emotional support for the family; the healthcare environment; information needs are met; logistical support is available; and positive relationships with staff.

**How this study might affect research, practice or policy:**

- It is vital that international and local organisations working to ensure high-quality and equitable care for small and sick newborns worldwide understand what matters to families.
- Further research is needed on the views of fathers and other family members, and on families receiving community-based care especially in low and middle-income countries.

## INTRODUCTION

Infants born preterm and/or with low birth weight (less than 2500g (LBW)) are at high risk of mortality and morbidity.[1–3] Their survival has been substantially improved by recent advances in neonatal care.[4 5] However, they often require complex and prolonged medical intervention, with survivors at increased risk of lifelong disability and poor quality of life. [6 7] While risks can be reduced through interventions provided before or during pregnancy and after birth,[8] these births represent a major cause of poor health globally,[1] with the highest burden in low and middle-income countries.[9–11]

As a result, there has recently been renewed focus on ensuring high-quality and equitable care for small and sick neonates worldwide.[11] Family-centred care, where family members work with healthcare professionals (HCPs) to play an active role in providing emotional, social, and developmental support to the vulnerable infant, has demonstrable benefits[12] but is not available in all settings. The impact of a preterm or LBW birth on families is also increasingly understood, with consequences for attachment and bonding and parental health and wellbeing immediately after birth and beyond.[13 14]

An understanding of families’ views and values is needed, to ensure that healthcare meets the needs of infants and carers and to identify improvements required to existing care.[15 16] The aim of this qualitative evidence synthesis (QES) was to systematically review literature that aimed to understand the perspectives of families about the care provided by health services for preterm or LBW infants in hospitals and in the community.

## METHODS

The protocol for this QES was registered with PROSPERO on 6 July 2021 (CRD42021261934). The review is reported in accordance with PRISMA guidelines (see Supplementary Table 1 for PRISMA checklist) and the Enhancing Transparency in Reporting the synthesis of Qualitative research (ENTREQ) statement.[17]

### Eligibility criteria: Topic of interest

We used the PEO (Population, Exposure, Outcomes) framework to set out the inclusion criteria for the QES (Table 1).

**Table 1.** Review Population, Exposure, Outcomes

|  |  |
| --- | --- |
| **Population and their problems** | Included populations were:<br>- Mothers, fathers, parents, families, carers, or other family members with first-hand experience of healthcare for a preterm or low birth weight infant;<br>- In all study settings (low, middle and high income);<br>- In all healthcare settings (home, community, primary, secondary and tertiary care). |
| **Exposure** | Healthcare delivered from birth to 24 months of age for:<br>- Infants born preterm (<37 weeks or sub-categories);<br>- Infants born with low birth weight (<2500g or sub-categories). |
| **Outcomes or themes** | Qualitative or mixed methods studies examining views or values, including:<br>- What matters to, or is important to, or is valued by families;<br>- What they find acceptable and not acceptable in the healthcare of their infant. |

We aimed to synthesise the views of family members including mothers, fathers, parents, carers, grandparents, siblings, or other family members. The views of healthcare workers (paid or unpaid) were outside the remit of the review. Studies in all countries (low, middle, and high-income) and healthcare settings (home, community, primary, secondary and tertiary care) were eligible for inclusion. A preterm infant was defined as an infant born alive before 37 completed weeks of gestation,[18] with further sub-divisions possible into extremely preterm (less than 28 weeks), very preterm (28 to 31 weeks), and moderate to late preterm (32 to 36 weeks). Low birth weight was defined as weight at birth of less than 2500 g.[19] This could be further categorized into very low birth weight (<1500 g), and extremely low birth weight (<1000 g).

We excluded studies that only described families’ experiences (for example, how long the infant was in hospital, where the parents stayed during the admission, and so on). We did not exclude studies that explored parents’ views in the context of asking about a specific intervention (for example, kangaroo mother care or continuous positive airway pressure). However, the information we extracted from those studies related to how the intervention contributed to parents’ views of healthcare in general, rather than their views about the specific intervention.

In our protocol, we specified that data collected from birth until the infant was 24 months of age would be included. However, as we screened studies, we found several high-quality studies that had collected information when the children were older. We therefore changed our criteria to include studies collecting data later in childhood, on the condition that the data collection was clearly referring to the care received before the infant was 24 months of age. We used uncorrected chronological age at the time of the data collection where possible, but studies that used corrected age were eligible for inclusion if this was the only measure reported.

### Eligibility criteria: Types of studies

Primary studies that used qualitative study designs such as ethnography, phenomenology, case studies, grounded theory studies and qualitative process evaluations were eligible for inclusion. We included studies that used both qualitative methods for data collection (for example, focus group discussions, individual interviews, observation, diaries, document analysis, open-ended survey questions) and qualitative methods for data analysis (for example, thematic analysis, framework analysis, grounded theory). We excluded studies that collected data using qualitative methods but did not analyse these data using qualitative analysis methods (for example, open-ended survey questions where the response data are analysed using descriptive statistics only). Mixed methods studies were eligible if it was possible to extract the data that were collected and analysed using qualitative methods.

To ensure that the data reflected the views of a contemporary cohort of parents, studies published in or after 2000 were eligible. We excluded studies that had only been published as conference abstracts and unpublished PhD or Masters theses. Studies had to be published in English (the language spoken by the review team) to ensure that themes were appropriately identified, understood, and represented.

### Reflexive note

We are a multi-disciplinary team including a neonatologist, epidemiologist with expertise in child health, qualitative methodologist, information specialist, medical students, and specialty registrars in Public Health and Paediatrics. None of the team had personal experience of being a parent of a preterm or low birthweight infant. We maintained a reflexive stance throughout the different review stages. Our varied backgrounds and seniority, and different levels of topic expertise helped us to remain mindful of our presuppositions, as we encouraged each other to consciously acknowledge our own professional assumptions and how these skewed our interpretation of our findings. The senior author kept a reflexive journal throughout the review process to document and reflect on progress and decisions made.

### Information sources

We searched African Journals Online, ASSIA, CINAHL, Cochrane Central Register of Controlled Trials, LILACS, MEDLINE, PsycINFO, Sociological Abstracts, and Web of Science for primary studies published between 1 January 2000 and 14 June 2021. We also reviewed the reference lists of all included studies and relevant systematic reviews.

### Search strategy

Key words and Medical Subject Headings (MeSH) terms for the three main topic areas (qualitative research and values; preterm and low birth weight; and healthcare) were identified and combined using OR. The three groups were combined using “AND”, and limited to papers published in or after 2000. A full list of search terms used and the results of a scoping search conducted in MEDLINE are presented in Web Appendix 1. The MEDLINE search strategy was adapted for the other databases, with searches in some (such as African Journals Online and LILACS) limited to key words as these databases do not include MeSH terms. No language restrictions were placed on the searches. Instead, we identified English language papers during the screening process.

### Study screening

Searches from individual databases were downloaded into Endnote 20 and duplicates removed. Titles and abstracts were screened independently by two reviewers against our established eligibility criteria. Full-text versions were obtained for the papers potentially meeting the inclusion criteria and were screened independently by two reviewers. Disagreements were resolved through discussion and in consultation with others in the review group.

### Data extraction

The data extraction form collected details on the methods of included studies (including the population studied, healthcare setting, inclusion and exclusion criteria, recruitment method, and data collection and analysis methods); the results (including the themes identified, and any relevant quantitative data); and the quality assessment items from the Critical Appraisal Skills Programme (CASP) checklist (see Web Appendix 2). The form was piloted using two different studies before data extraction began. Data from each paper were extracted by two reviewers. Multiple publications from the same study were linked and compared for completeness and contradictions.

### Quality assessment

We used the CASP checklist for qualitative studies to assess the quality of all eligible studies.[20] Each paper was classified as high, moderate, or low quality, based on a qualitative assessment of all elements of the checklist. Two reviewers completed the assessment independently for each paper. A third reviewer evaluated all discrepancies, and these were resolved in consultation with the review group.

### Study sampling

Qualitative evidence synthesis aims to identify variation in concepts rather than an exhaustive sample of papers. When large numbers of eligible studies are identified, the Cochrane Effective Practice and Organisation of Care (EPOC) Qualitative Evidence Syntheses guidance recommends sampling studies because too much data can impair analysis quality.[21] We limited our sample to 49 studies (51 papers) from the total eligible as this allowed for data saturation. The sampling criteria were pre-specified, to: include studies from different countries with varying resources; prioritise the inclusion of studies that scored three or higher on a data richness scale (see Web Appendix 3)[21]; and select studies identified as good or average quality according to the CASP classification.

### Synthesis methods

Thematic synthesis was used,[22] drawing on concepts used for thematic analysis in qualitative research in primary studies to identify and develop themes across the included studies. Included papers were imported into NVivo12,[23] and information from all sections was used in the data coding. Data were coded inductively using a line-by-line method according to meaning and content (descriptive coding). Codes could be structured in tree form or in free form without hierarchical structure. New codes were created as necessary as we progressed through the papers and similar, or related, codes were grouped. Finally, coded data were synthesised into analytical themes which brought together groupings of the descriptive themes. The initial analysis was conducted by the senior author, and revised in discussion with the author group.

### Certainty assessment

We used the GRADE-CERQual (Confidence in the Evidence from Reviews of Qualitative research) approach to assess our confidence in each finding.[24] The GRADE-CERQual assessment is based on four components: 1) methodological limitations of included studies; 2) coherence of the review finding; 3) adequacy of the data contributing to a review finding; and 4) relevance of the included studies to the review finding.[25–28] The author group discussed and made a judgement (based on a consensus view) about the overall confidence in the evidence supporting each finding (high, moderate, low, or very low). All findings started as high confidence and were downgraded if there were concerns regarding any of the GRADE-CERQual components.

## RESULTS

### Included studies

7302 studies were identified by database screening once duplicates were removed (see Figure 1). 6550 studies were excluded by title and abstract screening. 750 full text papers were assessed for eligibility (2 reports could not be retrieved). Of these, 203 studies (reported in 208 papers) were eligible for inclusion. We selected 49 studies, reported in 51 papers, from 25 countries for inclusion in the analysis using the pre-specified criteria.[29–79] The studies that were eligible but not included are listed in Web Appendices 4 and 5.

**Figure 1.**
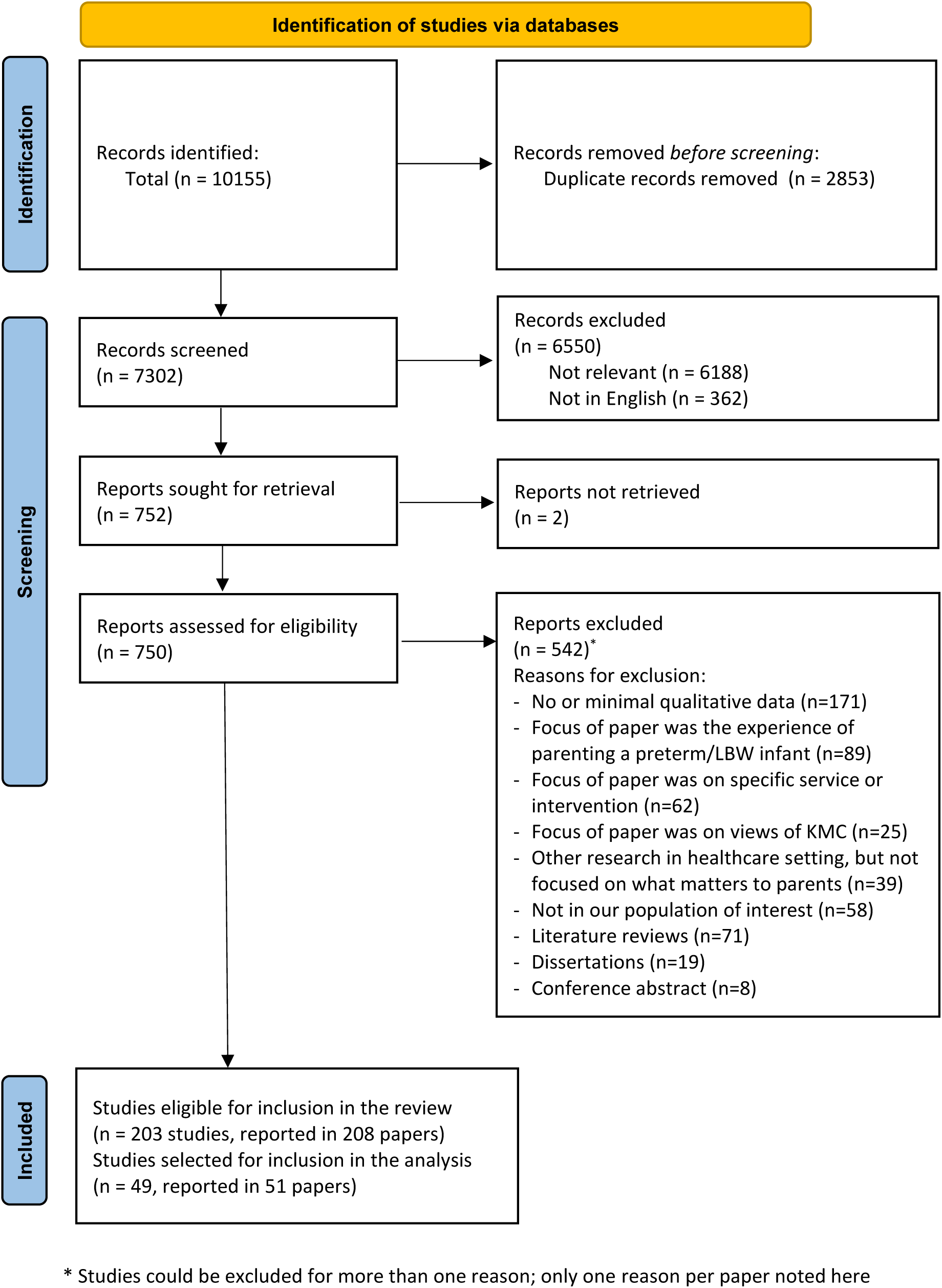
Prisma flow diagram

### Study and participant characteristics

Table 2 (next page) shows the characteristics of the included studies. 45 of the 51 papers were published in 2012 or later. Most (34 of 49 studies) were conducted in high-income settings, with nine studies from countries classified by the World Bank as upper-middle income, four in lower-middle income countries, one in a low-income country (Malawi), and one from Taiwan (which is not included in the World Bank list). Interviews were used for data collection in 38 of 49 studies. Three studies used focus groups, four used a combination of participant observation and interviews, one analysed free-text responses from a questionnaire, two used data collected via a voice-recording App, and one asked participants to keep a daily journal.

**Table 2.** Characteristics of the sampled studies (n=49)

| Author / Date | Aim of study (as reported in the papers) | Country (income)* | Data collection dates | Methods | Healthcare setting of interest | Recruitment method | Data richness score | CASP rating |
| --- | --- | --- | --- | --- | --- | --- | --- | --- |
| Abeasi 2020 | To explore challenges of mothers with preterm infants during hospitalisation in a tertiary institution in Nigeria | Nigeria (lower-middle) | 2019 | Qualitative interviews (location not reported)<br>Content analysis | Neonatal unit | Purposive sampling from SCBU at a teaching hospital | 4 | Good |
| Adama 2017 | To explore Ghanaian fathers' experiences of caring for preterm infants in the neonatal unit after discharge | Ghana (lower-middle) | Feb-June 2015 | Qualitative interviews (conducted in family home)<br>Thematic analysis, using a narrative space framework | Neonatal unit & community | Recruited from four government hospitals as part of a larger study; no further details reported | 4 | Average |
| Amorim 2019 | To explore needs of parents of very preterm infants hospitalised in neonatal intensive care units according to their socioeconomic position, obstetric history and infant's characteristics | Portugal (high) | Nov 2013-April 2014 | Mixed methods (qualitative element = interviews; conducted in the family home (n=19), at a university (n=6) or in hospital (n=1))<br>Thematic content analysis | Neonatal unit | Recruited from all seven public level III NICU in the Northern Health Region<br>Purposive sampling from those who had completed quantitative questionnaires | 4 | Good |
| Arnold 2013 | To assess parents' first experiences of their very preterm infants and the neonatal intensive care unit | UK (high) | Not reported | Qualitative interviews (conducted in hospital or at the family home)<br>Thematic analysis | Neonatal unit | Recruited from three NICU in tertiary care hospitals in South East England<br>All eligible parents invited by letter and sent reminder letter by research nurse | 3 | Good |
| Aydon 2018 | To explore the experiences of parents with infants born between 28-32 weeks' gestation during transition through the neonatal intensive care unit and discharge | Australia (high) | Oct 2014-Feb 2015 | Qualitative interviews (pre- and post-discharge; location not reported) and an online survey<br>Thematic analysis | Neonatal unit & transition post-discharge | Recruited at a tertiary maternity hospital<br>Parents invited by neonatal nurse researchers not directly involved in infant care | 4 | Good |
| Blomqvist 2012 | To describe fathers' experiences of providing kangaroo mother care to their preterm infants | Sweden (high) | 2009 | Qualitative interviews (conducted at family home)<br>Thematic content analysis | Neonatal unit | Recruited from NICUs in two hospitals<br>Questionnaire given to all fathers on NICU with option to be invited for interview; all who agreed approached by mail and phone call | 4 | Good |
| Brødsgaard 2015 | An evaluation of an Early Discharge Programme model for preterm infants based on family-centred care, to describe its impact on the infants and families. | Denmark (high) | Not reported | Mixed methods (qualitative element = focus groups, conducted in hospital after discharge)<br>Deductive theory-driven and directed content analysis | Neonatal unit & community | Purposive sampling of parents who had been enrolled in an Early Discharge Programme for preterm infants; parents contacted by telephone after discharge | 4 | Good |
| Chang Lee 2009 | To explore Taiwanese mothers' parenting experiences when their preterm infants were in NICUs | Taiwan (not in World Bank list) | Not reported | Qualitative interviews and participant observation (both in NICU)<br>Grounded theory analysis | Neonatal unit | Mothers recruited from major neonatal care centre; no further details reported | 4 | Average |
| Dorner 2020 | To determine where, and how, neonatal intensive care unit parents want to receive early neurodevelopmental screening information about their child's future risk of cerebral palsy and other disabilities | USA (high) | March 2018-June 2018 | Qualitative interviews (location not reported)<br>Thematic content analysis | Neonatal unit | Study conducted at a level IV NICU and an associated level III NICU; parents approached at bedside by researcher to explain the study and obtain consent | 4 | Good |
| dos Santos 2014 | To understand the meaning of home visits by neonatal nurses for mothers of premature infants | Brazil (upper-middle) | Not reported | Qualitative interviews (conducted at follow-up clinic)<br>Thematic content analysis | Community (with a focus on home visits) | Participants were part of another project; mothers were approached by female graduate students when they attended a follow-up clinic | 4 | Good |
| Feeley 2013 | To explore what fathers perceive to be facilitators or barriers to their involvement with their infants | Canada (high) | Not reported | Qualitative interviews (conducted in hospital whilst infant was admitted)<br>Thematic content analysis | Neonatal unit | Participants from two NICUs in a major Canadian urban centre<br>Approached by clinical staff members to obtain permission for researcher to contact them | 3 | Good |
| Fernandez Medina 2021 | To explore and describe the experiences of parents of technology-dependent extremely preterm infants of socio-family support after hospital discharge | Spain (high) | Oct 2019-Dec 2019 | Qualitative interviews (conducted by phone after discharge)<br>Analysis using philosophical hermeneutics | Community & outpatient care | Purposeful sampling from four Spanish organisations that support families with preterm infants; organisations received a letter and identified participants, who then contacted main author | 3 | Good |
| Finlayson 2014 | To explore mothers' perceptions of family centred care in neonatal intensive care units in England | UK (high) | Not reported | Qualitative interviews (conducted in hospital whilst infant was admitted)<br>Thematic networks analysis | Neonatal unit | Convenience sample identified by the admissions officer at three NICUs in north west England | 5 | Good |
| Franck 2017 | To discover parents' views, experiences, concerns, and recommendations about the care provided to them and their babies throughout the perinatal and neonatal healthcare journey | UK (high) | Not reported | Focus groups (conducted in "local setting")<br>Thematic content analysis | Neonatal unit & post-discharge care | Parents of infants who received care in 1 of the 7 NICUs in Northern Ireland were invited to participate via a notice posted on the TinyLife Facebook page. | 4 | Good |
| Gallegos-Martinez 2013 | To identify and analyse the significance of participation for patients in a Neonatal Unit of a maternity hospital in San Luis Potosí | Mexico (upper-middle) | Not reported | Qualitative interviews (location not reported)<br>Thematic content analysis | Neonatal unit | Recruited from level II neonatal unit in a public maternity hospital; no further details provided | 4 | Average |
| Glazer 2021 | To understand how a racially and ethnically diverse sample of mothers experienced high-risk obstetric and neonatal care, and whether or not there were differences in these experiences by race and ethnicity that may suggest reasons for variation in quality of care and outcomes | USA (high) | Not reported | Focus groups (location not reported)<br>Thematic analysis | Neonatal unit | Purposive, convenience sample from deliveries in a New York medical centre; accessed through medical records then contacted by telephone and via flyers in hospital-affiliated clinics | 4 | Good |
| Granrud 2014 | To describe how the parents of premature infants experience the transportation of their infant from the NICU at a university hospital to a unit at a local hospital | Norway (high) | April -June 2011 | Qualitative interviews (conducted in hospital or at family home)<br>Inductive content analysis | Neonatal unit & transfers between hospitals | Consecutive selection from two NICUs at two hospitals and retrospective recruitment of additional participants | 4 | Good |
| Guillaume 2013 | To explore, through parents' accounts, how an early bond with their very premature child is established and to identify their experiences of caregivers, and the concrete things that helped and hindered them | France (high) | Nov 2009-March 2012 | Qualitative interviews (conducted in hospital whilst infant was admitted)<br>Discourse analysis | Neonatal unit | Recruited from three tertiary care centres in Paris; identified and approached by nurses participating in the research | 4 | Good |
| Hägi-Petersen 2021 | To gain in-depth knowledge of mothers' and fathers' experiences of the whole trajectory of an early in- | Denmark (high) | Sept 2018-Jan 2020 | Qualitative interviews (conducted at the family home) | Neonatal unit & community | Convenience sample of parents recruited from two neonatal wards that offered | 4 | Good |
|  | home care programme supported by video consultations with a neonatal nurse |  |  | Inductive content analysis | (focus was in-home care) | early in-home care programmes; approached by neonatal nurse on ward |  |  |
| Hendriks 2017 | To explore parental attitudes and values in the end-of-life decision-making process of extremely preterm infants (gestational age < 28 weeks) | Switzerland (high) | Not reported | Qualitative interviews (conducted in location selected by parents)<br>Thematic content analysis using elements of grounded theory | Neonatal unit | Purposive sample recruited using letters from attending neonatologist and former director of neonatology. | 4 | Good |
| Ignell Modé 2014 | To explore fathers' perception of information received during their infants' care at a NICU | Sweden (high) | Not reported | Qualitative interviews (conducted in hospital whilst infant was admitted)<br>Thematic content analysis | Neonatal unit | Strategic sampling to obtain varied father-infant pairs from two NICUs; no further details reported | 4 | Good |
| Jantsch 2021 | To analyse the Health Care Network (dis)articulation of late and moderate premature infants in the first year of life | Brazil (upper-middle) | Not reported | Qualitative interviews (conducted at family home)<br>Thematic content analysis | Community | Convenience sample recruited from the Obstetric Center with potential participants identified from the birth registration book | 4 | Good |
| Kim 2020 | To assess mothers' perspectives on their NICU experiences and their unmet needs within the South Korean cultural context | South Korea (high) | Nov 2017-Jan 2018 | Free text comments in a questionnaire (delivered online)<br>Thematic content analysis, guided by critical incident technique (CIT) method | Neonatal unit | Survey conducted using participants recruited via online postings on three community portals exclusively by parents of preterm infants. Mothers with preterm infants in four hospitals also recruited in person by author at NICU discharge. All mothers provided with \$4 gift certificate incentive. | 4 | Good |
| Klawetter 2019 | To contribute an in-depth understanding of maternal engagement and the NICU experience from the perspective of mothers of preterm infants in the NICU | USA (high) | June-Dec 2017 | Qualitative interviews (conducted in hospital whilst infant was admitted)<br>Thematic analysis | Neonatal unit | Purposive sampling from two NICUs; \$20 gift card incentive given to participants | 4 | Good |
| Leonard 2008 | To explore parents' lived experience of providing kangaroo care to their preterm infants in a tertiary hospital in Cape Town | South Africa (upper-middle) | Not reported | Qualitative interviews (conducted in hospital whilst infant was admitted)<br>Thematic analysis | Neonatal unit | Purposefully sampled parents from neonatal nursery and kangaroo care ward at a tertiary maternity hospital | 4 | Good |
| Lian 2020 | To explore coping strategies of fathers of very low birth weight infants in NICU | Singapore (high) | Feb -Nov 2016 | Qualitative interviews (conducted in hospital whilst infant was admitted)<br>Thematic content analysis | Neonatal unit | Convenience sampling in level III NICU; fathers approached after 72+ hours stay by member of clinical staff, then contacted by research team | 3 | Good |
| Liu 2019 | To explore support for mothers and fathers in single-family rooms of a NICU | Canada (high) | July 2017-May 2018 | Qualitative data collected through diary voice app (for 48 hour period)<br>Thematic content analysis | Neonatal unit | Convenience sampling from a level III NICU; identified by NICU nurses who assisted with recruitment | 4 | Good |
| Lomotey 2020 | To describe the lived experiences of mothers with preterm infants at a Mother and Baby Unit of a tertiary hospital | Ghana (lower-middle) | Not reported | Qualitative interviews (conducted in hospital whilst infant was admitted)<br>Thematic analysis | Neonatal unit | Purposive sampling on the preterm unit; mothers recruited by researchers | 4 | Good |
| Lorié 2021 | To explore parents' needs and perceived gaps concerning communication with healthcare professionals during their preterm infants' admission to NICU | The Netherlands (high) | April – May 2020 | Qualitative interviews (conducted using online video conferencing tool)<br>Thematic analysis | Neonatal unit | Assisted by 'Kleine Kanjers' support network through online announcement; parents consecutively recruited based on order of registration | 4 | Good |
| Lundqvist 2019 | To present parents' lived experience of having a preterm infant cared for at the neonatal unit until discharge from hospital-based neonatal home care | Sweden (high) | Not reported | Qualitative interviews (conducted at the family home)<br>Phenomenological analysis | Neonatal unit & community | Recruited by three nurses in a level IIb NICU; 19 couples interviewed as part of a broader study, and the 6 couples providing the richest narrative included in this analysis | 3 | Good |
| Mihae 2021 | To clarify and define the concept of nursing support as perceived by mothers of preterm infants | South Korea (high) | Nov 2017-March 2018 | Qualitative interviews (location not reported)<br>Thematic analysis | Neonatal unit | First participant enrolled by nurse at NICU, then snowball sampling | 4 | Good |
| Ncube 2016 | To explore and describe the lived experiences of mothers with regard to the care of their hospitalised preterm | Botswana (upper-middle) | Dec 2010-Jan 2011 | Qualitative interviews (conducted in hospital whilst infant was admitted) | Neonatal unit | Purposive sampling from a NICU in a referral hospital; | 4 | Good |
|  | infants, in a NICU where mothers had restricted interaction with their preterm infants |  |  | Thematic analysis |  | recruited by duty nurse who informed mothers of study |  |  |
| Neu 2020 | To compare mothers' experiences in NICUs where family-centred care is the standard of care and to compare these with the experiences of mothers two decades ago | USA (high) | Not reported | Qualitative interviews (conducted in hospital whilst infant was admitted)<br>Thematic analysis | Neonatal unit | Purposive sampling from two NICUs in teaching hospitals; no further details reported | 4 | Good |
| Norén 2018 | To describe mothers' experiences of providing their preterm infants with Kangaroo Mother Care | Sweden (high) | 2009 | Qualitative interviews (conducted at the family home)<br>Content analysis | Neonatal unit & community | Consecutively recruited from two level 3 NICUs; no further details reported | 4 | Good |
| Nyondo-Mipando 2020 | To explore the experiences of caregivers in the implementation of Kangaroo Mother Care | Malawi (low) | April-June 2019 | Qualitative interviews and participant observation (in hospital whilst infant was admitted)<br>Thematic analysis | KMC ward | Purposive sampling, recruited by researchers supported by nursing officers in four hospitals (one tertiary, three secondary) | 3 | Good |
| Olsson 2017 | To describe fathers' experiences of skin-to-skin contact with their premature infant | Sweden (high) | Jan 2014-June 2015 | Qualitative interviews (conducted in hospital [n=19], or family home [n=1])<br>Thematic content analysis | Neonatal unit | Purposeful sample by designated nurses in the two neonatal units (one county, one university) to achieve maximum variation in demographics | 4 | Good |
| Orapiriyakul 2007 | To explore how mothers in Thailand develop maternal attachment to infants born preterm and requiring NICU hospitalisation | Thailand (upper-middle) | June 2005-Aug 2006 | Qualitative interviews (location not reported) and participant observation (in NICU)<br>Constant comparative analysis | Neonatal unit | Three participants purposively sampled in two NICUs (one public university, one provincial hospital), then subsequent theoretical sampling | 3 | Good |
| Petty 2018 | To gain insight into the post-discharge experiences of parents in relation to the adequacy of preparation for caring for extremely premature infants at home | UK (high) | Sept-Nov 2017 | Qualitative interviews (conducted at the family home)<br>Constant comparative analysis | Discharge from NICU & community | Purposive sampling via the coordinator of an NHS trust-based parent support group | 4 | Good |
| Petty 2019b<br>(2 papers) | To explore the narratives of parents to enable practitioners to understand what it is like to live through a period of neonatal care with their premature infant | UK<br>(high) | Not reported | Qualitative interviews<br>(conducted at the family home or a private location)<br>Thematic analysis | Neonatal unit,<br>discharge from NICU & community | Purposive sampling; volunteers were requested through a key gatekeeper linked to a UK parent support charity who disseminated the recruitment call nationally using their established email contact database. | 4 | Good |
| Premji 2017 | To explore mothers' experiences of caring for their late preterm infants in the community | Canada<br>(high) | April 2013-June 2014 | Qualitative interviews<br>(conducted at the family home or a private location)<br>Interpretative thematic analysis | Community | Multistage purposeful sampling to achieve rich narratives and diversity from four hospitals in Calgary; \$50 grocery gift card incentive | 3 | Good |
| Rossmann 2011 | To describe the experiences of mothers of VLBW infants who received lactation care from certified breastfeeding peer counsellors with special preparation for NICU care | USA<br>(high) | Oct 2008-Mar 2009 | Qualitative interviews<br>(conducted in hospital whilst infant was admitted)<br>Thematic content analysis | Neonatal unit | Convenience sampling, approached by NICU practitioner at a tertiary care NICU | 3 | Good |
| Russell 2014<br>(2 papers;<br>also Sawyer 2013) | To explore parents' views and experiences of the care for their very premature infant on NICU (Russell 2014)<br>To explore parents' experiences and satisfaction with care during very preterm birth and to identify domains associated with positive and negative experiences of care (Sawyer 2013) | UK<br>(high) | June – Nov 2011 | Qualitative interviews<br>(conducted in hospital or in the family home)<br>Inductive thematic analysis | Neonatal unit | Recruited from three tertiary care centres in England using posters in NICU or posted/personally-given letters; parents returned a card if they wanted to participate | 4 | Good |
| Skene 2012 | To explore how parents interact with their infants and nurses regarding the provision of comfort care in a NICU | UK<br>(high) | Jan – Nov 2008 | Focussed ethnography, with participant observations and qualitative interviews (in NICU)<br>Inductive thematic analysis | Neonatal unit | Approached by researcher in a regional NICU | 5 | Good |
| Treherne 2017 | To discover parents' perceptions of closeness to and separation from their preterm infants in the NICU | Canada<br>(high) | Feb 2015 –<br>Jan 2016 | Qualitative data collected through diary voice app (for 24 hours)<br>Thematic content analysis | Neonatal unit | Purposive sampling in an urban level III NICU, with parents approached by NICU staff | 4 | Good |
| Unsworth 2021 | To explore caregiver experiences and healthcare provider perspectives of accessing healthcare for low birth weight infants in rural Kenya | Kenya (lower-middle) | June 2019 | Qualitative interviews (conducted within the research area of the hospital)<br>Thematic analysis | Community | Convenience sampling from neonatal and postnatal ward registers at a county hospital or those known to community health volunteers; contacted by telephone or home visits | 4 | Good |
| Veronez 2017 | To describe the maternal care process mediated by nurses during the period of hospitalisation and discharge of premature infants | Brazil (upper-middle) | Oct-Dec 2011 | Daily journal written by mothers<br>Thematic content analysis | Neonatal unit | Recruited from neonatal units by researcher who was a member of nursing staff | 3 | Good |
| Villeneuve 2018 | To identify outcomes that were important to families of children requiring neonatal care | UK (high) | 2016-2017 | Qualitative interviews (conducted at a university, in the family home, or in a children's centre)<br>Thematic analysis | Neonatal unit | Participants purposively sampled from: lists held by neonatal support groups (BLISS, SNUG); conference on neonatal services; families known to NICU at Royal Devon and Exeter NHS Trust | 5 | Good |
| Wernet 2015 | To analyse the maternal experience in a neonatal intensive care unit, focusing on relations of recognition | Brazil (upper-middle) | May – August 2013 | Qualitative interviews (location not reported)<br>Thematic analysis | Neonatal unit | Mothers recruited if preterm neonates had left the regional NICU less than one month prior to the interview; no further detail given on recruitment | 4 | Good |
| Yu 2020 | To explore Chinese parents' experiences and expectations of having preterm infants in a Chinese NICU | China (upper-middle) | Jan – May 2018 | Qualitative interviews (conducted in hospital)<br>Thematic analysis | Neonatal unit | Purposively sampled by first author from a NICU in a tertiary hospital | 3 | Good |

\* According to the World Bank's income classification 2021-2022 (https://datatopics.worldbank.org/world-development-indicators/the-world-by-income-and-region.html)
Abbreviations: CASP = Critical Appraisal Skills Programme; NICU = Neonatal Intensive Care Unit; NHS = National Health Service; SCBU = Special Care Baby Unit; VLBW = very low birthweight

34 studies focused on in-hospital care in a neonatal unit, although the level of care provided at these units varied by setting. Four studies examined views of community care, and eleven studies collected data on more than one setting or aspect of care (including transition from hospital to community care, or transfers between hospitals). The selected studies scored highly on data richness (38 scored four or five) and quality (46 assessed as “good”). All three studies assessed as being of “average” quality scored four for data richness.

Table 3 (next page) shows the characteristics of the study participants. Our analysis is based on the views of 1,143 caregivers from the primary studies. 18 studies included mothers only, six recruited fathers only, 23 included parents, and two included grandparents. The patient population was preterm infants in 29 studies, preterm and/or low birth weight in 19 studies, and low birth weight infants in one study. Studies used different gestational age limits, although all were births under 37 weeks gestation. Ten included only very preterm (<32 weeks) infants, and three included only extremely preterm (<28 weeks) infants. Specified birthweight limits, observed birthweight ranges, the ages of the infants/children at the time of the study, and characteristics of the carers were not consistently reported. Four of the included studies collected some of their data when the children were older than 24 months, with the oldest children being 6 years old.

**Table 3.** Characteristics of the study populations in sampled studies (n=49)

| Author / Date | Inclusion/exclusion criteria | Sample size | Patient population | Gestational age range at birth | Birthweight range | Age of child at data collection | Other information |
| --- | --- | --- | --- | --- | --- | --- | --- |
| Abeasi 2020 | **Included:** Mothers who had delivered an infant at < 37 weeks; infant had been in the hospital for >5 days; infant's condition had improved if the infant had been very ill; and mother was not too anxious for an interview | 12 | Preterm and/or LBW | 30-35 weeks | 1800g-2200g | Not reported | Women aged 20-36; all had completed secondary (n=7) or tertiary (n=5) education |
| Adama 2017 | **Included:** Fathers who were aged 18+; their preterm infants had no disabilities | 9 (interviewed 3 times) | Preterm | 26-36 weeks | Not reported | Not reported | Men aged 20-38; all in full time employment |
| Amorim 2019 | **Included:** Parents whose very preterm infants survived; who were present in NICU during the hospitalisation period; able to speak and write in Portuguese; infant discharged and alive | 52 (26 couples) | Preterm and/or LBW | <28 weeks | Not reported | Not reported | Not reported |
| Arnold 2013 | **Included:** Parents who could speak fluent English and had a preterm baby born prior to 32 weeks gestation in a 6-month period (Jan-June 2011) | 39 (32 mothers, 7 fathers) | Preterm | 24-31 weeks | Not reported | 44 – 344 days | Parents aged 25-44; 29 of 39 were White European; 37 were married or cohabiting |
| Aydon 2018 | **Included:** Parents with babies admitted to the neonatal clinical care unit whose gestation was between 28-32 weeks<br>**Excluded:** Parents with babies born with anomalies and/or not expected to survive; non-English speaking; or potentially difficult to follow-up due to involvement with the child and family protection services | 40 (20 couples, interviewed separately) | Preterm | 28-32 weeks | Not reported | 4-6 weeks old and 4-6 weeks post-discharge | Mothers aged 21-42; fathers aged 21-43; 33 of 40 were first-time parents; 17 of 20 couples were married |
| Blomqvist 2012 | **Included:** Fathers of an infant born at 28- 33 weeks; infants did not have a life-threatening condition | 7 | Preterm and/or LBW | 29-33 weeks | 1315-2500g | 4 months +/- 2 weeks (corrected age) | Fathers aged 25-36; all first-time fathers; all married or co-habiting with mother |
| Brødsgaard 2015 | **Included:** Parents who had participated in the Early Discharge Programme; infant born Jan - June 2012; gestational age 25-36 weeks; singleton or twins, first-time and experienced parents; able to understand and speak Danish well enough to participate actively in group discussions | 15 (2 focus groups) | Preterm | 28-34 weeks | 1230-2800g | Approximately 6 months old | Mothers aged 29-39; fathers aged 34-42 |
| Chang Lee 2009 | **Included:** Mothers of infants whose birth weight was < 1500g, and who were Taiwanese | 26 | Preterm and/or LBW | 25-34 weeks | 530-1490g | Not reported | Mothers aged 22-36 |
|  | **Excluded:** Single mothers; teenage mothers (aged < 20); foreign mothers; mothers not physically or mental fit to be interviewed; multiple births; and infants with life-limiting illness or congenital abnormalities |  |  |  |  |  |  |
| Dorner 2020 | **Included:** English-speaking parents of infants born preterm who, at the time of interview: (1) were between 28 and 34 weeks' corrected age, and (2) had not yet been screened with General Movements Assessment examinations | 19<br>(15 mothers,<br>4 fathers) | Preterm | Median 29.6<br>(IQR 25.7 –<br>31.2) | Median 1175g<br>(IQR 740-1735g) | Not reported,<br>although may<br>be ~ 4 weeks<br>after birth | Median age of parents 32<br>(IQR 27-38); 14 of 19 had<br>been in higher education;<br>11 of 19 first-time parents |
| dos Santos<br>2014 | **Included:** Mothers of preterm infants, born at < 32 weeks and/or weighing < 1,500g; admitted to the NICU of the University Hospital of Londrina; who participated in the project "A support network for the premature infant's family" | 21 | Preterm<br>and/or LBW | Range not<br>reported | Range not<br>reported | Up to 6<br>months<br>(chronological<br>age) | Mothers aged 14-42; 50%<br>married; 60% had other<br>children and < 8 years of<br>education |
| Feeley 2013 | **Included:** Infant's biological father; lived with infant's mother; infant had been hospitalised for 7+ days; infant's medical condition was stable; could communicate in English or French<br>**Excluded:** Fathers with a previous experience of NICU; infants with grade 3-4 intraventricular haemorrhage or major congenital anomaly | 18 | Preterm | Mean 28<sup>+6</sup><br>weeks | Mean 1173g | Mean age 55<br>days | Mean age of fathers 37.7; 8<br>of 18 has university<br>education; all were<br>currently employed |
| Fernandez<br>Medina<br>2021 | **Included:** Parents > 18 years old; with an extremely preterm infant who had been discharged from NICU with some type of technological dependency during the last 24 months<br>**Excluded:** Parents of infants with a congenital disease | 17<br>(12 mothers,<br>5 fathers) | Preterm | 24-27 weeks | Not reported | Not reported;<br>infants<br>discharged in<br>last 24 months | Mean age of parents 34.2;<br>12 of 17 were married; |
| Finlayson<br>2014 | **Included:** English-speaking mothers; > 16 years; infant treated on the unit for 7 days or more<br>**Excluded:** Mothers whose infants were receiving intensive care at the time of interview | 12 | Preterm<br>and/or LBW | 25-31 weeks | 595-1517g | Not reported | Mothers aged 21-40; 11 of<br>12 identified as White-British; 6 of 12 university<br>educated; all had partners |
| Franck 2017 | **Included:** Parents whose babies had received NICU care within the past 3 years | 40<br>(33 mothers,<br>7 fathers) | Preterm | Not<br>reported | Not reported | Not reported;<br>NICU care was<br>received in<br>past 3 years | 47% of participants were<br>aged >35 & 45% were aged<br>26-35; 85% were married;<br>90% were employed |
| Gallegos-Martinez<br>2013 | **Included:** Parents with a preterm baby hospitalized in the neonatal unit | 31<br>(9 mothers,<br>11 couples) | Preterm | Not<br>reported | Not reported | Not reported | 70% "nuclear" families; 89%<br>of mothers "homemakers";<br>82% of fathers were<br>labourers; high school<br>completion <35% |
| Glazer 2021 | **Included:** Mothers with a live birth between June 2016-June 2018; very preterm or very low birthweight (<1,500g) delivery with a minimum five-day NICU stay | 20 | Preterm and/or LBW | “Very preterm” | “Very low birthweight” | Not reported | 40% Black, 40% White or Asian, 20% Latina; 8 of 12 Black or Latina participants has Medicaid cover, all White or Asian participants privately insured |
| Granrud 2014 | **Included:** Parents of a premature baby born at a university hospital and transferred to a local hospital; able to speak Norwegian<br>**Excluded:** prior experiences of preterm delivery | 20<br>(2 mothers, 9 couples) | Preterm | 26-32 weeks | Not reported | Not reported | Mothers aged 22-40, fathers aged 23-40 |
| Guillaume 2013 | **Included:** Parents who spoke French; whose child was born before 32 weeks of gestation; was 15 to 30 days old at inclusion; and had no recent severe clinical aggravation | 60<br>(30 mothers, 30 fathers) | Preterm | Mean 27<sup>+2</sup> weeks<br>Maximum 31<sup>+6</sup> weeks | Mean 956g | 15-30 days old | Mothers mean age 30.7, fathers mean age 33.5; 92% employed; 8% single parents |
| Hägi-Petersen 2021 | **Included:** Parents from two neonatal wards offering early in-home care programmes with nurse-supported video consultations (programme had inclusion criteria not specified in the paper) | 11<br>(1 mother, 5 couples) | Preterm | 27-35 weeks | Not reported | Not reported; 14-30 days after discharge from in-home care programme | Mothers aged 21-41, fathers aged 24-38 |
| Hendriks 2017 | **Included:** Parents of infants born alive at <28 weeks and died in the delivery room or in the NICU from 2013 through 2015<br>**Excluded:** Participants who gave birth to two or more infants; or if infants were stillborn | 20<br>(5 mothers, 1 father, 7 couples) | Preterm and/or LBW | 22-27 weeks | 340-1100g | 1-2 years after the infant’s death | 14 of 20 aged 35+; 18 of 20 Christian |
| Ignell Modé 2014 | **Included:** Fathers of infants treated at one of two Swedish NICUs; able to speak Swedish. Infant inclusion criteria were absence of an acute life-threatening condition and a stay of at least 1 week in NICU | 8 | Preterm | 23-36 weeks | Not reported | Not reported | Fathers aged 20-24 years |
| Jantsch 2021 | **Included:** Mothers of preterm infants born May 2016 - May 2018; who lived in the city of Santa Maria; who developed chronic and acute health conditions | 15 | Preterm | 32-36 weeks | Not reported | 3-9 months | 11 of 15 in socio-economic classes C or D; 6 of 15 infants had chronic health problems |
| Kim 2020 | **Included:** Mothers of infants born <37 weeks or LBW <2500g; if admission lasted at least seven days; and if infants did not have congenital anomalies and did not require life-prolonging treatment | 232 | Preterm and/or LBW | Mean 30<sup>+3</sup> weeks | Mean 1523g | Up to 18 months | Mothers mean age 34.19; 99% married; 99% with high school education or above |
| Klawetter 2019 | **Included:** English-speaking mothers of infants born at <32 weeks; hospitalised in NICU 2+ weeks; 33-34 weeks at the time of interview<br>**Excluded:** Mothers <18 years; with a diagnosed psychiatric disorder and/or recorded or stated illicit substance use | 14 | Preterm | Maximum 31<sup>+6</sup> weeks | Not reported | 33-34 weeks corrected gestational age | 9 of 14 mothers were White, 3 African-American; 7 of 14 had Medicaid cover; 13 educated to high-school level or higher; 11 were married |
| Leonard 2008 | **Included:** Parents of preterm infants receiving kangaroo care at the hospital at the time of the interview; able to speak English; singleton births; >7 days old; weight > 1000g; not receiving critical care (intubation and/or life support) | 6<br>(4 mothers, 2 fathers) | Preterm | Not reported | Minimum 1000g; maximum not reported | Not reported | 5 married or co-habiting, 1 single mother |
| Lian 2020 | **Included:** Fathers with full custody of their infants<br>**Excluded:** Fathers whose infants died during the NICU stay; were unable to give informed consent; or unable to comprehend interview questions in English | 15 | Preterm and/or LBW | 25-34 weeks | 580-1474g | Not reported | 9 of 15 aged 31-40; all married; 10 of 15 educated at degree level; 13 of 15 employed full-time |
| Liu 2019 | **Included:** Parents visiting their infant; able to speak and read English or French; aged > 18; infants hospitalised for 48+ hours<br>**Excluded:** Infants with grade 4 intraventricular haemorrhage, trisomy 21 or other chromosomal abnormalities, in palliative care, and/or was being placed in foster care | 15<br>(9 mothers, 6 fathers) | Preterm | 24-35 weeks | 620-3605g | 5-108 days | Mothers mean age 34.6; fathers mean age 35; all married or cohabiting; 14 of 15 had college or degree education; 4 of 14 unemployed |
| Lomotey 2020 | **Included:** Mothers with babies born <37 weeks<br>**Excluded:** Mothers whose preterm babies were critically ill or had congenital anomalies | 10 | Preterm | 26-36 weeks | Not reported | Not reported | Mothers aged 17-38 years; all educated to junior high school level; 9 were Christian; all married or cohabiting |
| Lorié 2021 | **Included:** Parents of preterm infants (born < 37 weeks' gestation); admitted to a Dutch level 2-4 NICU for 1+ week; able to speak Dutch | 20<br>(19 mothers, 1 father) | Preterm | 24-35 weeks | Not reported | 1-5 years | Parents aged 28-38 years; living across the Netherlands; used 12 different NICUs |
| Lundqvist 2019 | **Included:** Parents of infants born <37 weeks; who had ended the care period in hospital-based neonatal home care; able to communicate in Swedish or English | 12<br>(6 couples) | Preterm | 26-36 weeks | 655-3200g | Not reported | Mothers aged 30-40, fathers aged 32-43; 3 couples from urban and 3 from rural area |
| Mihae 2021 | **Included:** Mothers who understood the purpose of the study and agreed to participate; with infants born <37 weeks; | 10 | Preterm and/or LBW | 24-34 weeks | 655-2000g | 1-10 months | Mothers aged 22-43; all married; 7 of 10 first-time mothers |
|  | without congenital deformities and hereditary diseases; and within a year of childbirth |  |  |  |  |  |  |
| Ncube 2016 | **Included:** Mothers of singleton infants born <37 weeks; hospitalised for 5+ days but with stable or improving health; able to speak Setswana or English<br>**Excluded:** Mothers of infants acutely ill at the time of data collection; or whose infant had a congenital abnormality | 8 | Preterm | Not reported | Not reported | Not reported | Mothers aged 23-30 |
| Neu 2020 | **Included:** Mothers of infants born <32 weeks; at least 33 weeks postconceptional age; hospitalized for 2+ weeks<br>**Excluded:** Mothers diagnosed with a psychiatric disorder such as bipolar disorder or schizophrenia and/or recorded or stated illicit substance use | 14 | Preterm | <32 weeks | Not reported | At least 33 weeks corrected age | Mothers mean age 28; 9 of 14 White, 3 of 14 Black or African American; 10 had college or degree level education; 11 married; 10 working full-time |
| Norén 20108 | **Included:** Mothers of singleton infants born at 28-33 weeks; whose condition was not life-threatening; able to speak Swedish | 13 | Preterm and/or LBW | 29-33 weeks | 1175-2500g | 4 months (+/- 2 weeks) corrected age | Mothers aged 25-42; all married or cohabiting with father; 8 of 13 first-time mothers |
| Nyondo-Mipando 2020 | **Included:** Caregivers of preterm or LBW infants who were in a stable condition; were providing KMC; and had been in the KMC ward for 5+ hours | 24 (14 mothers, 6 fathers, 3 grand-mothers, 1 grand-father) | Preterm and/or LBW | Not reported | <2500g | Not reported | Not reported |
| Olsson 2017 | **Included:** Fathers of preterm infants; who had provided skin-to-skin care to the infant at least once | 20 | Preterm | 25-35 weeks | Not reported | 2-74 days old | Fathers aged 23-45 |
| Orapiriyakul 2007 | **Included:** Thai mothers; living with partners/ husbands; with preterm infants <37 weeks by Ballard scores assessment; birth weight <1500g; no congenital anomaly; requiring mechanical ventilation and hospitalized in NICU | 15 | Preterm and/or LBW | 26-33 weeks | 740-1400g | 2-33 days | Mothers aged 16-41; 12 of 15 were Buddhists; 5 of 15 educated to diploma or degree level; 12 reported low family income |
| Petty 2018 | **Included:** Parents whose infants were born at ≤30 weeks; who had been discharge home within the previous one to six years | 15 (12 mothers, 1 father, 1 couple) | Preterm and/or LBW | 24-30 weeks | 615-1600g | 1-6 years | Parent characteristics not reported |
| Petty 2019b<br>(2 papers) | **Included:** Parents whose infants were born at <37 weeks; had spent more than a week receiving neonatal care; and who had been discharged home in the preceding ten years | 23<br>(16 mothers,<br>1 father, 3<br>couples) | Preterm<br>and/or LBW | 24-32 weeks | 500-1500g | 1-10 years | Parent characteristics not reported |
| Premji 2017 | **Included:** Mothers of late preterm infants regardless of mode of delivery and admission status (newborn nursery, secondary hospital or NICU, or length of stay)<br>**Excluded:** Mothers unable to read/write English; or could not be contacted in time for them to complete the maternal confidence in care survey | 11 | Preterm | 34-36 weeks | 1822-3630g | Not reported | Mothers mean age 31.1; all married; 6 of 10 had completed higher education; 8 of 10 born in Canada; 6 of 10 White |
| Rossmann<br>2011 | **Included:** Mothers of a VLBW infant in the NICU who was expected to survive; maternal age 18+ years; able to speak and understand English; 3+ interactions with a breastfeeding peer counsellor | 21 | Preterm<br>and/or LBW | 24-31 weeks | 511-1460g | 12-80 days | Mothers mean age 29.3; 15 of 21 African-American; 11 of 21 married; 17 of 21 had some college education; 12 employed full-time |
| Russell<br>2014;<br>Sawyer 2013 | **Included:** Parents whose infants were born at <32 weeks (also birth <6 months previously in Sawyer); had been on neonatal unit for 2+ weeks; spoke English well; at least one member of the couple wanted to participate or they were single; included parents of babies who had died | 39<br>(32 mothers,<br>7 fathers) | Preterm | 24-31<sup>+6</sup><br>weeks | Not reported | 44-344 days<br>(Russell); <6<br>months<br>(Sawyer); 2<br>babies had<br>died | Parents aged 25-44; 37 of 39 married or co-habiting; 29 White European; 33 employed |
| Skene 2012 | **Included:** Parents >16 years; considered as suitable for inclusion by the nurse in charge | 19<br>(2 mothers,<br>1 father, 8<br>couples) | Preterm<br>and/or LBW | 23-32 weeks | 520-1615g | 5-31 days at<br>recruitment | Majority of parents were White British |
| Treherne<br>2017 | **Included:** Parents whose infants were born at <37 weeks; hospitalised in the NICU; infant was stable; able to read English or French; able to provide informed consent | 20<br>(13 mothers,<br>7 fathers) | Preterm | 24-33 weeks | 615-3030g | 8-94 days | Mothers mean age 32.2; fathers mean age 37.3; 10 of 20 had university degrees; all in employment |
| Unsworth<br>2021 | **Included:** Caregivers of LBW infants < 24 months of age | 11<br>(all female,<br>roles not<br>specified) | Low birth<br>weight | Not<br>reported | Not reported | <24 months | 9 of 11 caregivers aged >25 |
| Veronez<br>2017 | **Included:** Mothers of infants born at <37 weeks; birth weight more than 1500g; hospitalisation time ≥72 hours; and residents of the municipality of Maringá or in the 15<sup>th</sup> health region | 7 | Preterm<br>and/or LBW | 31-36 weeks | 1560-2460g | Not reported | Mothers aged 16-31 |
| Villeneuve 2018 | **Included:** Parents who had used a neonatal service and been discharge from the service within the last 6 months to 5 years | 12<br>(8 mothers, 2 couples) | Preterm | 24-34 weeks | Not reported | 6 months to 5 years post-discharge | For “half of the families”, this was their first child |
| Wernet 2015 | **Included:** Mothers of infants born at $\leq 34$ weeks; without any congenital syndromes; who stayed in the NICU for at least one week<br>**Excluded:** Mothers who had another preterm child; those with any mental health problems | 10 | Preterm | 26-34 weeks | Not reported | Roughly aged up to 5 months | 6 of 10 mothers aged 18-25, 3 aged 25-30, 1 over 40 years; 5 of 10 were first-time mothers; 7 of 10 lived with partners and 3 were single mothers |
| Yu 2020 | **Included:** Parents of a preterm infant; admitted to the NICU for 7+ days; infant’s condition was stable; parents were aged 18+; and were the primary caregiver<br>**Excluded:** History of mental illness; infants were abandoned or deceased | 15<br>(10 mothers, 5 fathers) | Preterm | 27-36 weeks | 1010-2850g | Not reported | Mean age of parents 31.1 years; 11 of 15 were first-time parents; 8 of 15 had completed high school, 4 of 14 had completed a degree |

### Analysis findings

We identified 31 descriptive themes, 21 of which we graded with high confidence, and nine with moderate confidence. Only one theme was graded with low confidence. We grouped these descriptive themes into eight overarching analytical themes. Table 4 (next page) shows the framework of themes, associated CERQual gradings and supporting data. What mattered to carers was a positive outcome for the child; active involvement in care; support to cope at home after discharge; emotional support for the family; the healthcare environment; information needs were met; logistical support was available; and positive relationships with staff. The full GRADE-CERQual evidence profiles for each descriptive theme are shown in Supplementary Tables 2-9. Additional evidence from the primary studies is presented in Supplementary Tables 10-17.

**Table 4.**
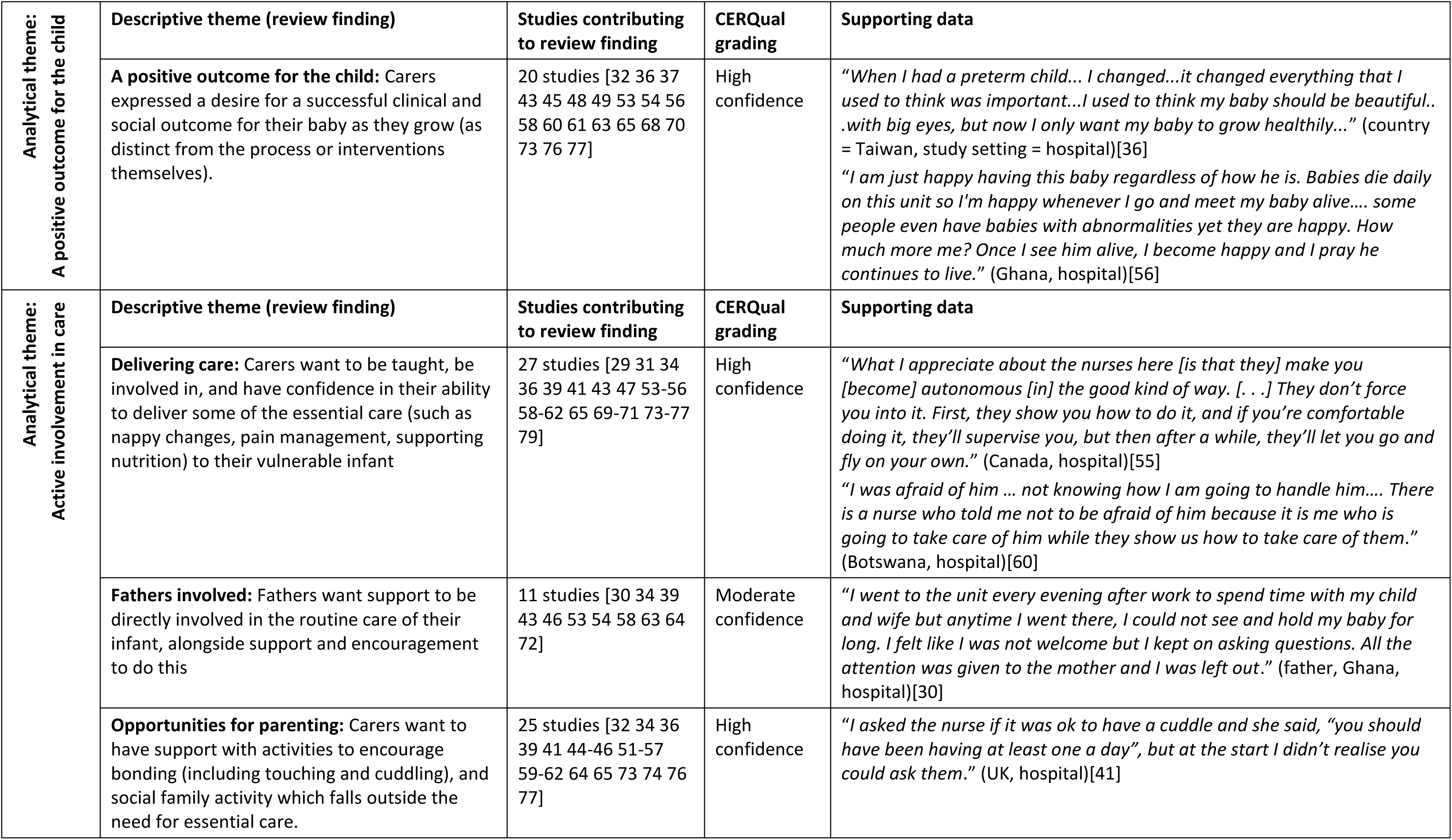
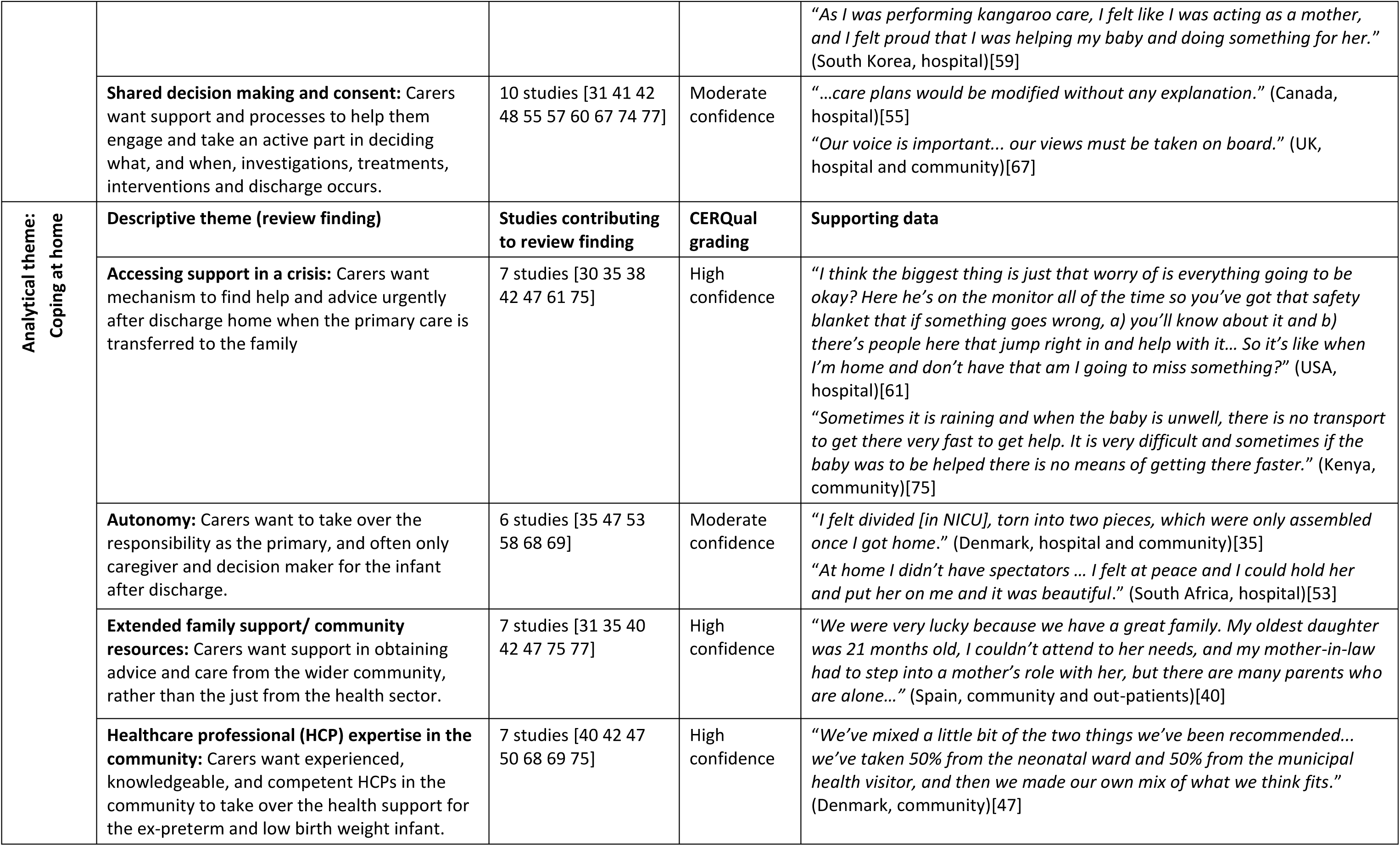
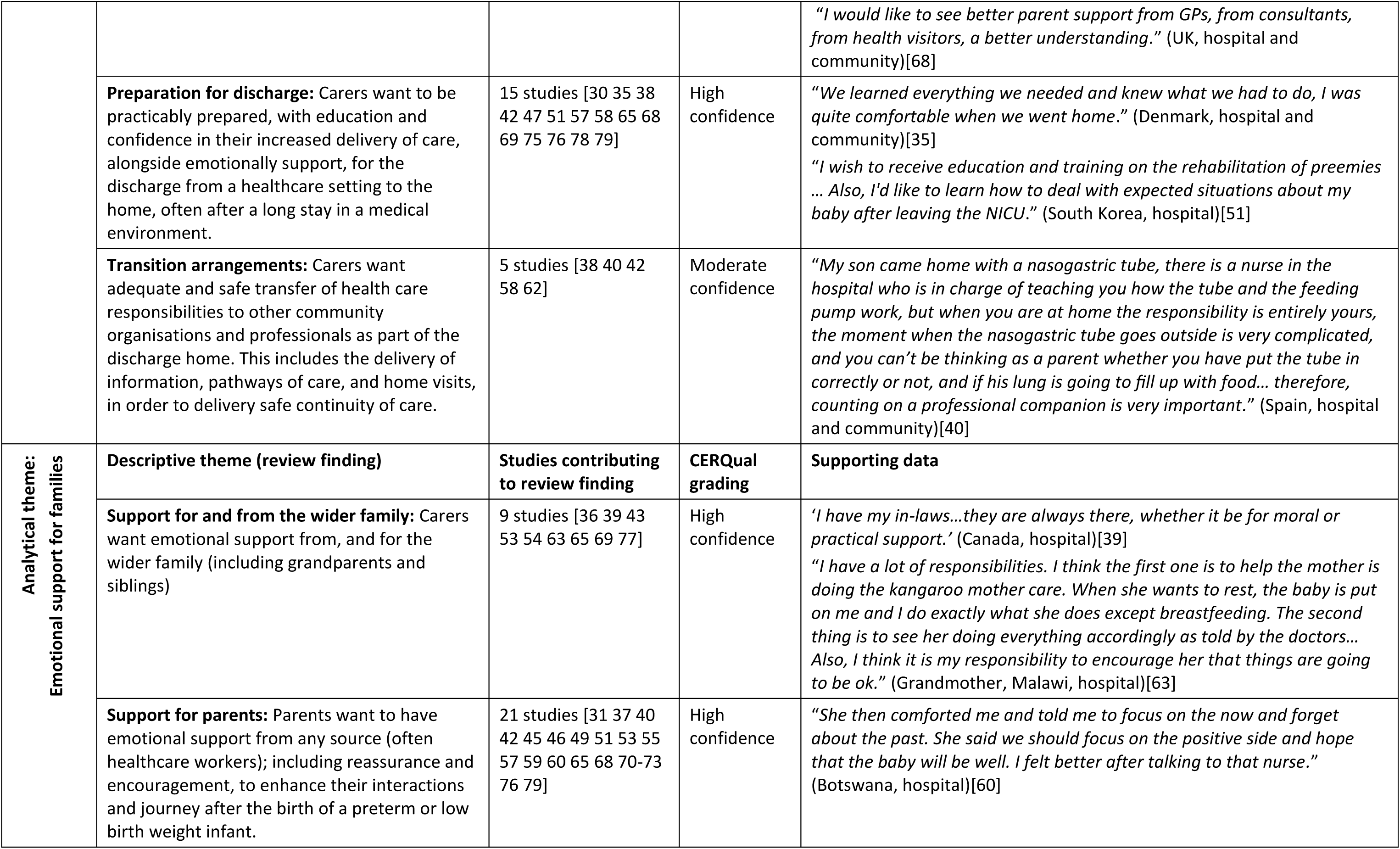
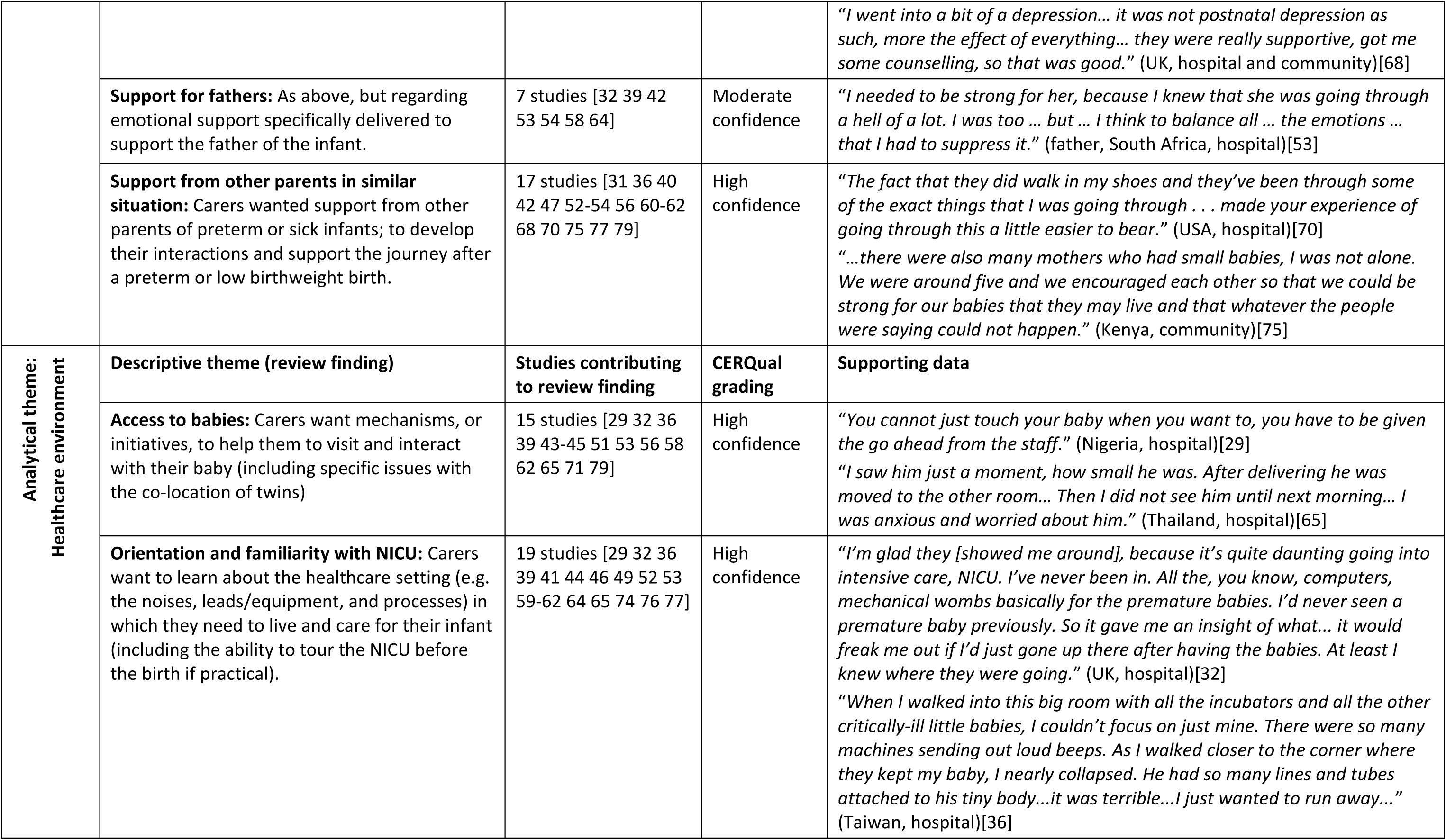
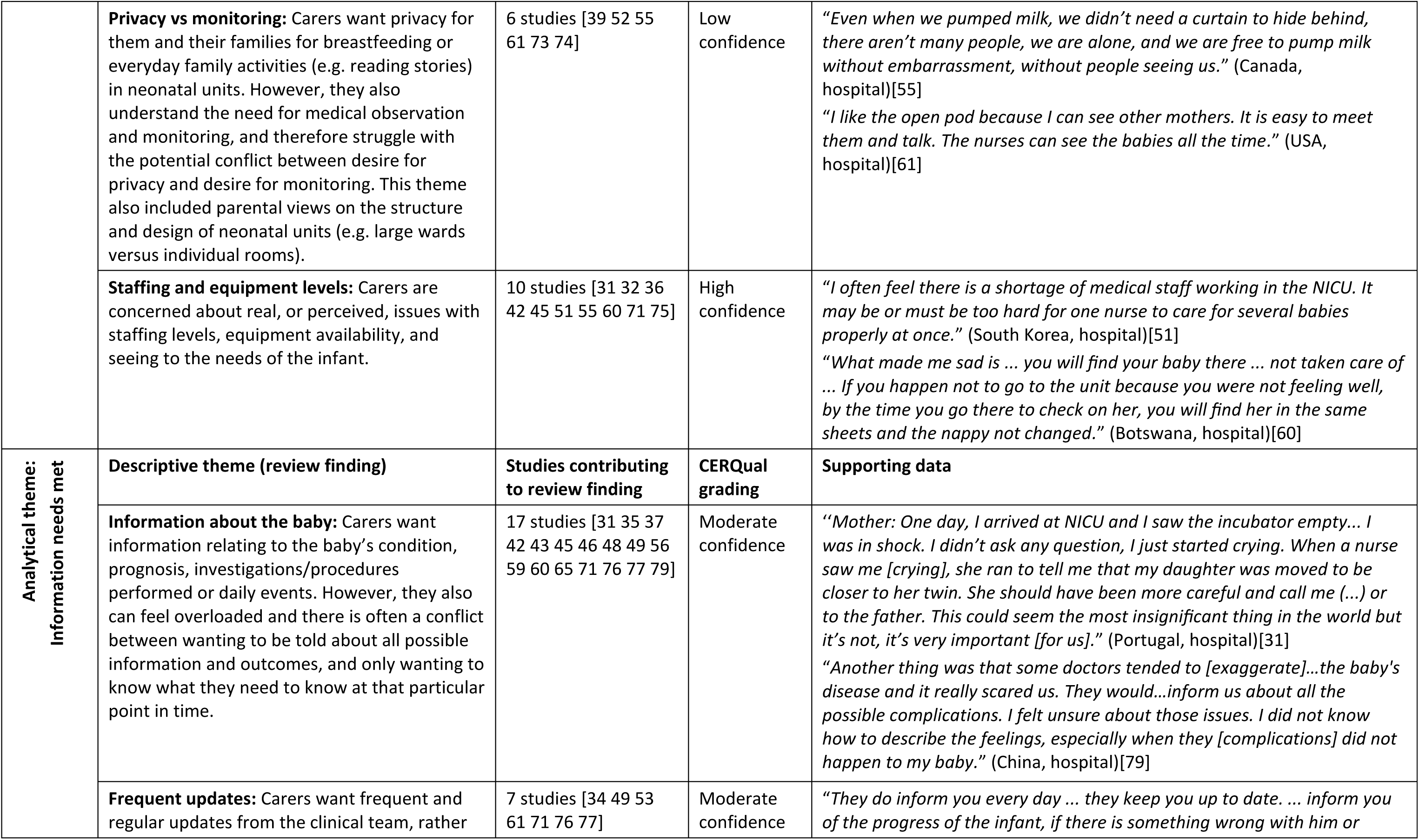
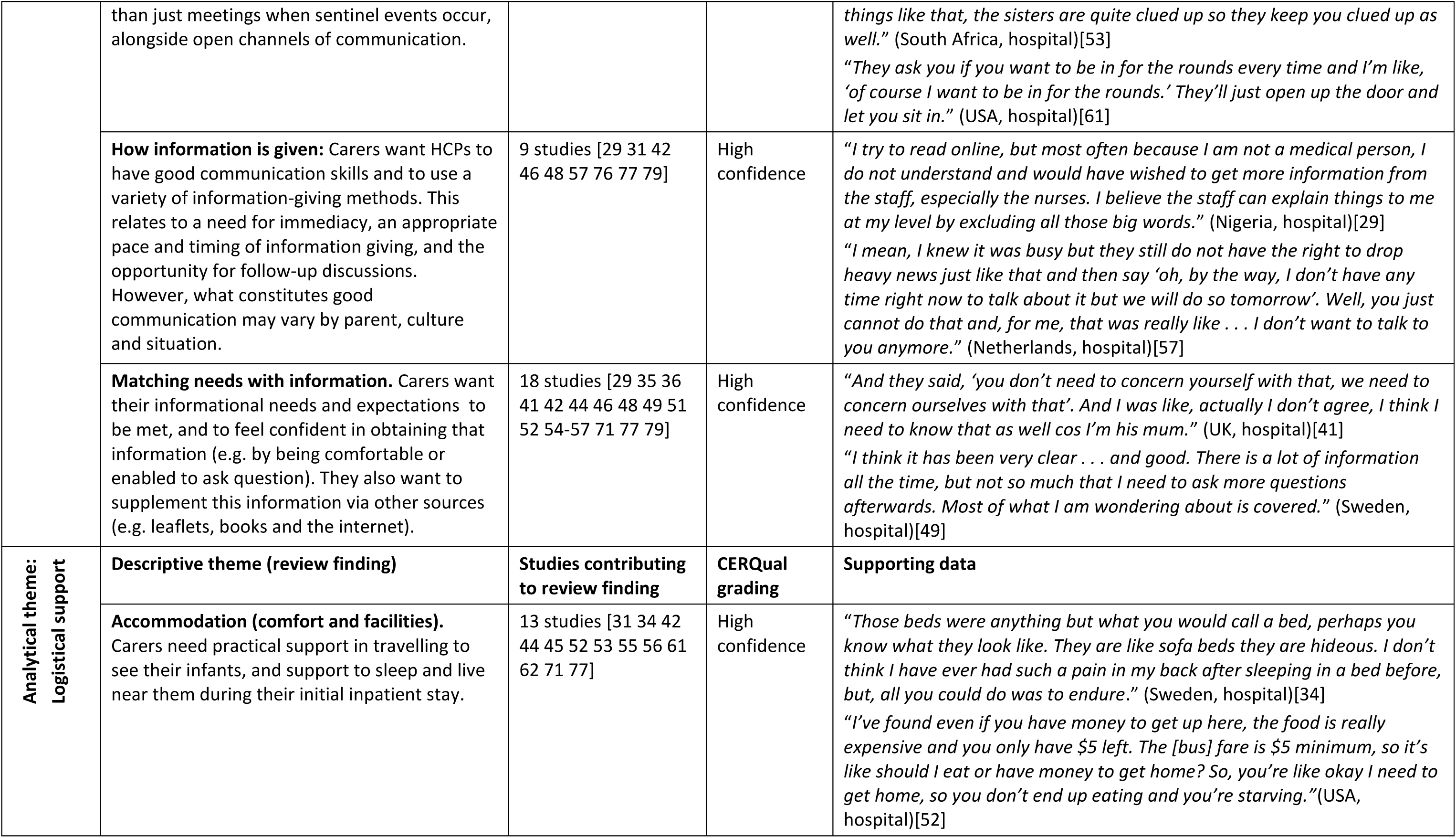
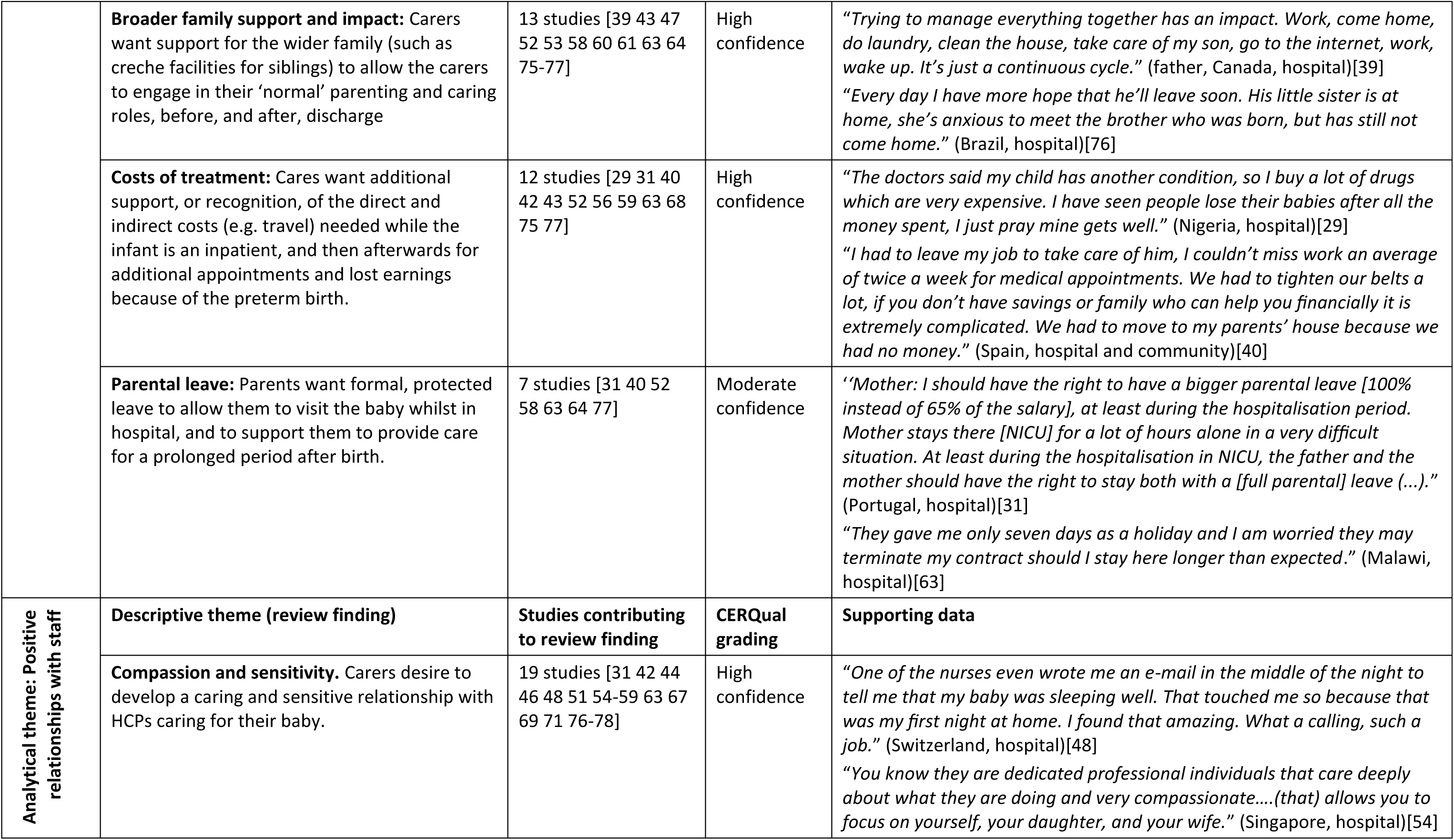
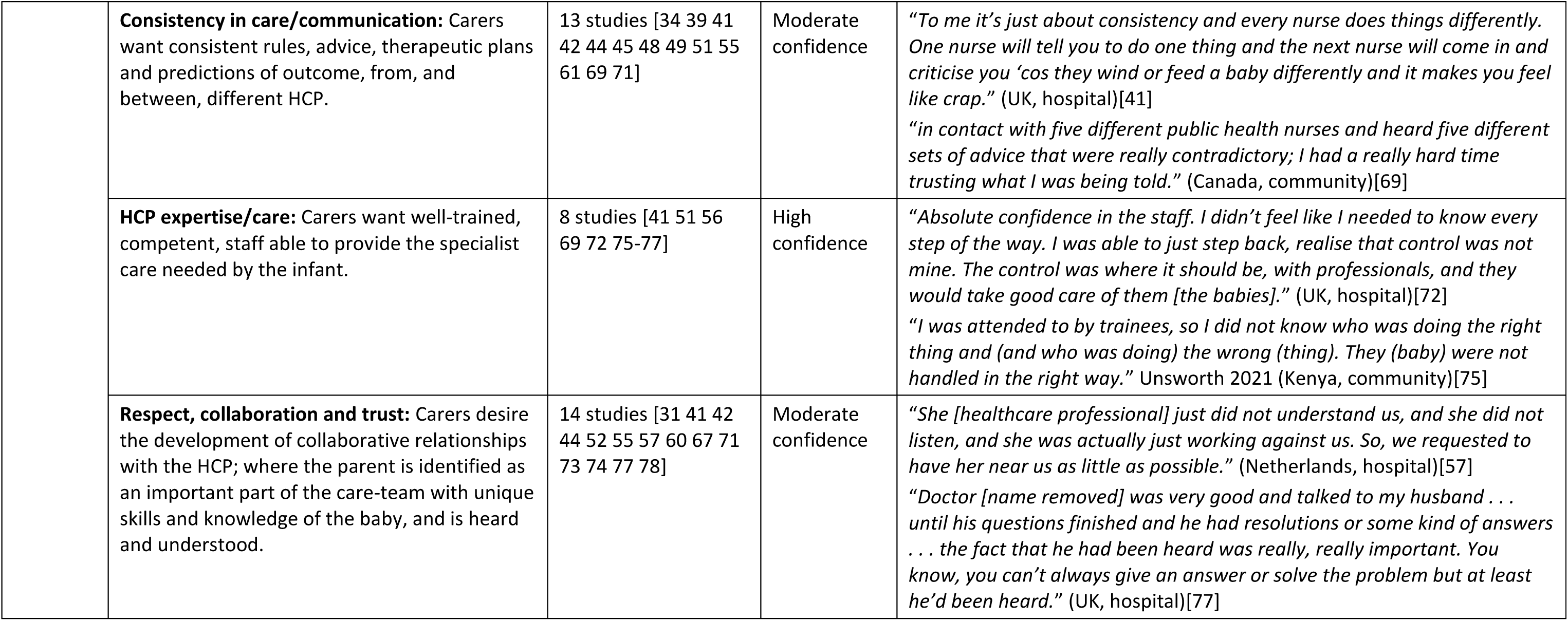
Framework of themes emerging from the data

We now discuss each of the eight analytical themes in more detail. They are listed in alphabetical order to avoid undue emphasis being placed on one over another because of the team’s biases and prior beliefs.

### A positive outcome for the child

20 studies contributed data to this analytical theme, with high confidence in the findings and consistency across different geographic and resource settings. Carers expressed a clear desire for positive clinical and social outcomes for their infant. This was often reflected as a reframing of their priorities about what mattered, particularly in relation to their expectations for the infant. The main outcome hoped for was the infant’s survival, but families also hoped for good clinical outcomes such as weight gain. They also discussed their hopes for the child coming home and being a member of the family or community. Hopes for their child beyond childhood were rarely mentioned. Instead, parents focussed on short term outcomes, perhaps reflecting their need to stay focussed on the present.

### Active involvement in care

This analytical theme included four descriptive themes. The first related to how carers wanted to be taught, be involved in, and have confidence in their ability to deliver essential care (such as nappy changes, pain management, supporting nutrition) to their vulnerable infant. In addition, carers wanted to be supported to be involved in activities other than essential care which encourage opportunities for parenting, including bonding (for example, touching and cuddling), and social family activities (for example, singing or reading). Parents discussed the importance of kangaroo mother care, or skin to skin care, as both an opportunity to deliver care and as an opportunity for parenting. These themes were ubiquitous across all settings (with high confidence).

We had moderate confidence in the other descriptive themes in this group. Fathers wanted to be directly involved in the routine care of their infant, and to be supported to do this. However, they sometimes felt that they were not encouraged and not as welcome in the neonatal unit. It was not clear whether fathers had been asked for their views, or were willing and able to express these, in different locations and across different cultural groups. Carers also wanted support and processes to help them engage and take an active part in deciding what, and when, investigations, treatments, interventions and discharge occurs, although findings varied between settings with different cultural expectations and legal responsibilities.

### Coping at home

Carers wanted to be able to access support and advice urgently should they need it once the infant was discharged. Sometimes carers felt torn between the desire for freedom from continual observation and input from HCPs, and the pressure resulting from this autonomy as they became the primary carer for their child. Because of this conflict, and because this was not discussed in any low or middle-income country study, we had moderate confidence in this finding. The importance of wider family and community support (for practical arrangements and advice) was clear. Expertise from HCPs in the community was also valued, although professional support in the community was described as inconsistent.

Carers also wanted to be prepared for their infant’s discharge home. This included being taught how to look after their infant and gradually developing their experience in delivering practical care. Carers also discussed the value of preparation for discharge in emotional terms, such as the need to build their confidence after a long stay in a medical environment. Although fewer studies contributed to these descriptive themes, with less variability in geography and resources than with previous themes, we had high confidence in these findings due to high consistency between studies.

Lastly, many parents valued transition arrangements which they felt helped facilitate the safe transfer of care. This included the delivery of information, planned care pathways, and home visits, to ensure continuity of care. We had moderate confidence in this finding because the complexity and duration of transitional arrangements varied greatly between settings, and there were no studies from low or middle-income countries that contributed to this finding.

### Emotional support for families

In addition to practical support, emotional support from family members, particularly grandparents and siblings, was highly valued by parents and seen as essential both whilst the infant was in hospital and after discharge. Carers also wanted emotional support from HCPs, including reassurance and encouragement. Specialist counsellors were valued for their ability to provide emotional support, particularly when parents were faced with bereavement or a poor prognosis. Carers also valued support from other parents of preterm or sick infants who were considered helpful in providing information about the healthcare environment or specific morbidities, emotional support, and the provision of hope and comfort.

Fathers also valued emotional support, but often felt that they had to be emotionally strong to support the infant’s mother. We had moderate confidence in this finding, because there were only a few studies (especially from low and middle-income countries) that had explored the emotional support needs of fathers.

### Healthcare environment

Parents expressed strong views about the environments in which their infants were cared for, especially neonatal units. Carers wanted easy access to their infants. This included mechanisms or initiatives to help them to visit and interact with their infant and, where possible, the co-location of twins. Carers wanted to learn about the complex and sometimes frightening setting (in terms of noises, leads/equipment, and processes) in which they needed to live and care for their infant. They valued the ability to tour the NICU before the birth (where practical, for example, if a preterm or LBW birth was suspected in pregnancy). Carers were also concerned about real, or perceived, issues with staffing levels, equipment availability, and capability of the healthcare system to meet the needs of their infant.

Carers wanted privacy for breastfeeding or everyday family activities or interactions (for example, reading stories) in neonatal units. However, they also understood the need for medical observation, and therefore struggled with the conflict between their desire for seclusion and for close monitoring by staff. This was the descriptive theme for which we identified the most conflicting views, and it was not mentioned in any studies from low or middle-income countries. We therefore have low confidence in this finding.

### Information needs met

Carers described wanting a great deal of information relating to the infant’s condition, prognosis, investigations and procedures performed, as well as routine information about the infant’s daily events. However, they also felt overloaded. There was a tension between wanting to be told about all possible eventualities, and only wanting to know what they needed to know at any particular point in time. Due to this conflict, we had moderate confidence in this finding. Carers also wanted frequent and regular updates from the clinical team (rather than just meetings when sentinel events occur) and open channels of communication. We had moderate confidence in this finding due to a lack of evidence from low and middle-income countries.

Carers wanted HCPs to have good communication skills and to use a variety of information-giving methods. This related to a need for immediacy, an appropriate pace and timing of information-giving, and the opportunity for follow-up discussions. However, what constitutes good communication may vary by parent, culture, and situation. Although we had minor concerns that there was variability in describing what constituted good communication skills in different settings, we graded this finding with high confidence because all carers agreed that they wanted clear and appropriate communication from HCPs. Carers also wanted to feel confident in obtaining information from multiple sources including from HCPs (for example, by being comfortable or enabled to ask questions) and supplementing this information via other trusted sources such as leaflets, books and the internet.

### Logistical support

Carers wanted a range of practical and logistical support during the first two years of their infant’s life. Perhaps most immediately, they needed practical support to travel to see their infant, and support to sleep and live near them during their inpatient stay. In addition, they wanted support for the wider family (for example, crèche facilities for siblings) to allow them to engage in their ‘normal’ parenting and caring roles, before, and after, discharge. In all settings, carers wanted additional support and recognition of the direct (for example, medical costs) and indirect (for example, travel or lost earnings) financial burdens, both while the infant was an inpatient and afterwards. Finally, parents wanted formal, protected leave to allow them to visit the infant whilst in hospital, and to support them to provide care for a prolonged period after birth. Given the large variation in employment practices and laws across settings, this finding was graded with moderate confidence.

### Positive relationships with staff

Carers wanted to develop caring and sensitive relationships with the HCPs caring for their infant. They also wanted consistency in care and communication, so that they could better understand rules, advice, therapeutic plans and predictions of outcome, from and between, different HCPs. We have moderate confidence in this finding, as this was not discussed in studies from low and middle-income countries. Carers wanted HCPs to be well-trained, competent, and able to provide the specialist care needed by the infant. However, carers also desired the development of respectful, collaborative relationships with the HCP, where the parent was identified as an important part of the care-team with unique skills and knowledge of the infant, and that they were therefore heard and understood.

## DISCUSSION

### Main Findings

This systematic qualitative evidence synthesis of a contemporary evidence-base identified eight analytic themes and 31 descriptive themes. Nearly all of our descriptive themes were graded with high or moderate confidence. Families of preterm and LBW infants experience an extremely difficult time in the post-natal period. Whilst the ultimate desire for the majority of parents is a good outcome for their child, we found many other issues of importance to families related to processes within the health care environment. Unwelcoming hospital environments, poor logistical support, poor emotional support, non-collaborative relationships with staff, and lack of information all resulted in a limited ability for parents to engage in activities that matter most to them: to be actively involved in delivering care to, and parenting, their infant. The data also indicated that the attitudes and behaviours of HCPs are very important to families. This includes good communication and social skills and clinical competence as well as consistency between practitioners.

Previous research in this field has highlighted the state of liminality that parents of infants in NICU find themselves in, where parents feel like life is on hold, feel alienated, and struggle with their identity, sometimes not feeling like a real parent.[80 81] This can often lead parents to engaging in a deferential attitude towards the HCP ‘experts’ as they do not wish to destabilise the status quo for fear of damaging relationships. The results of this qualitative evidence synthesis enhance this picture, revealing that parents want support to be able to attend to the care of their infants and require positive relationships with staff from whom they feel fully informed about their infant’s progress.

### Strengths and weaknesses

We identified more than 200 studies where the views of carers of preterm, or LBW, infants had been investigated, across a range of healthcare and resource settings. Our sampling strategy allowed us to select 49 of these studies that represented a good geographical coverage, whilst retaining the studies with the highest methodological quality and with data that were rich and relevant to the research question.

Agreement in the views of parents appeared coherent across most of the evidence reviewed, but much of the work was performed when the infant was still receiving neonatal care, or about to be transferred to a community setting; little evidence of what mattered to carers in community-based care settings or in later years was identified. In addition, where there was less certainty in what was important to parents (including the ability to be involved in decisions about their infant’s care and the need for privacy), our lower confidence in these descriptive themes reflects the variation in how parents discussed these aspects of care, rather than the fact that the issues were not discussed at all.

Although we aimed to focus on what matters to families in the care of their preterm or LBW infant, we acknowledge that families’ views were sometimes influenced by their perception of the quality of the care that they had experienced. In addition, although we ensured our search included databases in which studies conducted in low resource countries might be listed (LILACS, African Journals Online) we elected to focus on papers published in English because this was the language of the study team. We did not translate studies from other languages because we were concerned about a loss of meaning and inaccuracies in our representation of the views of parents. However, we do acknowledge that this is a limitation, because papers published in other languages may have described what matters to parents differently.

### Implications for Research, Policy and Practice

Enabling a positive post-natal period for families of preterm and LBW infants is challenging. The focus of HCPs and the healthcare system is naturally on the medical needs and well-being of the infant and the parents’ practical and emotional needs are secondary to this. However, this review identifies a number of approaches that health care services and professionals can implement to improve the experience of parents. Many of these are consistent with the Family Centred Care model of healthcare which acknowledges the role of family members in supporting the well-being of the hospitalised infant, recognising the mutually beneficial partnerships among HCPs, parents and families.[12]

In this work, we have attempted to sample from a broad range of countries with varying resources, and from different healthcare settings. Given this, it is perhaps surprising how coherent the wishes of carers were across studies. However, practical barriers to implementation of care do affect families in different ways, and support needs to be bespoke between, and even within, different communities. Local services are therefore likely to be best placed to identify what support (for example, financial or logistic) might be most needed in working towards better support for families in this vulnerable period. One example of this may be the involvement of fathers in neonatal care, which is influenced by different cultural, social and legal barriers across different communities. Our conclusions in relation to fathers are graded with moderate, rather than high, confidence and further research is needed in low and middle-income countries to understand how best to involve and support fathers in those settings. We also found limited evidence on the views of extended family members. However, we hope that these data provide a framework for making decision that allows for the consideration of the views and needs of the wider family, especially as they were identified as important sources of support by parents.

Where we graded our findings with moderate or low confidence, this was often because the issues had not been reported in studies from low and middle-income countries. Preterm and LBW infants require high-quality inpatient care in dedicated facilities staffed by specialized HCPs, and this provision is known to be inadequate in many settings.[82] In the absence of appropriate infrastructure and expertise, it is not surprising that families are not able to participate in shared decision-making or express expectations about consistency of care, communication, and their need for privacy. Our data therefore supports calls to “*strengthen and invest*” in care for these infants, “*improve the quality of maternal and newborn care*” and “*harness the power of parents, families and communities*” in these settings.[11]

## CONCLUSION

We found high consistency in what matters to families, even though parents and family members reported a variety of experiences in the care of preterm or LBW infants. Local solutions may be best placed to address the needs of families. Most studies to date have been based in, or around, neonatal units. Further research is needed on what matters to parents who require community-based care at birth or after discharge and on the views and needs of fathers and extended family members, especially in low and middle-income countries.

## CONTRIBUTORSHIP STATEMENT

LH, DO and FW conceived and designed the study. MM designed the search strategy (in consultation with other review authors) and performed the database searches. LH, DO, HB, ED, TI and AA screened, selected and reviewed eligible reports. LH, DO, HB, ED and FW extracted data. LH, DO and FW completed the first draft of the paper. All authors commented on and revised the paper, and approved the final version. LH is the guarantor for the paper, and accepts full responsibility for the finished work and/or the conduct of the study, had access to the data, and controlled the decision to publish.

## Supporting information

Web Appendices

## Data Availability

No primary data were collected for this qualitative evidence synthesis. All data used are available in the original articles, manuscript, supporting information, and appendices.

## ACKNOWLEDGMENTS

We are grateful to Clive Gregory, Research Manager, for his support during the set-up of this project, and to Delyth Morris, Cardiff University Subject Librarian, and other Cardiff University library staff for their support in accessing the publications included in this review.

## ETHICAL APPROVAL

Ethical approval was not required for this qualitative evidence synthesis because no primary data were collected.

