## Supplementary material for "What matters to families about the healthcare of preterm or low birth weight infants: A qualitative evidence synthesis": Web Appendices

**Web Appendix 1. MEDLINE search (with results of scoping search conducted on May 19th 2021)**

- Key words and MeSH headings for qualitative research and values (combined using OR) highlighted in green
- Key words and MeSH headings for preterm and low birth weight babies (combined using OR) highlighted in orange
- Key words and MeSH headings for healthcare (combined using OR) highlighted in blue
- Final search, combining the three groups using AND and limiting to papers published in or after 2000, highlighted in pink

**Database: Ovid MEDLINE(R) ALL <1946 to May 19, 2021>**

--------------------------------------------------------------------------------

1 Qualitative Research/ (62667)

2 interviews as topic/ or focus groups/ or narration/ or Qualitative research/ (139784)

3 ((("semi-structured" or semistructured or unstructured or informal or "in-depth" or indepth or "face-to-face" or structured or guide) adj3 (interview* or discussion* or questionnaire*)) or (focus group* or qualitative or ethnograph* or fieldwork or "field work" or "key informant")).ti,ab. (374406)

4 (views or view or view point or experience* or perceive* or perception* or perspective* or satisfaction).tw. (2192306)

5 ((mother* or father* or families or family or parent* or carers) adj3 (value* or acceptable or important or matter*)).tw. (17243)

6 or/1-5 (2446590)

7 ((preterm or pre-term or premature or "low birth weight") adj3 (baby or babies or newborn or infant* or neonat*)).tw. (70177)

8 Infant, Low Birth Weight/ (19004)

9 Infant, Premature, Diseases/ (20849)

10 Infant, Premature/ (55559)

11 Infant, Extremely Low Birth Weight/ (1984)

12 Infant, Very Low Birth Weight/ (8814)

13 or/7-12 (110322)

14 Patient-Centered Care/ (20582)

| | |
|---|---|
| 15 | Aftercare/ (10008) |
| 16 | "Delivery of Health Care"/ (96387) |
| 17 | follow-up care.tw. (4990) |
| 18 | (care or health care or healthcare).tw. (1641511) |
| 19 | or/14-18 (1694044) |
| 20 | 6 and 13 and 19 (3538) |
| 21 | limit 20 to yr="2000 - 2021" (2866) |

**Web Appendix 2. Data extraction form**

| | | | | |
|---|---|---|---|---|
| **REVIEWER DETAILS** | | | | |
| Date of Review: | | | Reviewer: | |
| Study ID: | | | Journal: | |
| Year of Publication: | | | First Author: | |
| Article title: | | | | |
| **STUDY DETAILS** | | | | |
| Sources of funding | | | | |
| Study design | | | | |
| Authors' objective(s) | | | | |
| **METHODS** | | | | |
| Country: | | | | |
| Participants: | | | | |
| Patient population: (Tick all that apply) | ☐ Preterm | ☐ Low birth weight | ☐ Other (specify) ---------------------------------------- |  |
| Gestational Range (weeks) | Lower Limit | | Upper limit | |
| Birthweight range (g) | Lower Limit | | Upper limit | |
| Setting of healthcare (Tick all that apply) | ☐ Community | ☐ Paediatric unit | ☐ Neonatal unit | ☐ Outpatient care |
|  | ☐ Combined (specify) ----------------------------------------- |  | ☐ Other (specify) ---------------------------------------- |  |
| Was the study focused on a specific intervention: | ☐ No ☐ Yes<br>If yes, specify: ______________________________________________ | | | |
| Age of child at data collection: | | | | |
| Inclusion/ exclusion criteria: | | | | |
| Method of recruitment: | | | | |
| Method of data collection: | | | | |
| Method of data analysis: | | | | |
| Research context: | | | | |
| Phenomena of interest: | | | | |
| Dates or duration of data collection: | | | | |

| RESULTS | |
|---|---|
| Total participants by type (families) (individual parents) | |
| Participant characteristics: | |
| Findings/ theme 1 | |
| Findings/ theme 2 | |
| Findings / theme 3 | |
| Findings / theme 4 | |
| Findings / theme 5 | |
| Findings / theme 6 | |
| Findings / theme 7 | |
| Authors' comments on strengths and weaknesses | |
| Any quantitative findings of relevance | ☐ Yes ☐ No |
| If yes, what was the question examined? | |
| If yes, what data was presented? | |
| If yes, provide summary of data: | |
| **SUMMARY** | |
| Authors' overall conclusions: | |
| **CASP CHECKLIST ITEMS** | |
| **Section A: Are the results valid?** | |
| Q1. Was there a clear statement of the aims of the research?<br>☐ Yes ☐ Can't tell ☐ No | |
| Comments: | |
| Q2. Is a qualitative methodology appropriate?<br>☐ Yes ☐ Can't tell ☐ No | |
| Comments: | |

| |
|---|
| Q3. Was the research design appropriate to address the aims of the research?<br>☐ Yes ☐ Can’t tell ☐ No |
| Comments: |
| Q4. Was the recruitment strategy appropriate to the aims of the research?<br>☐ Yes ☐ Can’t tell ☐ No |
| Comments: |
| Q5. Was the data collected in a way that addressed the research issue?<br>☐ Yes ☐ Can’t tell ☐ No |
| Comments: |
| Q6. Has the relationship between researcher and participants been adequately considered?<br>☐ Yes ☐ Can’t tell ☐ No |
| Comments: |
| **Section B: What are the results?** |
| Q7. Have ethical issues been taken into consideration?<br>☐ Yes ☐ Can’t tell ☐ No |
| Comments: |
| Q8. Was the data analysis sufficiently rigorous?<br>☐ Yes ☐ Can’t tell ☐ No |
| Comments: |
| Q9. Is there a clear statement of findings?<br>☐ Yes ☐ Can’t tell ☐ No |
| Comments: |
| **Section C: Will the results help locally?** |
| Q10. How valuable is the research?<br>☐ Yes ☐ Can’t tell ☐ No |
| Comments: |
| **Overall assessment of study quality** |
| ☐ High ☐ Moderate ☐ Low |

**Web Appendix 3: Criteria used in data richness score (from EPOC 2017)**

| Score | Measure | Example |
|---|---|---|
| 1 | Very little qualitative data presented that relate to the synthesis objective. Those findings that are presented are fairly descriptive | For example, a mixed methods study using open ended survey questions or a more detailed qualitative study where only part of the data relates to the synthesis objective |
| 2 | Some qualitative data presented that relate to the synthesis objective | For example, a limited number of qualitative findings from a mixed methods or qualitative study |
| 3 | A reasonable amount of qualitative data that relate to the synthesis objective | For example, a typical qualitative research article in health services journal |
| 4 | A good amount and depth of qualitative data that relate to the synthesis objective | For example, a qualitative research article in a social sciences journal with more context and setting descriptions |
| 5 | A large amount and depth of qualitative data that relate in depth to the synthesis objective | For example, from a detailed ethnography or a published qualitative article with the same objectives as the synthesis. |

**Web Appendix 4. Other studies eligible for inclusion, but not selected for analysis (n=153, reported in 157 papers)**

| Reference | Year | Country | Setting | CASP Score | Data richness | Comment on reason for not selecting for analysis for studies classified as GOOD and with data richness of 3+ |
|---|---|---|---|---|---|---|
| (Alderdice et al. 2018) | 2018 | UK | Other | G | 2 | |
| (Aliabadi et al. 2014) | 2014 | Iran | Hospital | A | 2 | |
| (Alinejad-Naeini, Peyrovi, and Shoghi 2020) | 2020 | Iran | Hospital | G | 2 | |
| (Arivabene and Tyrrell 2010) | 2010 | Brazil | Hospital and Community | Poor (P) | 3 | |
| (Axelin et al. 2010) | 2010 | Finland | Hospital | G | 3 | Main focus is on a specific intervention (facilitated tucking by parents for infant pain) |
| (Baum et al. 2012) | 2012 | Israel | Hospital | A | 2 | |
| (Bernaix et al. 2006) | 2006 | USA | Hospital | A | 2 | |
| (Bissell and Long 2003) | 2003 | UK | Hospital and Community | A | 2 | |
| (Black, Holditch-Davis, and Miles 2009) | 2009 | USA | Hospital and Community | P | 2 | |
| (Blomqvist et al. 2013) | 2013 | Sweden | Hospital | G | 3 | |
| (Boykova 2016) | 2016 | USA, Can, Aust, NZ | Hospital | G | 2 | |
| (Brazy et al. 2001) | 2001 | USA | Hospital | A | 2 | |
| (Brodsgaard et al. 2017) | 2017 | Denmark | Hospital | G | 2 | |
| (Brotherton et al. 2021) | 2021 | Gambia | Hospital and Community | G | 3 | Main focus is on a specific intervention (kangaroo mother care) |

| | | | | | | |
|---|---|---|---|---|---|---|
| (Brown et al. 2013) | 2013 | USA | Hospital and Community | G | 2 | |
| (Brown and Pickler 2013) | 2013 | USA | Hospital and Community | A | 3 | |
| (Bujold et al. 2018) | 2018 | Canada | Hospital | A | 2 | |
| (Cervantes, Feeley, and Lariviere 2011) | 2011 | Canada | Hospital | G | 3 | Main focus is on a specific intervention (supplemental oxygen) |
| (Cescutti-Butler, Hewitt-Taylor, and Hemingway 2020) | 2020 | UK | Hospital | A | 3 | |
| (Chang, Cheng, and Lin 2007) | 2007 | Taiwan | Hospital | G | 2 | |
| (Changiz and Namnabati 2021) | 2021 | Iran | Hospital and Community | P | 2 | |
| (Clarke et al. 2021) | 2021 | UK | Hospital | A | 3 | |
| (Dadkhahtehrani et al. 2018) | 2018 | Iran | Hospital | A | 2 | |
| (de Cássia de Jesus Melo, de Oliveira Souza, and de Paula 2014) | 2014 | Brazil | Hospital | A | 3 | |
| (de Souza et al. 2010) | 2010 | Brazil | Hospital and Community | G | 2 | |
| (den Boer et al. 2021) | 2021 | Netherlands | Hospital | G | 3 | Main focus is on a specific intervention (showing recordings of neonatal resuscitation to parents) |

| | | | | | | |
|---|---|---|---|---|---|---|
| (Donohue et al. 2009) | 2009 | USA | Hospital | G | 3 | Main focus is on hospital transfers; no consensus amongst parents; content rather than thematic analysis |
| (Dur et al. 2018) | 2018 | Austria | Hospital | G | 2 | |
| (Dusing, Murray, and Stern 2008) | 2008 | USA | Hospital | G | 3 | Main focus is on a specific intervention (motor development education in NICU) |
| (Dutra Brazão Lelis et al. 2018) | 2018 | Brazil | Hospital | A | 3 | |
| (Ericson and Palmer 2019) | 2019 | Sweden | Community | A | 2 | |
| (Espitia and Ocampo González 2013) | 2013 | Colombia | Hospital and Community | A | 2 | |
| (Esquerra-Zwiers et al. 2016) | 2016 | USA | Hospital | G | 3 | Main focus is on a specific intervention (pasteurisation of donor milk) |
| (Falck, Moorthy, and Hussey-Gardner 2016) | 2017 | USA | Hospital | G | 2 | |
| (Fegran, Helseth, and Fagermoen 2008) | 2008 | Norway | Hospital | G | 2 | |
| (Fernandez Medina et al. 2018) | 2018 | Spain | Hospital | G | 2 | |
| (Fernandez Medina et al. 2019) | 2019 | Spain | Hospital | G | 2 | |
| (Ferreira de Lima and de Azevedo Mazza 2019) | 2019 | Brazil | Hospital | A | 2 | |
| (Flacking et al. 2006) | 2006 | Sweden | Hospital | G | 2 | |
| (Flacking, Thomson, and Axelin 2016) | 2016 | Sweden, UK, Finland | Hospital | G | 2 | |

| | | | | | | |
|---|---|---|---|---|---|---|
| (Fowler et al. 2019) | 2019 | Australia | Hospital | G | 2 | |
| (Garne et al. 2016) | 2016 | Denmark | Community | A | 4 | |
| (Garne Holm et al. 2019) | 2019 | Denmark | Community | A | 3 | |
| (Gibbs, Boshoff, and Stanley 2016) | 2016 | UK | Hospital | A | 3 | |
| (Gomes et al. 2021) | 2021 | Brazil | Community | A | 3 | |
| (Gonçalves Vieira Fernandes and Batoca Silva 2015) | 2015 | Portugal | Hospital | A | 3 | |
| (Granero-Molina et al. 2019) | 2019 | Spain | Hospital and Community | G | 2 | |
| (Gravem et al. 2009) | 2009 | USA | Hospital | G | 3 | Main focus is on a specific intervention (infant exercise in NICU) |
| (Griffin and Pickler 2011) | 2011 | USA | Hospital and Community | G | 2 | |
| (Hagen, Iversen, and Svindseth 2016) | 2016 | Norway | Hospital | G | 2 | |
| (Hakstad, Obstfelder, and Oberg 2016) | 2016 | Norway | Community | A | 3 | |
| (Hall and Brinchmann 2009) | 2009 | Denmark | Hospital | A | 2 | |
| (Hall et al. 2013) | 2013 | Denmark | Hospital and Community | A | 2 | |
| (Hasselberg, Huus, and Golsater 2016) | 2016 | Tanzania | Hospital | G | 2 | |

| | | | | | |
|---|---|---|---|---|---|
| (Heermann, Wilson, and Wilhelm 2005) | 2005 | USA | Hospital | P | 2 |
| (Helth and Jarden 2013) | 2013 | Denmark | Hospital | G | 2 |
| (Heydarpour, Keshavarz, and Bakhtiari 2017) | 2016 | Iran | Hospital | G | 2 |
| (Holditch-Davis and Miles 2000) | 2000 | USA | Hospital | G | 2 |
| (Holdren, Fair, and Lehtonen 2019) | 2019 | Finland, USA | Hospital | G | 2 |
| (Hollywood and Hollywood 2011) | 2011 | Ireland | Hospital | A | 3 |
| (Hua et al. 2021) | 2021 | China | Hospital and Community | G | 2 |
| (Hugill et al. 2013) | 2013 | UK | Hospital | A | 2 |
| (Hurst 2001a, 2001b) | 2001 | USA | Hospital | A | 3 |
| (Hurst, Engebretson, and Mahoney 2013) | 2013 | USA | Hospital | A | 2 |
| (Hwang et al. 2020) | 2021 | USA | Community | G | 2 |
| (Hyun-Ju 2018) | 2018 | South Korea | Hospital and Community | G | 2 |
| (Ikonen, Paavilainen, and Kaunonen 2016) | 2016 | Finland | Hospital | G | 2 |
| (Ireland et al. 2019) | 2019 | Australia | Hospital and Community | G | 2 |
| (Jackson, Ternestedt, and Schollin 2003) | 2003 | Sweden | Hospital and Community | G | 2 |

| | | | | | | |
|---|---|---|---|---|---|---|
| (Jager et al. 2020) | 2020 | USA | Hospital | G | 2 | |
| (Jebessa et al. 2021) | 2021 | Ethiopia | Hospital and Community | G | 2 | |
| (Jerntorp, Sivberg, and Lundqvist) | 2020 | Sweden | Hospital and Community | A | 2 | |
| (Johnson 2007) | 2007 | USA | Hospital | A | 4 | |
| (Kambarami, Mutambirwa, and Maramba 2002) | 2002 | Zimbabwe | Hospital and Community | P | 3 | |
| (Kennedy, Oakland, and Brotherson 2000) | 2000 | USA | Hospital and Community | A | 2 | |
| (Kim, Garfield, and Lee 2015) | 2015 | USA | Hospital | P | 1 | |
| (Kim 2018) | 2018 | USA | Community | A | 2 | |
| (Kim 2020) | 2020 | USA | Community | A | 2 | |
| (Koliouli, Gaudron, and Raynaud 2016) | 2016 | France | Hospital | P | 2 | |
| (Lara and Kind 2014) | 2014 | Brazil | Hospital | A | 3 | |
| (Lee, Kang, and Ji 2019) | 2019 | South Korea | Community | G | 3 | Main focus is on a specific intervention (home visits to support parents of LBW babies) |
| (Lindberg, Axelsson, and Ohrling 2007) | 2007 | Sweden | Hospital | G | 3 | Other included Swedish studies and included studies of fathers have richer data |
| (Lindberg, Axelsson, and Öhrling 2008; Lindberg and Ohrling 2008) | 2008 | Sweden | Hospital | G | 2 | |
| (Lindberg, Axelsson, and Öhrling 2009) | 2009 | Sweden | Community | G | 3 | Main focus is on a specific intervention (video conferencing at home) |

| | | | | | | |
|---|---|---|---|---|---|---|
| (Lindberg 2013) | 2013 | Sweden | Community | G | 3 | Main focus is on a specific intervention (video conferencing at home) |
| (Logan and Dormire 2018) | 2018 | USA | Hospital | G | 2 | |
| (Lundqvist, Westas, and Hallstrom 2007) | 2007 | Sweden | Hospital | G | 2 | |
| (Ma et al. 2021) | 2021 | China | Hospital and Community | G | 2 | |
| (Maastrup et al. 2018) | 2018 | Denmark | Hospital | A | 4 | |
| (MacDonald 2007) | 2007 | Canada | Hospital | A | 3 | |
| (Machado Pieszak et al. 2017) | 2017 | Brazil | Hospital | A | 3 | |
| (Mai and Wagner 2005) | 2005 | Denmark | Community | P | 2 | |
| (Martel et al. 2016) | 2016 | Canada | Hospital | A | 2 | |
| (Martin et al. 2016) | 2016 | USA | Hospital | A | 2 | |
| (Mathias, Mianda, and Ginindza 2020) | 2020 | Malawi | Community | A | 3 | |
| (Mello et al. 2002) | 2002 | Brazil | Community | P | 2 | |
| (Miele et al. 2018) | 2018 | Brazil | Hospital | A | 2 | |
| (Munhak da Silva et al. 2020; Munkah de Silva et al. 2020) | 2020 | Brazil | Community | A | 3 | |
| (Nascimento et al. 2020) | 2020 | Brazil | Hospital | A | 2 | |
| (Nelson and Bedford 2016) | 2016 | USA | Hospital | G | 3 | Main focus is on a specific intervention (Newborn Individualized Care and Assessment Program) |

| | | | | | | |
|---|---|---|---|---|---|---|
| (Nicolaou et al. 2009) | 2009 | UK | Hospital and Community | G | 2 | |
| (Niela-Vilen et al. 2015) | 2015 | Finland | Hospital and Community | G | 2 | |
| (Ntswane-Lebang and Khoza 2010) | 2010 | South Africa | Hospital | G | 2 | |
| (Nylund et al. 2020) | 2020 | Sweden | Community | A | 2 | |
| (Nyqvist and Kylberg 2008) | 2008 | Sweden | Hospital | G | 2 | |
| (Øberg et al. 2019) | 2018 | Norway | Hospital | G | 3 | Main focus is on a specific intervention (a motor intervention in NICU) |
| (Ochandorena-Acha et al. 2020) | 2020 | Spain | Hospital and Community | G | 2 | |
| (Palmquist, Holdren, and Fair 2020) | 2020 | USA | Hospital | A | 2 | |
| (Parker et al. 2018) | 2018 | USA | Hospital | G | 2 | |
| (Parker et al. 2020) | 2020 | USA | Hospital | G | 2 | |
| (Pepper et al. 2012) | 2012 | USA | Hospital | A | 2 | |
| (Phillips-Pula et al. 2013) | 2013 | USA | Community | G | 2 | |
| (Pizur-Barnekow et al. 2020) | 2020 | USA | Hospital | A | 2 | |
| (Pohlman 2009) | 2009 | USA | Hospital | G | 2 | |
| (Provenzi et al. 2016) | 2016 | Italy | Hospital | A | 2 | |
| (Rafael-Gutierrez et al. 2020) | 2020 | Spain | Hospital | G | 2 | |

| | | | | | | |
|---|---|---|---|---|---|---|
| (Reddy and McInerney 2007) | 2007 | South Africa | Hospital | A | 3 | |
| (Rossman et al. 2017; Rossman, Greene, and Meier 2015) | 2015 / 2017 | USA | Hospital | G | 2 | |
| (Rowe, Gardner, and Gardner 2005) | 2005 | Australia | Hospital and Community | A | 3 | |
| (Sarapat et al. 2017) | 2017 | Thailand | Hospital | A | 3 | |
| (Schrauwen, Kommers, and Oetomo 2018) | 2018 | Netherlands | Hospital | A | 2 | |
| (Sisk et al. 2010) | 2010 | USA | Hospital | G | 1 | |
| (Smith et al. 2012) | 2012 | USA | Hospital | G | 2 | |
| (Soderbck and Erlandsson 2012) | 2012 | Mozambique | Hospital and Community | A | 4 | |
| (Sommer and Cook 2015) | 2015 | New Zealand | Hospital | G | 3 | Main focus is on hospital transfers |
| (Spinelli et al. 2016) | 2016 | Italy | Hospital | G | 2 | |
| (Stacey, Osborn, and Salkovskis 2015) | 2015 | UK | Hospital | G | 3 | Other UK studies have richer data |
| (Stevens, Gazza, and Pickler 2014) | 2014 | USA | Hospital | G | 2 | |
| (Steyn, Poggenpoel, and Myburgh 2017) | 2017 | South Africa | Hospital | G | 2 | |
| (Strauss, Avrech Bar, and Stanger 2019) | 2019 | Israel | Hospital | A | 3 | |
| (Stuart Bright et al. 2020) | 2020 | Canada | Community | G | 2 | |

| | | | | | | |
|---|---|---|---|---|---|---|
| (Tabarsy et al. 2020) | 2020 | Iran | Hospital and Community | G | 2 | |
| (Thapa et al. 2018) | 2018 | Nepal | Hospital and Community | G | 2 | |
| (Tomeleri da Fonseca Pinto et al. 2017) | 2018 | Brazil | Hospital | G | 3 | Other included Brazilian studies have richer data |
| (Toral-López et al. 2016) | 2016 | Spain | Community | A | 3 | |
| (Torkomian Joaquim et al. 2018) | 2018 | Brazil | Hospital | A | 2 | |
| (Turner, Winefield, and Chur-Hansen 2013) | 2013 | Australia | Hospital and Community | P | 3 | |
| (Tzu-Ying et al. 2009) | 2009 | Taiwan | Hospital and Community | G | 1 | |
| (Vaerland, Vevatne, and Brinchmann 2017) | 2017 | Norway | Hospital | G | 2 | |
| (Værland, Vevatne, and Brinchmann 2018) | 2018 | Norway | Hospital | G | 2 | |
| (van den Hoogen et al. 2021) | 2021 | Netherlands | Hospital | G | 2 | |
| (van der Pal et al. 2014) | 2014 | Netherlands | Hospital and Community | A | 4 | |
| (Waiswa et al. 2010) | 2010 | Uganda | Community | A | 3 | |
| (Wakely, Rae, and Cooper 2010) | 2010 | Australia | Hospital | G | 2 | |
| (Watson 2011) | 2011 | UK | Hospital | A | 2 | |

| | | | | | | |
|---|---|---|---|---|---|---|
| (Weis, Zoffmann, and Egerod 2015) | 2015 | Denmark | Hospital | G | 3 | Other included Danish studies have richer data |
| (Widding et al. 2020) | 2020 | Sweden | Hospital | G | 2 | |
| (Yang et al. 2017) | 2017 | Singapore | Hospital | G | 2 | |
| (Yang et al. 2019) | 2019 | China | Hospital | G | 2 | |
| (Yue et al. 2020) | 2020 | China | Hospital | A | 2 | |
| (Zamanzadeh et al. 2013) | 2013 | Iran | Hospital and Community | A | 3 | |
| (Zani, Pamplona Tonete, and de Lima Parada 2014) | 2014 | Brazil | Hospital | A | 3 | |
| (Zhang and Wang 2019) | 2019 | China | Hospital | A | 2 | |

**Web Appendix 5. References for eligible studies not selected for inclusion in the analysis**

Alderdice, F., P. Gargan, E. McCall, and L. Franck. 2018. "Online information for parents caring for their premature baby at home: A focus group study and systematic web search." *Health Expectations* 21(4): 741-51.

Aliabadi, F., M. Kamali, L. Borimnejad, M. Rassafiani, M. Rasti, N. Shafaroodi, F. Rafii, and R. Askary Kachoosangy. 2014. "Supporting-emotional needs of Iranian parents with premature infants admitted to Neonatal Intensive Care Units." *Medical Journal of the Islamic Republic of Iran* 28: 53.

Alinejad-Naeini, M., H. Peyrovi, and M. Shoghi. 2020. "Emotional disorganization: The prominent experience of Iranian mothers with preterm neonate: A qualitative study." *Health Care for Women International*: 1-21.

Arivabene, J. C. and M. A. R. Tyrrell. 2010. "Kangaroo Mother Method: mothers' experiences and contributions to nursing." *Revista Latino-Americana de Enfermagem (RLAE)* 18(2): 262-68.

Axelin, A., L. Lehtonen, T. Pelander, and S. Salantera. 2010. "Mothers' Different Styles of Involvement in Preterm Infant Pain Care." *Jognn-Journal of Obstetric Gynecologic and Neonatal Nursing* 39(4): 415-24.

Baum, N., Z. Weidberg, Y. Osher, and D. Kohelet. 2012. "No Longer Pregnant, Not Yet a Mother: Giving Birth Prematurely to a Very-Low-Birth-Weight Baby." *Qualitative Health Research* 22(5): 595-606.

Bernaix, L. W., C. A. Schmidt, P. A. Jamerson, L. Seiter, and J. Smith. 2006. "The NICU experience of lactation and its relationship to family management style." *MCN: The American Journal of Maternal Child Nursing* 31(2): 95-100.

Bissell, G. and T. Long. 2003. "From the neonatal unit to home: how do parents adapt to life at home with their baby?" *Journal of Neonatal Nursing* 9(1): 7-12.

Black, B. P., D. Holditch-Davis, and M. S. Miles. 2009. "Life Course Theory as a Framework to Examine Becoming a Mother of a Medically Fragile Preterm Infant." *Research in Nursing & Health* 32(1): 38-49.

Blomqvist, Y. T., L. Frolund, C. Rubertsson, and K. H. Nyqvist. 2013. "Provision of Kangaroo Mother Care: supportive factors and barriers perceived by parents." *Scandinavian Journal of Caring Sciences* 27(2): 345-53.

Boykova, M. 2016. "Life After Discharge: What Parents of Preterm Infants Say About Their Transition to Home." *Newborn & infant nursing reviews* 16(2): 58-65.

Brazy, J. E., B. M. H. Anderson, P. T. Becker, and M. Becker. 2001. "How parents of premature infants gather information and obtain support." *Neonatal Network* 20(2): 41-48.

Brodsgaard, A., T. Helth, B. L. Andersen, and M. Petersen. 2017. "Rallying the Troops How Sharing Knowledge With Grandparents Supports the Family of the Preterm Infant in Neonatal Intensive Care Unit." *Advances in Neonatal Care* 17(3): E1-E10.

Brotherton, H., M. Daly, P. Johm, B. Jarju, J. Schellenberg, L. Penn-Kekana, and J. E. Lawn. 2021. ""We All Join Hands": Perceptions of the Kangaroo Method Among Female Relatives of Newborns in The Gambia." *Qualitative Health Research* 31(4): 665-76.

Brown, L. F., J. Griffin, B. Reyna, and M. Lewis. 2013. "The development of a mother's internal working model of feeding." *Journal for Specialists in Pediatric Nursing* 18(1): 54-64.

Brown, L. F. and R. Pickler. 2013. "A guided feeding intervention for mothers of preterm infants: Two case studies." *Journal for Specialists in Pediatric Nursing* 18(2): 98-108.

Bujold, M., N. Feeley, A. Axelin, and C. Cinquino. 2018. "Expressing Human Milk in the NICU Coping Mechanisms and Challenges Shape the Complex Experience of Closeness and Separation." *Advances in Neonatal Care* 18(1): 38-48.
Cervantes, A. C., N. Feeley, and J. Lariviere. 2011. "The experience of mothers whose very low-birth-weight infant requires the delivery of supplemental oxygen in the neonatal intensive care unit." *Advances in Neonatal Care* 11(1): 54-61.
Cescutti-Butler, L., J. Hewitt-Taylor, and A. Hemingway. 2020. "Powerless responsibility: A feminist study of women's experiences of caring for their late preterm babies." *Women and Birth* 33(4): E400-E08.
Chang, C., S. Cheng, and L. Lin. 2007. "Mothers' experiences of caring for premature children without major physical impairment in Taiwan." *Asian Journal of Nursing* 10(2): 121-28.
Changiz, T. and M. Namnabati. 2021. "Management of Comprehensive Care of multiple-birth infants from fetal to infancy period: challenges, training, strategies." *BMC Pediatrics* 21(1): 1-9.
Clarke, P., E. Allen, S. Atuona, and P. Cawley. 2021. "Delivery room cuddles for extremely preterm babies and parents: concept, practice, safety, parental feedback." *Acta Paediatrica* 110(5): 1439-49.
Dadkhahtehrani, T., N. Eskandari, Z. Khalajinia, and H. Ahmari-Tehran. 2018. "Experiences of Fathers with Inpatient Premature Neonates: Phenomenological Interpretative Analysis." *Iranian Journal of Nursing and Midwifery Research* 23(1): 71-78.
de Cássia de Jesus Melo, R., Í. E. de Oliveira Souza, and C. C. de Paula. 2014. "The voice of the woman-mother of a premature baby in the neonatal unit: a phenomenological approach." *Online Brazilian Journal of Nursing* 13(2): 194-202.
de Souza, N. L., A. C. Pinheiro-Fernandes, C. Clara-Costa Ido, B. Cruz-Enders, J. B. de Carvalho, and L. da Silva Mde. 2010. "Domestic maternal experience with preterm newborn children." *Revista de Salud Publica* 12(3): 356-67.
den Boer, M. C., M. Houtlosser, R. Witlox, R. van der Stap, M. C. de Vries, E. Lopriore, and A. B. Te Pas. 2021. "Reviewing recordings of neonatal resuscitation with parents." *Archives of Disease in Childhood Fetal & Neonatal Edition* 106(4): 346-51.
Donohue, P. K., B. Hussey-Gardner, L. J. Sulpar, R. Fox, and S. W. Aucott. 2009. "Parents' perception of the back-transport of very-low-birth-weight infants to community hospitals." *Journal of Perinatology* 29(8): 575-81.
Dur, M., V. Bruckner, C. Oberleitner-Leeb, R. Fuiko, B. Matter, and A. Berger. 2018. "Clinical relevance of activities meaningful to parents of preterm infants with very low birth weight: A focus group study." *Plos One* 13(8).
Dusing, S. C., T. Murray, and M. Stern. 2008. "Parent preferences for motor development education in the neonatal intensive care unit." *Pediatric Physical Therapy* 20(4): 363-8.
Dutra Brazão Lelis, B., M. Isicawa de Sousa, D. Faleiros de Mello, M. Wernet, A. B. Ferreira Velozo, and A. Moraes Leite. 2018. "MATERNAL RECEPTION IN THE CONTEXT OF PREMATURITY." *Journal of Nursing UFPE / Revista de Enfermagem UFPE* 12(6): 1563-69.
Ericson, J. and L. Palmer. 2019. "Mothers of preterm infants' experiences of breastfeeding support in the first 12months after birth: A qualitative study." *Birth-Issues in Perinatal Care* 46(1): 129-36.
Espitia, E. C. and M. P. Ocampo González. 2013. "Retrieving the offspring and caring for it during the first night at home." *Investigacion & Educacion en Enfermeria* 31(3): 354-63.

Esquerra-Zwiers, A., B. Rossman, P. Meier, J. Engstrom, J. Janes, and A. Patel. 2016. “"It's Somebody Else's Milk": Unraveling the Tension in Mothers of Preterm Infants Who Provide Consent for Pasteurized Donor Human Milk.” *Journal of Human Lactation* 32(1): 95-102.
Falck, A. J., S. Moorthy, and B. Hussey-Gardner. 2016. “Perceptions of Palliative Care in the NICU.” *Advances in Neonatal Care* 16(3): 191-200.
Fegran, L., S. Helseth, and M. S. Fagermoen. 2008. “A comparison of mothers' and fathers' experiences of the attachment process in a neonatal intensive care unit.” *Journal of Clinical Nursing* 17(6): 810-16.
Fernandez Medina, I. M., C. Fernandez-Sola, M. M. Lopez-Rodriguez, J. M. Hernandez-Padilla, M. D. M. Jimenez Lasserrotte, and J. Granero-Molina. 2019. “Barriers to Providing Mother's Own Milk to Extremely Preterm Infants in the NICU.” *Advances in Neonatal Care* 19(5): 349-60.
Fernandez Medina, I. M., J. Granero-Molina, C. Fernandez-Sola, J. M. Hernandez-Padilla, M. Camacho Avila, and M. D. M. Lopez Rodriguez. 2018. “Bonding in neonatal intensive care units: Experiences of extremely preterm infants' mothers.” *Women & Birth: Journal of the Australian College of Midwives* 31(4): 325-30.
Ferreira de Lima, V. and V. de Azevedo Mazza. 2019. “INFORMATION NEEDS OF FAMILIES ON THE HEALTH/DISEASE OF PRETERM INFANTS IN A NEONATAL INTENSIVE CARE UNIT.” *Texto & Contexto Enfermagem* 28: 1-17.
Flacking, R., U. Ewald, K. H. Nyqvist, and B. Starrin. 2006. “Trustful bonds: A key to "becoming a mother" and to reciprocal breastfeeding. Stories of mothers of very preterm infants at a neonatal unit.” *Social Science & Medicine* 62(1): 70-80.
Flacking, R., G. Thomson, and A. Axelin. 2016. “Pathways to emotional closeness in neonatal units - a cross-national qualitative study.” *Bmc Pregnancy and Childbirth* 16.
Fowler, C., J. Green, D. Elliott, J. Petty, and L. Whiting. 2019. “The forgotten mothers of extremely preterm babies: A qualitative study.” *Journal of Clinical Nursing* 28(11-12): 2124-34.
Garne Holm, K., A. Brodsgaard, G. Zachariassen, A. C. Smith, and J. Clemensen. 2019. “Parent perspectives of neonatal tele-homecare: A qualitative study.” *Journal of Telemedicine & Telecare* 25(4): 221-29.
Garne, K., A. Brodsgaard, G. Zachariassen, and J. Clemensen. 2016. “Telemedicine in Neonatal Home Care: Identifying Parental Needs Through Participatory Design.” *JMIR Research Protocols* 5(3): e100.
Gibbs, D. P., K. Boshoff, and M. J. Stanley. 2016. “The acquisition of parenting occupations in neonatal intensive care: A preliminary perspective.” *Canadian Journal of Occupational Therapy-Revue Canadienne D Ergotherapie* 83(2): 91-102.
Gomes, M. P., S. B. Saraty, A. A. Pereira, A. T. Parente, M. E. Santana, M. Cruz, and A. D. M. Figueira. 2021. “Mothers' knowledge of premature newborn care and application of Kangaroo Mother Care at home.” *Revista Brasileira de Enfermagem* 74(6): e20200717.
Gonçalves Vieira Fernandes, N. and E. M. Batoca Silva. 2015. “Parents' experience during the hospitalisation of the preterm infant.” *Revista de Enfermagem Referência* 4(4): 107-15.
Granero-Molina, J., I. M. Fernandez Medina, C. Fernandez-Sola, J. M. Hernandez-Padilla, M. D. M. Jimenez Lasserrotte, and M. D. M. Lopez Rodriguez. 2019. “Experiences of Mothers of Extremely Preterm Infants after Hospital Discharge.” *Journal of pediatric nursing* 45: e2-e8.
Gravem, D., K. D. Lakes, L. Teran, J. Rich, D. Cooper, and E. Olshansky. 2009. “Maternal perceptions of infant exercise in the neonatal intensive care unit.” *JOGNN: Journal of Obstetric, Gynecologic & Neonatal Nursing* 38(5): 527-33.

Griffin, J. B. and R. H. Pickler. 2011. “Hospital-to-Home Transition of Mothers of Preterm Infants.” *Mcn-the American Journal of Maternal-Child Nursing* 36(4): 252-57.
Hagen, I. H., V. C. Iversen, and M. F. Svindseth. 2016. “Differences and similarities between mothers and fathers of premature children: a qualitative study of parents' coping experiences in a neonatal intensive care unit.” *BMC Pediatrics* 16.
Hakstad, R. B., A. Obstfelder, and G. K. Oberg. 2016. “Parents' Perceptions of Primary Health Care Physiotherapy With Preterm Infants: Normalization, Clarity, and Trust.” *Qualitative Health Research* 26(10): 1341-50.
Hall, E. O. and B. S. Brinchmann. 2009. “Mothers of preterm infants: Experiences of space, tone and transfer in the neonatal care unit.” *Journal of Neonatal Nursing* 15(4): 129-36.
Hall, E. O. C., H. Kronborg, H. Aagaard, and B. S. Brinchmann. 2013. “The journey towards motherhood after a very preterm birth: Mothers' experiences in hospital and after home-coming.” *Journal of Neonatal Nursing* 19(3): 109-13.
Hasselberg, M., K. Huus, and M. Golsater. 2016. “Breastfeeding Preterm Infants at a Neonatal Care Unit in Rural Tanzania.” *Jognn-Journal of Obstetric Gynecologic and Neonatal Nursing* 45(6): 825-35.
Heermann, J. A., M. E. Wilson, and P. A. Wilhelm. 2005. “Mothers in the NICU: outsider to partner.” *Pediatric Nursing* 31(3): 176-86.
Helth, T. D. and M. Jarden. 2013. “Fathers' experiences with the skin-to-skin method in NICU: Competent parenthood and redefined gender roles.” *Journal of Neonatal Nursing* 19(3): 114-21.
Heydarpour, S., Z. Keshavarz, and M. Bakhtiari. 2017. “Factors affecting adaptation to the role of motherhood in mothers of preterm infants admitted to the neonatal intensive care unit: a qualitative study.” *Journal of Advanced Nursing* 73(1): 138-48.
Holditch-Davis, D. and M. S. Miles. 2000. “Mothers' stories about their experiences in the neonatal intensive care unit.” *Neonatal Network* 19(3): 13-21.
Holdren, S., C. Fair, and L. Lehtonen. 2019. “A qualitative cross-cultural analysis of NICU care culture and infant feeding in Finland and the US.” *Bmc Pregnancy and Childbirth* 19(1).
Hollywood, M. and E. Hollywood. 2011. “The lived experiences of fathers of a premature baby on a neonatal intensive care unit.” *Journal of Neonatal Nursing* 17(1): 32-40.
Hua, W. Z., L. Y. Wang, C. X. Li, J. M. Simoni, W. C. Yuwen, and L. P. Jiang. 2021. “Understanding preparation for preterm infant discharge from parents' and healthcare providers' perspectives: Challenges and opportunities.” *Journal of Advanced Nursing* 77(3): 1379-90.
Hugill, K., G. Letherby, T. Reid, and T. Lavender. 2013. “Experiences of Fathers Shortly After the Birth of Their Preterm Infants.” *Jognn-Journal of Obstetric Gynecologic and Neonatal Nursing* 42(6): 655-63.
Hurst, I. 2001a. “Mothers' strategies to meet their needs in the newborn intensive care nursery.” *Journal of Perinatal & Neonatal Nursing* 15(2): 65-82.
Hurst, I. 2001b. “Vigilant watching over: mothers' actions to safeguard their premature babies in the newborn intensive care nursery.” *Journal of Perinatal & Neonatal Nursing* 15(3): 39-57.
Hurst, N., J. Engebretson, and J. S. Mahoney. 2013. “Providing Mother's Own Milk in the Context of the NICU: A Paradoxical Experience.” *Journal of Human Lactation* 29(3): 366-73.
Hwang, S. S., M. G. Parker, B. N. Colvin, E. S. Forbes, K. Brown, and E. R. Colson. 2020. “Understanding the barriers and facilitators to safe infant sleep for mothers of preterm infants.” *Journal of Perinatology* 07: 07.

Hyun-Ju, K. 2018. “Experiences of Mothers of Premature Infants Receiving Rehabilitation Therapy.” *Child Health Nursing Research* 24(3): 298-309.
Ikonen, R., E. Paavilainen, and M. Kaunonen. 2016. “Trying to Live With Pumping: Expressing Milk for Preterm or Small for Gestational Age Infants.” *MCN: The American Journal of Maternal Child Nursing* 41(2): 110-15.
Ireland, S., R. A. Ray, S. Larkins, and L. Woodward. 2019. “Perspectives of time: a qualitative study of the experiences of parents of critically ill newborns in the neonatal nursery in North Queensland interviewed several years after the admission.” *BMJ Open* 9(5).
Jackson, K., B. Ternestedt, and J. Schollin. 2003. “From alienation to familiarity: experiences of mothers and fathers of preterm infants.” *Journal of Advanced Nursing (Wiley-Blackwell)* 43(2): 120-29.
Jager, S., K. Kavanaugh, S. Hoffman, T. Laitano, E. Jeffries, and B. Tucker Edmonds. 2020. “Parents' Descriptions of Neonatal Palliation as a Treatment Option Prior to Periviable Delivery.” *Journal of Perinatal & Neonatal Nursing* 34(2): 178-85.
Jebessa, S., J. A. Litch, K. Senturia, T. Hailu, A. Kahsay, K. A. Kuti, E. Wolka, A. M. Teklu, and W. Gezahegn. 2021. “Qualitative Assessment of the Quality of Care for Preterm, Low Birth Weight, and Sick Newborns in Ethiopia.” *Health Services Insights*: 1-13.
Jerntorp, S. H., B. Sivberg, and P. Lundqvist. “Fathers' lived experiences of caring for their preterm infant at the neonatal unit and in neonatal home care after the introduction of a parental support programme: A phenomenological study.” *Scandinavian Journal of Caring Sciences*.
Johnson, A. N. 2007. “The maternal experience of kangaroo holding.” *JOGNN: Journal of Obstetric, Gynecologic & Neonatal Nursing* 36(6): 568-73.
Kambarami, R. A., J. Mutambirwa, and P. P. Maramba. 2002. “Caregivers' perceptions and experiences of 'kangaroo care' in a developing country.” *Tropical Doctor* 32(3): 131-3.
Kennedy, T. S., M. J. Oakland, and M. J. Brotherson. 2000. “Making feeding decisions for preterm low birth weight infants: a family systems approach.” *Topics in Clinical Nutrition* 15(2): 38-84.
Kim, H. N. 2018. “Social Support Provision: Perspective of Fathers With Preterm Infants.” *Journal of Pediatric Nursing-Nursing Care of Children & Families* 39: 44-48.
Kim, H. N. 2020. “Information and Communications Technology and Education Customized for Fathers of Preterm Infants.” *Neonatal Network* 39(2): 66-74.
Kim, H. N., C. Garfield, and Y. S. Lee. 2015. “Paternal and Maternal Information and Communication Technology Usage as Their Very Low Birth Weight Infants Transition Home From the NICU.” *International Journal of Human-Computer Interaction* 31(1): 44-54.
Koliouli, F., C. Z. Gaudron, and J.-P. Raynaud. 2016. “Life experiences of French premature fathers: A qualitative study.” *Journal of Neonatal Nursing* 22(5): 244-49.
Lara, K. L. and L. Kind. 2014. “Subjectification processes experienced by mothers in a neonatology unit.” *Psicologia em Estudo* 19(4): 575-85.
Lee, J., J. C. Kang, and E. S. Ji. 2019. “Experiences of Mothers' Attachment in a Follow-Up Program Using Early Intervention for Low-Birth-Weight Infants.” *Asian Nursing Research* 13(3): 177-83.
Lindberg, B. 2013. “Access to videoconferencing in providing support to parents of preterm infants: Ascertaining parental views.” *Journal of Neonatal Nursing* 19(5): 259-65.
Lindberg, B., K. Axelsson, and K. Ohrling. 2007. “The birth of premature infants: Experiences from the fathers' perspective.” *Journal of Neonatal Nursing* 13(4): 142-49.

Lindberg, B., K. Axelsson, and K. Öhrling. 2008. “Adjusting to being a father to an infant born prematurely: experiences from Swedish fathers.” *Scandinavian Journal of Caring Sciences* 22(1): 79-85.
Lindberg, B., K. Axelsson, and K. Öhrling. 2009. “Taking care of their baby at home but with nursing staff as support: the use of videoconferencing in providing neonatal support to parents of preterm infants.” *Journal of Neonatal Nursing* 15(2): 47-55.
Lindberg, B. and K. Ohrling. 2008. “EXPERIENCES OF HAVING A PREMATURELY BORN INFANT FROM THE PERSPECTIVE OF MOTHERS IN NORTHERN SWEDEN.” *International Journal of Circumpolar Health* 67(5): 461-71.
Logan, R. M. and S. Dormire. 2018. “Finding My Way A Phenomenology of Fathering in the NICU.” *Advances in Neonatal Care* 18(2): 154-62.
Lundqvist, P., L. H. Westas, and I. Hallstrom. 2007. “From distance toward proximity: fathers lived experience of caring for their preterm infants.” *Journal of pediatric nursing* 22(6): 490-7.
Ma, R. H., Q. Zhang, Z. H. Ni, and H. T. Lv. 2021. “Transitional care experiences of caregivers of preterm infants hospitalized in a neonatal intensive care unit: A qualitative descriptive study.” *Nursing Open* 05: 05.
Maastrup, R., J. Weis, A. B. Engsig, K. L. Johannsen, and V. Zoffmann. 2018. “"Now she has become my daughter': parents' early experiences of skin-to-skin contact with extremely preterm infants.” *Scandinavian Journal of Caring Sciences* 32(2): 545-53.
MacDonald, M. 2007. “Mothers of pre-term infants in neonate intensive care.” *Early Child Development and Care* 177(8): 821-38.
Machado Pieszak, G., A. Moreira Paust, G. Calcagno Gomes, A. Moreira Arrué, E. Tatsch Neves, and L. Martins Machado. 2017. “Hospitalization of premature infants: parents' perceptions and revelations about nursing care.” *Rev Rene* 18(5): 591-97.
Mai, D. and L. Wagner. 2005. “'Home Early Program' -- experiences of parents to premature infants' one year after discharge.” *Nordic Journal of Nursing Research & Clinical Studies / Vård i Norden* 25(1): 60-63.
Martel, M. J., I. Milette, L. Bell, D. S. C. Tribble, and A. Payot. 2016. “Establishment of the Relationship Between Fathers and Premature Infants in Neonatal Units.” *Advances in Neonatal Care* 16(5): 390-98.
Martin, A. E., J. A. D'Agostino, M. Passarella, and S. A. Lorch. 2016. “Racial differences in parental satisfaction with neonatal intensive care unit nursing care.” *Journal of Perinatology* 36(11): 1001-07.
Mathias, C. T., S. Mianda, and T. G. Ginindza. 2020. “Facilitating factors and barriers to accessibility and utilization of kangaroo mother care service among parents of low birth weight infants in Mangochi District, Malawi: a qualitative study.” *BMC Pediatrics* 20(1).
Mello, D. F., S. M. Rocha, C. G. Scochi, and R. A. Lima. 2002. “Brazilian mothers' experiences of home care for their low birth weight infants.” *Neonatal Network - Journal of Neonatal Nursing* 21(1): 30-4.
Miele, M. J. O., R. C. Pacagnella, M. J. D. Osis, C. R. Angelini, J. L. Souza, and J. G. Cecatti. 2018. “"Babies born early?" - silences about prematurity and their consequences.” *Reproductive Health* 15.
Munhak da Silva, R. M., A. Zilly, E. R. dos Santos Nonose, L. M. Monti Fonseca, and D. Falleiros de Mello. 2020. “Care opportunities for premature infants: home visits and telephone support.” *Revista Latino-Americana de Enfermagem (RLAE)* 28: 1-8.

Munkah de Silva, A. Zilly, A. P. C. Toninato, L. Pancieri, M. C. C. Furtado, and D. F. Mello. 2020. “The vulnerabilities of premature children: home and institutional contexts.” *Revista Brasileira de Enfermagem* 73(suppl 4): e20190218.
Nascimento, A., A. C. Morais, R. D. C. Amorim, and D. V. D. Santos. 2020. “The care provided by the family to the premature newborn: analysis under Leininger's Transcultural Theory.” *Revista Brasileira de Enfermagem* 73(suppl 4): e20190644.
Nelson, A. M. and P. J. Bedford. 2016. “Mothering a Preterm Infant Receiving NIDCAP Care in a Level III Newborn Intensive Care Unit.” *Journal of Pediatric Nursing-Nursing Care of Children & Families* 31(4): E271-E82.
Nicolaou, M., R. Rosewell, N. Marlow, and C. Glazebrook. 2009. “Mothers' experiences of interacting with their premature infants.” *Journal of Reproductive & Infant Psychology* 27(2): 182-94.
Niela-Vilen, H., A. Axelin, H. L. Melender, and S. Salantera. 2015. “Aiming to be a breastfeeding mother in a neonatal intensive care unit and at home: a thematic analysis of peer-support group discussion in social media.” *Maternal and Child Nutrition* 11(4): 712-26.
Ntswane-Lebang, M. A. and S. Khoza. 2010. “MOTHERS' EXPERIENCES OF CARING FOR VERY LOW BIRTH WEIGHT PREMATURE INFANTS IN ONE PUBLIC HOSPITAL IN JOHANNESBURG, SOUTH AFRICA.” *Africa Journal of Nursing & Midwifery* 12(2): 69-82.
Nylund, A. G., M. G. Lindh, F. Ahlsson, and Y. T. Blomqvist. 2020. “Parents experiences of feeding their extremely preterm children during the first 2-3 years - A qualitative study.” *Acta Paediatrica* 109(5): 976-81.
Nyqvist, K. H. and E. Kylberg. 2008. “Application of the Baby Friendly Hospital Initiative to neonatal care: Suggestions by Swedish mothers of very preterm infants.” *Journal of Human Lactation* 24(3): 252-62.
Øberg, G. K., T. Ustad, L. Jørgensen, P. I. Kaaresen, C. Labori, and G. L. Girolami. 2019. “Parents' perceptions of administering a motor intervention with their preterm infant in the NICU.” *European Journal of Physiotherapy* 21(3): 134-41.
Ochandorena-Acha, M., R. Noell-Boix, M. Yildirim, M. Cazorla-Sanchez, M. Iriondo-Sanz, M. J. Troyano-Martos, and J. C. Casas-Baroy. 2020. “Experiences and coping strategies of preterm infants' parents and parental competences after early physiotherapy intervention: qualitative study.” *Physiotherapy Theory & Practice*: 1-14.
Palmquist, A. E., S. M. Holdren, and C. D. Fair. 2020. “"It was all taken away": Lactation, embodiment, and resistance among mothers caring for their very-low-birth-weight infants in the neonatal intensive care unit.” *Social Science & Medicine Vol 244 2020, ArtID 112648* 244.
Parker, M. G., S. S. Hwang, E. S. Forbes, B. N. Colvin, K. R. Brown, and E. R. Colson. 2020. “Use of the Theory of Planned Behavior Framework to Understand Breastfeeding Decision-Making Among Mothers of Preterm Infants.” *Breastfeeding Medicine* 15(10): 608-15.
Parker, M. G., A. M. Lopera, N. S. Kalluri, and C. J. Kistin. 2018. “"I Felt Like I Was a Part of Trying to Keep My Baby Alive": Perspectives of Hispanic and Non-Hispanic Black Mothers in Providing Milk for Their Very Preterm Infants.” *Breastfeeding Medicine* 13(10): 657-65.
Pepper, D., G. Rempel, W. Austin, C. Ceci, and L. Hendson. 2012. “More Than Information A Qualitative Study of Parents' Perspectives on Neonatal Intensive Care at the Extremes of Prematurity.” *Advances in Neonatal Care* 12(5): 303-09.
Phillips-Pula, L., R. Pickler, J. M. McGrath, L. F. Brown, and S. C. Dusing. 2013. “Caring for a Preterm Infant at Home A Mother's Perspective.” *Journal of Perinatal & Neonatal Nursing* 27(4): 335-44.

Pizur-Barnekow, K., U. O. Kim, S. I. Ahamed, M. K. K. Hasan, S. Dreier, S. R. Leuthner, N. Rau, and M. A. Basir. 2020. "Giving Voice to Parents in the Development of the Preemie Prep for Parents (P3) Mobile App." *Advances in Neonatal Care* 20(1): E9-E16.
Pohlman, S. 2009. "Fathering premature infants and the technological imperative of the neonatal intensive care unit: an interpretive inquiry." *Advances in Nursing Science* 32(3): E1-17.
Provenzi, L., S. Barello, M. Fumagalli, G. Graffigna, I. Sirgiovanni, M. Savarese, and R. Montirosso. 2016. "A Comparison of Maternal and Paternal Experiences of Becoming Parents of a Very Preterm Infant." *Jognn-Journal of Obstetric Gynecologic and Neonatal Nursing* 45(4): 528-41.
Rafael-Gutierrez, S. S., P. E. Garcia, A. S. Prellezo, L. R. Pauli, B. L. Del-Castillo, and R. B. Sanchez. 2020. "Emotional support for parents with premature children admitted to a neonatal intensive care unit: a qualitative phenomenological study." *Turkish Journal of Pediatrics* 62(3): 436-49.
Reddy, J. and P. A. McInerney. 2007. "The experiences of mothers who were implementing Kangaroo Mother Care (KMC) at a Regional Hospital in KwaZulu- Natal." *Curationis* 30(3): 62-67.
Rossman, B., M. M. Greene, A. L. Kratovil, and P. P. Meier. 2017. "Resilience in Mothers of Very-Low-Birth-Weight Infants Hospitalized in the NICU." *Jognn-Journal of Obstetric Gynecologic and Neonatal Nursing* 46(3): 434-45.
Rossman, B., M. M. Greene, and P. P. Meier. 2015. "The Role of Peer Support in the Development of Maternal Identity for "NICU Moms"." *Jognn-Journal of Obstetric Gynecologic and Neonatal Nursing* 44(1): 3-16.
Rowe, J. A., G. E. Gardner, and A. Gardner. 2005. "Parenting a preterm infant: experiences in a regional neonatal health services programme." *Neonatal, Paediatric & Child Health Nursing* 8(1): 18-24.
Sarapat, P., W. Fongkaew, U. Jintrawet, J. Mesukko, and L. Ray. 2017. "Perceptions and Practices of Parents in Caring for their Hospitalized Preterm Infants." *Pacific Rim International Journal of Nursing Research* 21(3): 220-33.
Schrauwen, L., D. R. Kommers, and S. B. Oetomo. 2018. "Viewpoints of Parents and Nurses on How to Design Products to Enhance Parent-Infant Bonding at Neonatal Intensive Care Units: A Qualitative Study Based on Existing Designs." *Herd-Health Environments Research & Design Journal* 11(2): 20-31.
Sisk, P., S. Quandt, N. Parson, and J. Tucker. 2010. "Breast Milk Expression and Maintenance in Mothers of Very Low Birth Weight Infants: Supports and Barriers." *Journal of Human Lactation* 26(4): 368-75.
Smith, V. C., G. K. Steelfisher, C. Salhi, and L. Y. Shen. 2012. "Coping with the neonatal intensive care unit experience: parents' strategies and views of staff support." *Journal of Perinatal & Neonatal Nursing* 26(4): 343-52.
Soderbck, M. and K. Erlandsson. 2012. "Kangaroo care in a Mozambican perinatal ward: A clinical case study." *African Journal of Midwifery & Women's Health* 6(1): 21-27.
Sommer, C. M. and C. M. Cook. 2015. "Disrupted bonds - parental perceptions of regionalised transfer of very preterm infants: a small-scale study." *Contemporary Nurse* 50(2-3): 256-66.
Spinelli, M., A. Frigerio, L. Montali, M. Fasolo, M. S. Spada, and G. Mangili. 2016. "'I still have difficulties feeling like a mother': The transition to motherhood of preterm infants mothers." *Psychology & Health* 31(2): 184-204.

Stacey, S., M. Osborn, and P. Salkovskis. 2015. “Life is a rollercoaster...What helps parents cope with the neonatal intensive care unit (NICU)?” *Journal of Neonatal Nursing* 21(4): 136-41.
Stevens, E. E., E. Gazza, and R. Pickler. 2014. “Parental Experience Learning to Feed Their Preterm Infants.” *Advances in Neonatal Care* 14(5): 354-61.
Steyn, E., M. Poggenpoel, and C. Myburgh. 2017. “Lived experiences of parents of premature babies in the intensive care unit in a private hospital in Johannesburg, South Africa.” *Curationis* 40(1): 1-8.
Strauss, Z., M. Avrech Bar, and V. Stanger. 2019. “Fatherhood of a Premature Infant: "A Rough Roller-Coaster Ride".” *Journal of Family Issues* 40(8): 982-1000.
Stuart Bright, K., C. Mannion, D. White, and S. R. Bouchal. 2020. “Transitioning Into the Role of Mother Following the Birth of a Very Low-Birth-Weight Infant: A Grounded Theory Pilot Study.” *Journal of Perinatal & Neonatal Nursing* 34(2): 125-33.
Tabarsy, B., J. Mirlashari, A. Nikbakht Nasrabadi, S. Joolaee, and H. Brown. 2020. “The bittersweet experience of parents living with premature multi-birth new-borns.” *Early Child Development and Care*: No Pagination Specified.
Thapa, K., D. Mohan, E. Williams, C. Rai, S. Bista, S. Mishra, and P. K. Hamal. 2018. “Feasibility assessment of an ergonomic baby wrap for kangaroo mother care: a mixed methods study from Nepal.” *Plos One* 13(11): e0207206.
Tomeleri da Fonseca Pinto, K. R., E. D. Gabriel Pinhatti, A. Valongo Zani, and C. M. Garcia de Lima Pàrada. 2017. “Maternal feelings about the hospitalization of the premature child: content analysis.” *Online Brazilian Journal of Nursing* 16(4): 9-9.
Toral-López, I. M. D., M. P. Fernández-Alcántara, P. P. González-Carrión, F. P. Cruz-Quintana, A. M. D. Rivas-Campos, and N. P. Pérez-Marfil. 2016. “Needs Perceived by Parents of Preterm Infants: Integrating Care Into the Early Discharge Process.” *Journal of pediatric nursing* 31(2).
Torkomian Joaquim, R. H. V., M. Wernet, A. Moraes Leite, L. M. Monti Fonseca, and D. Falleiros de Mello. 2018. “Early interactions between mothers and hospitalized premature babies: the focus on the essential needs of the child.” *Brazilian Journal of Occupational Therapy / Cadernos Brasileiros de Terapia Ocupacional* 26(3): 580-89.
Turner, M., H. Winefield, and A. Chur-Hansen. 2013. “The Emotional Experiences and Supports for Parents With Babies in a Neonatal Nursery.” *Advances in Neonatal Care* 13(6): 438-46.
Tzu-Ying, L., L. Hung-Ru, H. Tsu-Hsueh, H. Chyong-Hsin, and R. Bartlett. 2009. “Assuring the integrity of the family: being the father of a very low birth weight infant.” *Journal of Clinical Nursing (Wiley-Blackwell)* 18(4): 512-19.
Værland, I. E., K. Vevatne, and B. S. Brinchmann. 2018. “Mothers' experiences of having a premature infant due to pre-eclampsia.” *Scandinavian Journal of Caring Sciences* 32(2): 527-34.
Vaerland, I. E., K. Vevatne, and B. S. Brinchmann. 2017. “Fathers' experience of starting family life with an infant born prematurely due to mothers' severe illness.” *Sexual & Reproductive Healthcare* 13: 8-13.
van den Hoogen, A., R. Eijsermans, H. D. L. Ockhuijsen, F. Jenken, S. M. O. Maatman, M. J. Jongmans, L. Verhage, J. van der Net, and J. M. Latour. 2021. “Parents' experiences of VOICE: A novel support programme in the NICU.” *Nursing in Critical Care* 26(3): 201-08.

van der Pal, S. M., L. L. Alpay, G. J. van Steenbrugge, and S. B. Detmar. 2014. "An Exploration of Parents' Experiences and Empowerment in the Care for Preterm Born Children." *Journal of Child and Family Studies* 23(6): 1081-89.
Waiswa, P., S. Nyanzi, S. Namusoko-Kalungi, S. Peterson, G. Tomson, and G. W. Pariyo. 2010. "'I never thought that this baby would survive; I thought that it would die any time': perceptions and care for preterm babies in eastern Uganda." *Tropical Medicine & International Health* 15(10): 1140-47.
Wakely, L. T., K. Rae, and R. Cooper. 2010. "Stoic survival: the journey of parenting a premature infant in the bush." *Rural & Remote Health* 10(3): 1-10.
Watson, G. 2011. "Parental liminality: a way of understanding the early experiences of parents who have a very preterm infant." *Journal of Clinical Nursing* 20(9-10): 1462-71.
Weis, J., V. Zoffmann, and I. Egerod. 2015. "Enhancing person-centred communication in NICU: a comparative thematic analysis." *Nursing in Critical Care* 20(6): 287-98.
Widding, U., B. Hagglof, M. Adamsson, and A. Farooqi. 2020. "Parents of extremely and moderately preterm children reported long-lasting impressions of medical care and the hospital environment." *Acta Paediatrica* 109(9): 1772-77.
Yang, Y. Y., D. Brandon, H. Lu, and X. M. Cong. 2019. "Breastfeeding experiences and perspectives on support among Chinese mothers separated from their hospitalized preterm infants: a qualitative study." *International Breastfeeding Journal* 14(1).
Yang, Y. Y., H. G. He, S. Y. Lee, E. Holroyd, S. Shorey, and S. S. L. Koh. 2017. "Perceptions of Parents With Preterm Infants Hospitalized in Singaporean Neonatal Intensive Care Unit." *Journal of Perinatal & Neonatal Nursing* 31(3): 263-73.
Yue, J., J. Liu, S. Williams, B. Zhang, Y. Zhao, Q. Zhang, L. Zhang, X. Liu, S. Wall, G. Wetzel, G. Zhao, and J. Bouey. 2020. "Barriers and facilitators of kangaroo mother care adoption in five Chinese hospitals: a qualitative study." *BMC public health* 20(1): 1-11.
Zamanzadeh, V., M. Namnabati, L. Valizadeh, and Z. Badiee. 2013. "Mothers' Experiences of Infants Discharge in Iranian NICU Culture A Qualitative Study." *Advances in Neonatal Care* 13(4): E1-E7.
Zani, A. V., V. L. Pamplona Tonete, and C. M. G. de Lima Parada. 2014. "Maternal representations about the provision of care to newborns at risk: a collective discourse." *Online Brazilian Journal of Nursing* 13(3): 321-31.
Zhang, X. and J. Wang. 2019. "Massage intervention for preterm infants by their mothers: a randomized controlled trial." *Journal for Specialists in Pediatric Nursing* 24(2): e12238.
