## Supplementary Tables for "What matters to families about the healthcare of preterm or low birth weight infants: A qualitative evidence synthesis"

**Supplementary Table 1: PRISMA checklist**

| Section and Topic | Item # | Checklist item | Location where item is reported |
|---|---|---|---|
| **TITLE** | | | |
| Title | 1 | Identify the report as a systematic review/qualitative evidence synthesis. | Page 1 and 3 (title and abstract) |
| **ABSTRACT** | | | |
| Abstract | 2 | See the PRISMA 2020 for Abstracts checklist. | Page 3-4 |
| **INTRODUCTION** | | | |
| Rationale | 3 | Describe the rationale for the review in the context of existing knowledge. | Page 6 |
| Objectives | 4 | Provide an explicit statement of the objective(s) or question(s) the review addresses. | Page 6 |
| **METHODS** | | | |
| Eligibility criteria | 5 | Specify the inclusion and exclusion criteria for the review and how studies were grouped for the syntheses. | Page 7-9 |
| Information sources | 6 | Specify all databases, registers, websites, organisations, reference lists and other sources searched or consulted to identify studies. Specify the date when each source was last searched or consulted. | Page 10 |
| Search strategy | 7 | Present the full search strategies for all databases, registers and websites, including any filters and limits used. | Page 10 MEDLINE search in Appendix 1 |
| Selection process | 8 | Specify the methods used to decide whether a study met the inclusion criteria of the review, including how many reviewers screened each record and each report retrieved, whether they worked independently, and if applicable, details of automation tools used in the process. | Page 11 |
| Data collection process | 9 | Specify the methods used to collect data from reports, including how many reviewers collected data from each report, whether they worked independently, any processes for obtaining or confirming data from study investigators, and if applicable, details of automation tools used in the process. | Page 11 |
| Data items | 10a | List and define all outcomes for which data were sought. Specify whether all results that were compatible with each outcome domain in each study were sought (e.g. for all measures, time points, analyses), and if not, the methods used to decide which results to collect. | Table 1 |
| | 10b | List and define all other variables for which data were sought (e.g. participant and intervention characteristics, funding sources). Describe any assumptions made about any missing or unclear information. | Page 11 |
| Study risk of bias assessment | 11 | Specify the methods used to assess risk of bias in the included studies, including details of the tool(s) used, how many reviewers assessed each study and whether they worked independently, and if applicable, details of automation tools used in the process. | Page 12 |
| Effect measures | 12 | Specify for each outcome the effect measure(s) (e.g. risk ratio, mean difference) used in the synthesis or presentation of results. | N/A for a QES |

| Section and Topic | Item # | Checklist item | Location where item is reported |
|---|---|---|---|
| Synthesis methods | 13a | Describe the processes used to decide which studies were eligible for each synthesis (e.g. tabulating the study intervention characteristics and comparing against the planned groups for each synthesis (item #5)). | Study sampling described on page 12 |
| | 13b | Describe any methods required to prepare the data for presentation or synthesis, such as handling of missing summary statistics, or data conversions. | N/A for a QES |
| | 13c | Describe any methods used to tabulate or visually display results of individual studies and syntheses. | N/A for a QES |
| | 13d | Describe any methods used to synthesize results and provide a rationale for the choice(s). If meta-analysis was performed, describe the model(s), method(s) to identify the presence and extent of statistical heterogeneity, and software package(s) used. | Page 12 |
| | 13e | Describe any methods used to explore possible causes of heterogeneity among study results (e.g. subgroup analysis, meta-regression). | N/A for a QES |
| | 13f | Describe any sensitivity analyses conducted to assess robustness of the synthesized results. | N/A for a QES |
| Reporting bias assessment | 14 | Describe any methods used to assess risk of bias due to missing results in a synthesis (arising from reporting biases). | N/A for a QES |
| Certainty assessment | 15 | Describe any methods used to assess certainty (or confidence) in the body of evidence for an outcome. | Page 13 |
| **RESULTS** | | | |
| Study selection | 16a | Describe the results of the search and selection process, from the number of records identified in the search to the number of studies included in the review, ideally using a flow diagram. | Page 13-14 |
| | 16b | Cite studies that might appear to meet the inclusion criteria, but which were excluded, and explain why they were excluded. | PRISMA flow diagram (figure 1); web appendices 4 and 5 |
| Study characteristics | 17 | Cite each included study and present its characteristics. | Citations on page 13; Tables 2 and 3 |
| Risk of bias in | 18 | Present assessments of risk of bias for each included study. | Table 2 |

| Section and Topic | Item # | Checklist item | Location where item is reported |
|---|---|---|---|
| studies | | | |
| Results of individual studies | 19 | For all outcomes, present, for each study: (a) summary statistics for each group (where appropriate) and (b) an effect estimate and its precision (e.g. confidence/credible interval), ideally using structured tables or plots. | N/A for a QES |
| Results of syntheses | 20a | For each synthesis, briefly summarise the characteristics and risk of bias among contributing studies. | N/A for a QES |
| | 20b | Present results of all statistical syntheses conducted. If meta-analysis was done, present for each the summary estimate and its precision (e.g. confidence/credible interval) and measures of statistical heterogeneity. If comparing groups, describe the direction of the effect. | N/A for a QES |
| | 20c | Present results of all investigations of possible causes of heterogeneity among study results. | N/A for a QES |
| | 20d | Present results of all sensitivity analyses conducted to assess the robustness of the synthesized results. | N/A for a QES |
| Reporting biases | 21 | Present assessments of risk of bias due to missing results (arising from reporting biases) for each synthesis assessed. | N/A for a QES |
| Certainty of evidence | 22 | Present assessments of certainty (or confidence) in the body of evidence for each outcome assessed. | Table 4 |
| **DISCUSSION** | | | |
| Discussion | 23a | Provide a general interpretation of the results in the context of other evidence. | Pages 46, 48 |
| | 23b | Discuss any limitations of the evidence included in the review. | Pages 47 |
| | 23c | Discuss any limitations of the review processes used. | Pages 47-48 |
| | 23d | Discuss implications of the results for practice, policy, and future research. | Page 48 |
| **OTHER INFORMATION** | | | |
| Registration and protocol | 24a | Provide registration information for the review, including register name and registration number, or state that the review was not registered. | Page 7 |
| | 24b | Indicate where the review protocol can be accessed, or state that a protocol was not prepared. | Page 7 |
| | 24c | Describe and explain any amendments to information provided at registration or in the protocol. | Page 8-9 |
| Support | 25 | Describe sources of financial or non-financial support for the review, and the role of the funders or sponsors in the review. | Page 51 |
| Competing interests | 26 | Declare any competing interests of review authors. | Page 51 |
| Availability of data, code and other materials | 27 | Report which of the following are publicly available and where they can be found: template data collection forms; data extracted from included studies; data used for all analyses; analytic code; any other materials used in the review. | Data extraction form in web |

| Section and Topic | Item # | Checklist item | Location where item is reported |
|---|---|---|---|
| | | | appendix 2 |

**Supplementary Table 2. GRADE CERQual Evidence Profile (Analytical theme: A positive outcome for the child)**

| Analytical theme: A positive outcome for the child | Summary of review finding | Studies contributing to review finding | Methodological limitations | Coherence | Adequacy | Relevance | CERQual assessment | Explanation of CERQual assessment |
|---|---|---|---|---|---|---|---|---|
| | Positive outcome for the child | 20 studies | No or very minor concerns | No or very minor concerns | No or very minor concerns | No or very minor concerns<br>Good geographic and resource spread | High confidence | 20 studies with no or very minor methodological concerns, and coherence and adequacy of findings.<br>Consistent conclusions on the phenomenon of interest, and a good geographic and resource spread of studies. |

**Supplementary Table 3. GRADE CERQual Evidence Profile (Analytical theme: Active involvement in care)**

| Analytical theme: Active involvement in care | Summary of review finding | Studies contributing to review finding | Methodological limitations | Coherence | Adequacy | Relevance | CERQual assessment | Explanation of CERQual assessment |
|---|---|---|---|---|---|---|---|---|
| | **Delivering care** | 27 studies | No or very minor concerns | No or very minor concerns | No or very minor concerns | No or very minor concerns<br>Good geographic and resource spread | High confidence | 27 studies with no or very minor methodological concerns, and coherence and adequacy of findings.<br>Consistent conclusions on the phenomenon of interest, and a good geographic and resource spread of studies. |
| | **Fathers involved** | 11 studies | No or very minor concerns | No or very minor concerns | No or very minor concerns | Moderate concerns | Moderate confidence | 11 studies with no or very minor methodological concerns, and coherence and adequacy of findings.<br>Moderate concerns on the phenomenon, regarding whether fathers were willing and able to express their views and whether there was a breadth of studies in different locations and across different cultural groups |
| | **Opportunities for parenting** | 25 studies | No or very minor concerns | No or very minor concerns | No or very minor concerns | No or very minor concerns<br>Good geographic and resource spread | High confidence | 25 studies with no or very minor methodological concerns, and coherence and adequacy of findings.<br>Consistent conclusions on the phenomenon of interest, and a good geographic and resource spread of studies. |
| | **Shared decision making and consent** | 10 studies | No or very minor concerns | No or very minor concerns | No or very minor concerns | Moderate concerns | Moderate confidence | 10 studies with no or very minor methodological concerns, and coherence and adequacy of findings.<br>Moderate concerns on the relevance, regarding cultural expectations and legal responsibilities in different locations and across different cultural groups |

### Supplementary Table 4. GRADE CERQual Evidence Profile (Analytical theme: Coping at home)

| | Summary of review finding | Studies contributing to review finding | Methodological limitations | Coherence | Adequacy | Relevance | CERQual assessment | Explanation of CERQual assessment |
|---|---|---|---|---|---|---|---|---|
| **Analytical theme: Coping at home** | **Accessing support in a crisis** | 7 studies | No or very minor concerns | No or very minor concerns | No or very minor concerns | No or very minor concerns | High confidence | 7 studies with no or very minor methodological concerns, and coherence and adequacy of findings.<br>Consistent conclusions on the phenomenon of interest, and a good geographic and resource spread of studies. |
| | **Autonomy** | 6 studies | No or very minor concerns | Moderate concerns | No or very minor concerns | Moderate concerns<br>No lower resource studies | Moderate confidence | 6 studies with no or very minor methodological concerns, and adequacy of findings.<br>Moderate concerns for coherence on the reports of desire for genuine autonomy and the internal conflict that this brings.<br>No studies from lower resource settings. |
| | **Extended family support/ community resources** | 7 studies | No or very minor concerns | No or very minor concerns | No or very minor concerns | Minor concerns<br>Limited geographic and resource spread. | High confidence | 7 studies with no or very minor methodological concerns, and coherence and adequacy of findings.<br>Consistent conclusions on the phenomenon of interest<br>Only one study from a lower resource setting. |
| | **HCP expertise in the community** | 7 studies | No or very minor concerns | No or very minor concerns | No or very minor concerns | Minor concerns<br>Limited geographic and resource spread. | High confidence | 7 studies with no or very minor methodological concerns, and coherence and adequacy of findings.<br>Consistent conclusions on the phenomenon of interest<br>Only one study from a lower resource setting. |

| | | | | | | | | |
|---|---|---|---|---|---|---|---|---|
| | **Preparation for discharge** | 15 studies | No or very minor concerns | No or very minor concerns | No or very minor concerns | No or very minor concerns<br>Good geographic and resource spread | High confidence | 15 studies with no or very minor methodological concerns, and coherence and adequacy of findings.<br>Consistent conclusions on the phenomenon of interest, and a good geographic and resource spread of studies. |
| | **Transition arrangements** | 5 studies | No or very minor concerns | Moderate concerns | No or very minor concerns | Moderate concerns<br>No lower resource studies | Moderate confidence | 5 studies with no or very minor methodological concerns, and adequacy of findings.<br>Moderate concerns on coherence and on relevance particularly regarding the complexity and duration of transitional arrangements.<br>No lower resource setting studies. |

**Supplementary Table 5. GRADE CERQual Evidence Profile (Analytical theme: Emotional support for family)**

| | Summary of review finding | Studies contributing to review finding | Methodological limitations | Coherence | Adequacy | Relevance | CERQual assessment | Explanation of CERQual assessment |
|---|---|---|---|---|---|---|---|---|
| **Analytical theme: Emotional support for family** | **Support for and from the wider family** | 9 studies | No or very minor concerns | No or very minor concerns | No or very minor concerns | No or very minor concerns<br>Good geographic and resource spread | High confidence | 9 studies with no or very minor methodological concerns, and coherence and adequacy of findings.<br>Consistent conclusions on the phenomenon of interest, and a good geographic and resource spread of studies. |
| | **Support for parents** | 21 studies | No or very minor concerns | No or very minor concerns | No or very minor concerns | No or very minor concerns<br>Good geographic and resource spread | High confidence | 21 studies with no or very minor methodological concerns, and coherence and adequacy of findings.<br>Consistent conclusions on the phenomenon of interest, and a good geographic and resource spread of studies. |
| | **Support for fathers** | 7 studies | No or very minor concerns | No or very minor concerns | No or very minor concerns | Moderate concerns<br>No lower resource studies | Moderate confidence | 7 studies with no or very minor methodological concerns, and coherence and adequacy of findings.<br>Moderate concerns on the phenomenon, regarding whether fathers were willing and able to express their views on support and whether there was a breadth of studies in different locations and across different cultural groups |
| | **Support from other parents in similar situation** | 17 studies | No or very minor concerns | No or very minor concerns | No or very minor concerns | No or very minor concerns<br>Good geographic and resource spread | High confidence | 17 studies with no or very minor methodological concerns, and coherence and adequacy of findings.<br>Consistent conclusions on the phenomenon of interest, and a good geographic and resource spread of studies. |

**Supplementary Table 6. GRADE CERQual Evidence Profile (Analytical theme: Healthcare environment)**

| | Summary of review finding | Studies contributing to review finding | Methodological limitations | Coherence | Adequacy | Relevance | CERQual assessment | Explanation of CERQual assessment |
|---|---|---|---|---|---|---|---|---|
| **Analytical theme: Healthcare environment** | **Access to babies** | 15 studies | No or very minor concerns | No or very minor concerns | No or very minor concerns | No or very minor concerns<br>Good geographic and resource spread | High confidence | 15 studies with no or very minor methodological concerns, and coherence and adequacy of findings.<br>Consistent conclusions on the phenomenon of interest, and a good geographic and resource spread of studies. |
| | **Orientation and familiarity with NICU** | 19 studies | No or very minor concerns | No or very minor concerns | No or very minor concerns | No or very minor concerns<br>Good geographic and resource spread | High confidence | 19 studies with no or very minor methodological concerns, and coherence and adequacy of findings.<br>Consistent conclusions on the phenomenon of interest, and a good geographic and resource spread of studies. |
| | **Privacy vs monitoring** | 6 studies | No or very minor concerns | Moderate concerns | No or very minor concerns | Moderate concerns<br>No lower resource studies | Low confidence | 6 studies with no or very minor methodological concerns, and adequacy of findings.<br>Moderate concerns on coherence and on relevance with carers recognising the conflict between their need for privacy and the need for monitoring.<br>No lower resource setting studies. |
| | **Staffing and equipment levels** | 10 studies | No or very minor concerns | No or very minor concerns | No or very minor concerns | No or very minor concerns<br>Good geographic and resource spread | High confidence | 10 studies with no or very minor methodological concerns, and coherence and adequacy of findings.<br>Consistent conclusions on the phenomenon of interest, and a good geographic and resource spread of studies. |

**Supplementary Table 7. GRADE CERQual Evidence Profile (Analytical theme: Information needs met)**

| Analytical theme: Information needs met | Summary of review finding | Studies contributing to review finding | Methodological limitations | Coherence | Adequacy | Relevance | CERQual assessment | Explanation of CERQual assessment |
|---|---|---|---|---|---|---|---|---|
| | **Information about the baby** | 17 studies | No or very minor concerns | Moderate concerns | No or very minor concerns | No or very minor concerns<br>Good geographic and resource spread | Moderate confidence | 17 studies with no or very minor methodological concerns, and adequacy and relevance of findings.<br>A good geographic and resource spread of studies.<br>Moderate concerns on coherence because of tensions expressed by parents about wanting to know as much as possible and being overwhelmed with information |
| | **Frequent updates** | 7 studies | No or very minor concerns | No or very minor concerns | No or very minor concerns | Moderate concerns<br>No lower resource studies | Moderate confidence | 7 studies with no or very minor methodological concerns, and coherence and adequacy of findings.<br>Moderate concerns on relevance because of no lower resource setting studies. |
| | **How information is given** | 9 studies | No or very minor concerns | Minor concerns | No or very minor concerns | No or very minor concerns<br>Good geographic and resource spread | High confidence | 9 studies with no or very minor methodological concerns, and adequacy of findings.<br>Minor concerns on coherence due to variability in understanding of what good communication skills entail.<br>Consistent conclusions on the phenomenon of interest, and a good geographic and resource spread of studies. |
| | **Matching needs with information** | 18 studies | No or very minor concerns | No or very minor concerns | No or very minor concerns | No or very minor concerns<br>Good geographic and resource spread | High confidence | 18 studies with no or very minor methodological concerns, and adequacy of findings.<br>Minor concerns on coherence due to variability in understanding of what good communication skills entail. |

| | | | | | | | | |
|---|---|---|---|---|---|---|---|---|
| | | | | | | | | Consistent conclusions on the phenomenon of interest, and a good geographic and resource spread of studies. |

**Supplementary Table 8. GRADE CERQual Evidence Profile (Analytical theme: Logistical support)**

| Analytical theme: Logistical support | Summary of review finding | Studies contributing to review finding | Methodological limitations | Coherence | Adequacy | Relevance | CERQual assessment | Explanation of CERQual assessment |
|---|---|---|---|---|---|---|---|---|
| | **Accommodation (comfort and facilities)** | 13 studies | No or very minor concerns | No or very minor concerns | No or very minor concerns | No or very minor concerns<br>Only one lower resource study | High confidence | 13 studies with no or very minor methodological concerns, and coherence and adequacy of findings.<br>Consistent conclusions on the phenomenon of interest.<br>Only one lower resource setting study |
| | **Broader family support and impact** | 13 studies | No or very minor concerns | No or very minor concerns | No or very minor concerns | No or very minor concerns | High confidence | 13 studies with no or very minor methodological concerns, and coherence and adequacy of findings.<br>Consistent conclusions on the phenomenon of interest, and reasonable geographic and resource spread of studies. |
| | **Costs of treatment** | 12 studies | No or very minor concerns | No or very minor concerns | No or very minor concerns | No or very minor concerns | High confidence | 12 studies with no or very minor methodological concerns, and coherence and adequacy of findings.<br>Consistent conclusions on the phenomenon of interest, and reasonable geographic and resource spread of studies. |
| | **Parental leave** | 7 studies | No or very minor concerns | No or very minor concerns | No or very minor concerns | Moderate concerns<br>One lower resource study | Moderate confidence | 7 studies with no or very minor methodological concerns, and coherence and adequacy of findings.<br>Moderate concerns on relevance because there is only one lower resource setting study. |

**Supplementary Table 9. GRADE CERQual Evidence Profile (Analytical theme: Positive relationships with staff)**

| | Summary of review finding | Studies contributing to review finding | Methodological limitations | Coherence | Adequacy | Relevance | CERQual assessment | Explanation of CERQual assessment |
|---|---|---|---|---|---|---|---|---|
| **Analytical theme: Positive relationships with staff** | **Compassion and sensitivity** | 19 studies | No or very minor concerns | No or very minor concerns | No or very minor concerns | No or very minor concerns | High confidence | 19 studies with no or very minor methodological concerns, and coherence and adequacy of findings.<br>Consistent conclusions on the phenomenon of interest, and good geographic and resource spread of studies. |
| | **Consistency in care/communication** | 13 studies | No or very minor concerns | No or very minor concerns | No or very minor concerns | Moderate concerns<br>No lower resource study | Moderate confidence | 13 studies with no or very minor methodological concerns, and coherence and adequacy of findings.<br>Moderate concerns on the phenomenon because there was no study from a lower resource setting included. |
| | **HCP expertise/care** | 8 studies | No or very minor concerns | No or very minor concerns | No or very minor concerns | Minor concerns<br>Two lower resource studies | High confidence | 8 studies with no or very minor methodological concerns, and coherence and adequacy of findings.<br>Consistent conclusions on the phenomenon of interest.<br>Only two lower resource setting studies included. |
| | **Respect, collaboration and trust** | 14 studies | No or very minor concerns | No or very minor concerns | No or very minor concerns | Minor concerns<br>Two lower resource studies | High confidence | 14 studies with no or very minor methodological concerns, and coherence and adequacy of findings.<br>Consistent conclusions on the phenomenon of interest.<br>Only two lower resource setting studies included. |

### Supplementary Table 10. Synthesis results with additional supporting data for analytical theme: “A positive outcome for the child”

| Analytical theme: A positive outcome for the child | Descriptive theme (review finding) | Studies contributing to review finding | CERQual grading | Supporting data |
|---|---|---|---|---|
| | **A positive outcome for the child:** Carers expressed a desire for a successful clinical and social outcome for their baby as they grow (as distinct from the process or interventions themselves). | 20 studies<br><br>Arnold 2013<br>Chang Lee 2009<br>Dorner 2020<br>Gallegos-Martinez 2013<br>Granrud 2014<br>Hendriks 2017<br>Ignell Modé 2014<br>Leonard 2008<br>Lian 2020<br>Lomotey 2020<br>Lundqvist 2019<br>Ncube 2016<br>Neu 2020<br>Nyondo 2020<br>Orapiriyakul 2007<br>Petty 2018<br>Rossman 2011<br>Skene 2012<br>Veronez 2017<br>Villeneuve 2018 | High confidence | “When I had a preterm child... I changed...it changed everything that I used to think was important...I used to think my baby should be beautiful.. .with big eyes, but now I only want my baby to grow healthily...” Chang Lee et al 2009 (county = Taiwan, study setting = hospital)<br><br>“to tell you the truth, I don’t have no worry my baby going to talk or walk, whatever. As long as she’s in my presence and care and she’s still breathing and alive, I don’t care” Dorner et al 2020 (USA, hospital)<br><br>“He is going on four months here... what more could one want than for the child to be well and all, to have him at home like the others.” Gallegos-Martinez et al 2013 (Mexico, hospital)<br><br>“No one could promise us that it would be all right. I understand that they can't do that, but it was very hard not being able to obtain reassurance that things would go well.” Granrud et al 2014 (Norway, hospital)<br><br>“Weight gain for them is crucial ... that is the only way really, that one can see whether they are ... developing. Especially as a parent ... the doctors might be able to see through other things, but as a parent your only measure of how your child is doing is the influence of weight gain”. Leonard 2008 (South Africa, hospital)<br><br>“I know her weight every day, I’ve recorded it…I take a certain amount of comfort [in it], like if she’s putting on weight, she’s going in the right direction…I come out and look at the monitor. I’m checking the numbers. I’m kind of like that’s good!” Lian et al 2020 (Singapore, hospital)<br><br>“I am just happy having this baby regardless of how he is. Babies die daily on this unit so I'm happy whenever I go and meet my baby alive…. some people even have babies with abnormalities yet they are happy. How much more me? Once I |

| | | | | |
|---|---|---|---|---|
| | | | | see him alive, I become happy and I pray he continues to live." Lomotey et al 2020 (Ghana, hospital)<br><br>"What is frightening is that we are sleeping with other mothers who also have babies in the same room as mine. Sometimes when we come, maybe coming for a three o'clock feed or any other time, you will find that baby dead. That is what frightened me thinking that maybe one day, when I come here, I will be told that my baby is no longer alive." Ncube 2016 (Botswana, hospital)<br><br>"At first I was worried because my wife told me that the baby was very small and she doubted if the baby will survive, but later on after seeing other women going through the same and that their babies are surviving, I felt happy that my baby's life is going to be saved. You know, this is a good intervention…It's now three weeks since the baby was initiated on the kangaroo, the baby was so small but now it is growing, it is gaining weight and now I am happy…I know my baby could have easily died without this intervention but now I know there are very high chances that my baby will make it and we will be discharged from the hospital." Nyondo-Mipando et al 2020 (Malawi, hospital)<br><br>'It is horrible being scared to death… it is devastating them saying "He might have to have a shunt, he might have brain damage" all those things, horrible, horrible' Petty 2018 (UK, hospital and community) |

**Supplementary Table 11. Synthesis results with additional supporting data for analytical theme: "Active involvement in care"**

| Analytical theme: Active involvement in care | Descriptive theme (review finding) | Studies contributing to review finding | CERQual grading | Supporting data |
|---|---|---|---|---|
| | **Delivering care:** Carers want to be taught, be involved in, and have confidence in their ability to deliver some of the essential care (such as nappy changes, pain management, supporting nutrition) to their vulnerable infant | 27 studies<br><br>Abeasi 2020<br>Amorim 2019<br>Blomqvist 2012<br>Chang Lee 2009<br>Feeley 2013<br>Finlayson 2014<br>Gallegos-Martinez 2013<br>Hagi-Pederson 2021<br>Leonard 2008<br>Lian 2020<br>Liu 2019<br>Lomotey 2020<br>Lundqvist 2019<br>Mihae 2021<br>Ncube 2016<br>Neu 2020<br>Noren 2018<br>Orapiriyakul 2007<br>Premji 2017<br>Rossman 2011<br>Russell 2014<br>Skene 2012<br>Treherne 2017<br>Unsworth 2021<br>Veronez 2017<br>Villeneuve 2018<br>Yu 2020 | High confidence | "I was mortified when someone gave my child their first bath. I felt like saying, 'you shouldn't have done that . . . that's my baby'". Finlayson et al 2014 (UK, hospital)<br><br>"I would like them to tell me how to bathe him, what to feed him, if what I already did and is okay, or when he cries or when does something else (what to do), because I have already held him" Gallegos-Martinez et al 2013 (Mexico, hospital)<br><br>"About the third week there was less milk, the sisters encouraged me, by then she hadn't yet started breast-feeding ... but they encouraged me to keep putting her in the kangaroo care position and that really helped to stimulate the milk flow and when she eventually ...breastfed I had more than enough milk" Leonard 2008 (South Africa, hospital)<br><br>"What I appreciate about the nurses here [is that they] make you [become] autonomous [in] the good kind of way. [. . .] They don't force you into it. First, they show you how to do it, and if you're comfortable doing it, they'll supervise you, but then after a while, they'll let you go and fly on your own" Liu et al 2019 (Canada, hospital)<br><br>"I like the feeling when my baby is sucking more than when giving him expressed milk because it makes me feel l have really given birth." Lomotey et al 2020 (Ghana, hospital)<br><br>"In a way, like my mother… she taught me, told me how to care of the baby… you can fix these things… I think that's really necessary support for a mother of a premature infant." Mihae 2021 (South Korea, hospital)<br><br>"I was afraid of him … not knowing how I am going to handle him…. There is a nurse who told me not to be afraid of him because it is me who is |

| | | | | |
|---|---|---|---|---|
| | | | | going to take care of him while they show us how to take care of them" Ncube 2016 (Botswana, hospital)<br><br>"Obviously there's a lot of different nurses here, but they've been really good about helping teach you what to do and help you learn new things. They're going beyond just providing him care; they're helping us learn to be parents." Neu 2020 (USA, hospital)<br><br>"[Firstly] I didn't have courage to do anything... I had no skill to take care or even diapering him. I only touched and told him to get well soon, I had to come early in the morning because a nurse will allow me to hold my baby for a moment while cleaning an incubator, I wanted to take care for him by myself and give him my breast feeding...and take care for him with love after taking off the feeding tube." Orapiriyakul et al 2017 (Thailand, hospital)<br><br>"[The neonatal unit] are great about you being able to open the incubator and they get you very involved in looking after them, and changing nappies, and cleaning bottoms and so you're doing as much of the care as you can" Russell 2014 (UK, hospital)<br><br>"The nurse last night asked me, 'Can you change her nappy or do you want me to do it?' and I thought, I'm her bloody mother, of course I know how to change a nappy. Feeding is one thing because she is getting tube fed, but nappy changing, and cleaning and washing her down and stuff, that is my job." Skene 2012 (UK, hospital)<br><br>"[...] they are teaching me how to change her nappy. It was a great experience because I had never learned anything before. But I think I did it right." Veronez et al 2017 (Brazil, hospital)<br><br>"[. . .] that was my precious bit, that's what I could do for my baby, is get her dressed and try and feed her [. . .]" Villeneuve et al 2018 (UK, hospital) |
| | **Fathers involved:** Fathers want support to be directly involved in the routine care of their | 11 studies<br><br>Adama 2017 | Moderate confidence | "I went to the unit every evening after work to spend time with my child and wife but anytime I went there, I could not see and hold my baby for long. I felt like I was not welcome but I kept on asking questions. All the |

| | | | | |
|---|---|---|---|---|
| | infant, alongside support and encouragement to do this | Blomqvist 2012<br>Feeley 2013<br>Gallegos-Martinez 2013<br>Guillaume 2013<br>Leonard 2008<br>Lian 2020<br>Lundqvist 2019<br>Nyondo-Mipando 2020<br>Olsson 2017<br>Sawyer 2013 | | attention was given to the mother and I was left out.” Adama et al 2017 (Ghana, hospital)<br><br>‘Holding [his baby] is just out of this world’ Feeley et al 2013 (Canada, hospital)<br><br>“Kangaroo care has become so much part of me now. I am now owning this thing. I was thinking what was kangaroo care? It was nothing up in the sky, it was nothing other than what I was doing, all the other ideas, the breathing and the posture were part of it, and I developed it for myself so that it became more meaningful”. Leonard 2008 (South Africa, hospital)<br><br>“The more you visit, the better it is for you. Because it becomes more normal, you know what to expect and what not to expect.” Lian et al 2020 (Singapore, hospital) |
| | **Opportunities for parenting:** Carers want to have support with activities to encourage bonding (including touching and cuddling), and social family activity which falls outside the need for essential care. | 25 studies<br><br>Arnold 2013<br>Blomqvist 2012<br>Chang Lee 2009<br>Feeley 2013<br>Finlayson 2014<br>Glazer 2021<br>Granrud 2014<br>Guillaume 2013<br>Kim 2020<br>Klawetter 2019<br>Leonard 2008<br>Lian 2020<br>Liu 2019<br>Lomotey 2020<br>Lorie 2021<br>Mihae 2021<br>Ncube 2016<br>Neu 2020<br>Noren 2018 | High confidence | ‘‘I couldn’t verbalise the word ‘‘son” during the first days (...) I only started to internalise this from the moment I could touch him, starting to feel him (.. .) the first time they [nurses] put him in skin-to-skin contact with me it was the moment I felt: this is real, he is mine.” Amorim et al 2019 (Portugal, hospital)<br><br>“I felt much more comfortable to touch him when his condition got better...it made it much easier to interact with him, you know.. .the feeling of being afraid of hurting him eased...and I knew he was fine and there were always nurses around ...so.. .it was much easier...” Chang Lee et al 2009 (Taiwan, hospital)<br><br>“I asked the nurse if it was ok to have a cuddle and she said, “you should have been having at least one a day”, but at the start I didn’t realise you could ask them.” Finlayson 2014 (UK, hospital)<br><br>“I couldn’t touch her; well, far away on one side of the incubator because before I had to see her through glass; that is, not even near the incubator [...] Why do they do this? I don’t agree, it is offensive [...] I saw it as an obstacle [...] I am participating by bringing my milk” Gallegos-Martinez et al 2013 (Mexico, hospital) |

| | | | | |
|---|---|---|---|---|
| | | Olsson 2017<br>Orapiriyakul 2007<br>Skene 2012<br>Treherne 2017<br>Veronez 2017<br>Villeneuve 2018 | | "The hardest thing for me today, it's the kisses. The fact that I cannot kiss her, because the bond with her, it's all that: it's playing with all her senses, as much as possible, it's talking to her, touching her, being there, that she feels the love in our gestures" Guillaume et al 2013 (France, hospital)<br><br>"My first experience with kangaroo caring [for] her... when I took her out and put her against my skin, it was just ... a great sense of relief for the first time really I felt bonded with her, that it was my daughter. So I think that kangaroo care helps to bridge that initial ... gap that is between a mother and her preterm baby". Leonard 2008 (South Africa, hospital)<br><br>"I am happy whenever I place my baby in between my breast." Lomotey et al 2020 (Ghana, hospital)<br><br>"As I was performing kangaroo care, I felt like I was acting as a mother, and I felt proud that I was helping my baby and doing something for her." Mihae 2021 (South Korea, hospital)<br><br>"The moment she placed him on my chest, I felt this warm sense over my heart, and then I couldn't leave him, I just wanted to sit there all the time..." Noren 2018 (Sweden, hospital and community)<br><br>"It was the best. It was the feeling I ever had when he got to be with me" (STS). Olsson et al 2017 (Sweden, hospital)<br><br>"It's my job to make sure she's comfortable so after they've done what they need to, if I don't think she's comfortable, I'll calm her, turn her, give her a dummy (pacifier) and stroke her face until she's settled. I'll stay there until she's not moving around too much. Their job's not personal to her, because they do it with other babies, but mine's personal." Skene 2012 (UK, hospital)<br><br>"Today I got to hold my baby for the first time after her surgery. I am very happy about that [...] warm her with the warmth of my body" Veronez et al 2017 (Brazil, hospital) |

| | | | | |
|---|---|---|---|---|
| | **Shared decision making and consent:** Cares want support and processes to help them engage and take an active part in deciding what, and when, investigations, treatments, interventions and discharge occurs. | 10 studies<br><br>Amorim 2019<br>Finalyson 2014<br>Franck 2017<br>Hendriks and Abraham 2017<br>Liu 2019<br>Lorie 2021<br>Ncube 2016<br>Petty 2019b<br>Treherne 2017<br>Villeneuve 2018 | Moderate confidence | "I did not experience this moment as a freedom but rather as a responsibility of course because this baby cannot decide for herself. We are her parents and we should make this decision. And we should decide what is best for our baby. Now in retrospect, I regard that as a great act of love. But in those hours, I thought I would die. But you do not die and you go on and you have to decide." Hendriks 2017 (Switzerland, hospital)<br><br>"care plans would be modified without any explanation" Liu et al 2019 (Canada, hospital)<br><br>"Our voice is important... our views must be taken on board" Petty 2019b (UK, hospital and community) |

**Supplementary Table 12. Synthesis results with additional supporting data for analytical theme: "Coping at home"**

| | Descriptive theme (review finding) | Studies contributing to review finding | CERQual grading | Supporting data |
|---|---|---|---|---|
| Analytical theme: Coping at home | **Accessing support in a crisis:** Carers want mechanism to find help and advice urgently after discharge home when the primary care is transferred to the family | 7 studies<br><br>Adama 2017<br>Brødsgaard 2015<br>Dos Santos 2014<br>Franck 2017<br>Hägi-Pederson 2021a<br>Neu 2020<br>Unsworth 2021 | High confidence | "you don't really know who to call if something happens. I asked my wife wasn't given any number to call for help. When you're discharged, it's like you've to do everything but I don't know anything about preterm infants" Adama et al 2017 (Ghana, hospital)<br><br>"There were a few days that I was worried because the baby was already at home, and I called the women there and they treated me very well" dos Santos et al 2014 (Brazil, hospital)<br><br>"We had a video meeting with them that day. We talked to (the nurse) in there. She says, 'You have to come because... we have to do something about it...'" Hägi-Pedersen et al 2021 (Denmark, community)<br><br>"I think the biggest thing is just that worry of is everything going to be okay? Here he's on the monitor all of the time so you've got that safety blanket that if something goes wrong, a) you'll know about it and b) there's people here that jump right in and help with it. Because even now he still has times where his heart rate drops or his oxygen drops, and I know you (infant) have to go a certain amount of time without that happening before you can be discharged, but I'm a worrier by nature so it's the kind of what if? Because sometimes too, he'll be sleeping on me like this and I don't notice something is wrong until the alarms go off. So it's like when I'm home and don't have that am I going to miss something?" Neu 2020 (USA, hospital)<br><br>"Sometimes it is raining and when the baby is unwell, there is no transport to get there very fast to get help. It is very difficult and sometimes if the baby was to be helped there is no means of getting there faster." Unsworth et al 2021 (Kenya, community) |

| | | | | |
|---|---|---|---|---|
| | **Autonomy:** Carers want to take over the responsibility as the primary, and often only caregiver and decision maker for the infant after discharge. | 6 studies<br><br>Brødsgaard 2015<br>Hägi-Pederson 2021a<br>Leonard 2008<br>Lundqvist 2019<br>Petty 2018<br>Premji 2017 | Moderate confidence | "I felt divided [in NICU], torn into two pieces, which were only assembled once I got home." Brødsgaard et al 2015 (Denmark, hospital and community)<br><br>"At home I didn't have spectators … I felt at peace and I could hold her and put her on me and it was beautiful." Leonard 2006 (South Africa, hospital)<br><br>"Suddenly you are alone… it felt good to bring him home, of course that is what anyone wants, but it was nerve-racking" Petty 2018 (UK, hospital)<br><br>"I was definitely, definitely ready to be going home and I knew just being in my own house and just kind of get to our routine" Premji et al 2017 (Canada, community) |
| | **Extended family support/ community resources:** Carers want support in obtaining advice and care from the wider community, rather than the just from the health sector. | 7 studies<br><br>Amorim 2019<br>Brødsgaard 2015<br>Fernandez Medina 2021<br>Franck 2017<br>Hägi-Pederson 2021a<br>Unsworth 2021<br>Villeneuve 2018 | High confidence | "We were very lucky because we have a great family. My oldest daughter was 21 months old, I couldn't attend to her needs, and my mother-in-law had to step into a mother's role with her, but there are many parents who are alone…" Fernandez Medina et al 2021 (Spain, community and out-patients)<br><br>"Some tell you that those are curses from not following traditions, that maybe you did something during pregnancy" Unsworth et al 2021 (Kenya, community) |
| | **Healthcare professional (HCP) expertise in the community:** Carers want experienced, knowledgeable, and competent HCPs in the community to take over the health support for the ex-preterm and low birth weight infant. | 7 studies<br><br>Fernandez-Medina 2021<br>Franck 2017<br>Hägi-Pederson 2021<br>Jantsch 2021<br>Petty 2018 | High confidence | "Each professional gives you different advice, in the end you choose a professional and follow the steps. When I was in doubt, I always paid attention to the neonatologist, so I stopped taking him to the primary health centre" Fernandez Medina et al 2021 (Spain, community and out-patients)<br><br>"We've mixed a little bit of the two things we've been recommended... we've taken 50% from the neonatal ward and 50% |

| | | | | |
|---|---|---|---|---|
| | | Premji 2017<br>Unsworth 2021 | | from the municipal health visitor, and then we made our own mix of what we think fits" Hägi-Pedersen et al 2021 (Denmark, community)<br><br>"The only thing they tell you to do is treatment at a health center, consult with a pediatrician, always take them to see her, but I don't have it here in the neighborhood and nowhere." Jantsch 2021 (Brazil, community)<br><br>"I would like to see better parent support from GPs, from consultants, from health visitors, a better understanding" Petty 2018 (UK, hospital and community) |
| | **Preparation for discharge:** Carers want to be practicably prepared, with education and confidence in their increased delivery of care, alongside emotionally support, for the discharge from a healthcare setting to the home, often after a long stay in a medical environment. | 15 studies<br><br>Adama 2017<br>Brødsgaard 2015<br>Dos Santos 2014<br>Franck 2017<br>Hägi-Pederson 2021a<br>Kim 2020<br>Lorie 2021<br>Lundqvist 2019<br>Orapiriyakul 2007<br>Petty 2018<br>Premji 2017<br>Unsworth 2021<br>Veronez 2017<br>Wernet 2015<br>Yu 2020 | High confidence | "I wasn't told anything by the doctors or nurses, my wife just called me one day in a happy mood and said they have been discharged…no discharge education whatsoever, but I trust my wife may have been given some tips" Adama et al 2017 (Ghana, hospital)<br><br>"We learned everything we needed and knew what we had to do, I was quite comfortable when we went home." Brødsgaard et al 2015 (Denmark, hospital and community)<br><br>"They gave me a lot of strength ... so they guided me in everything, how to proceed, how to be careful with him ... and it was not for me getting so scared ... I was going to nail it.... and I'm achieving all that they went through with me" dos Santos et al 2014 (Brazil, hospital)<br><br>"I wish to receive education and training on the rehabilitation of preemies … Also, I'd like to learn how to deal with expected situations about my baby after leaving the NICU." Kim 2020 (South Korea, hospital)<br><br>"At home everything is different and I really felt insecure. I was not prepared at all and I did not have the entire team of doctors and nurses available anymore". Lorie et al 2021 (Netherlands, hospital) |

| | | | | |
|---|---|---|---|---|
| | | | | "Before you release the mum and baby, make sure they are in a proper pattern… I should have continued expressing and then I would have had more milk… there was no specialist support for feeding" Petty 2018 (UK, hospital and community)<br><br>"They give orders all the time and say a good mother is the one that provides good care. Then, they say 'Pay attention, learn it, then you'll do it alone.' Near the discharge, they give a brochure, a class, etc. I wanted them to see my pain. I tried that many times. They just said 'Provide good care and then you'll take the brochure at discharge.' But they don't allow us to provide care there (in the NICU), only two or three days before the discharge." Wernet et al 2015 (Brazil, hospital)<br><br>"When he was there in the hospital, they were in charge, but they said the mother had the obligation to provide care. I guess so, but there, it's impossible to know it. They are serious and the rules don't allow it. And then, you keep watching, watching, just watching. Then, you're afraid. I am afraid, I don't know if I provide proper care. I don't know if I provide good care. It's confusing." Wernet et al 2015 (Brazil, hospital) |
| | **Transition arrangements:** Carers want adequate and safe transfer of health care responsibilities to other community organisations and professionals as part of the discharge home. This includes the delivery of information, pathways of care, and home visits, in order to delivery safe continuity of care. | 5 studies<br><br>Dos Santos 2014<br>Fernandez Medina 2021<br>Franck 2017<br>Lundqvist 2019<br>Noren 2018 | Moderate confidence | "At home you talk more freely, you're there in your own place, the person will help you, he/she will see how your home and your life is... I think I would pick a thousand times only home rather than the hospital because at the hospital there are always a lot of people, everything is always very busy, you know? You can not keep asking things, there's no way that the person can stop caring for all children to pay attention to you. At home it is the opposite, they stop, they will be there just to talk to you, to see how the baby is... I think this is excellent, I think you could not take it from us" dos Santos et al 2014 (Brazil, hospital)<br><br>"My son came home with a nasogastric tube, there is a nurse in the hospital who is in charge of teaching you how the tube and the feeding pump work, but when you are at home the responsibility is entirely yours, the moment when the nasogastric tube goes outside |

| | | | | |
|---|---|---|---|---|
| | | | | is very complicated, and you can't be thinking as a parent whether you have put the tube in correctly or not, and if his lung is going to fill up with food… therefore, counting on a professional companion is very important". Fernandez Medina et al 2021 (Spain, hospital and community) |

**Supplementary Table 13. Synthesis results with additional supporting data for analytical theme: "Emotional support for families"**

| | Descriptive theme (review finding) | Studies contributing to review finding | CERQual grading | Supporting data |
|---|---|---|---|---|
| **Analytical theme: Emotional support for families** | **Support for and from the wider family:** Carers want emotional support from, and for the wider family (including grandparents and siblings) | 9 studies<br><br>Chang Lee 2009<br>Feeley 2013<br>Gallegos-Martinez 2013<br>Leonard 2008<br>Lian 2020<br>Nyondo-Mipando 2020<br>Orapiriyakul 2007<br>Premji 2017<br>Villeneuve 2018 | High confidence | "My mother helped me with everything...my parents-in-law were also great...they visited the baby at hospital regularly, and always asked me not to worry..." Chang Lee et al 2009 (Taiwan, hospital)<br><br>'I have my in-laws…they are always there, whether it be for moral or practical support' Feeley et al 2013 (Canada, hospital)<br><br>"I have a lot of responsibilities. I think the first one is to help the mother is doing the kangaroo mother care. When she wants to rest, the baby is put on me and I do exactly what she does except breastfeeding. The second thing is to see her doing everything accordingly as told by the doctors… Also, I think it is my responsibility to encourage her that things are going to be ok. " (Grandmother) Nyondo-Mipando et al 2020 (Malawi, hospital)<br><br>"I would say like just having family support was probably the most helpful thing once we were home" Premji et al 2017 (Canada, community) |
| | **Support for parents:** Parents want to have emotional support from any source (often healthcare workers); including reassurance and encouragement, to enhance their interactions and journey after the birth of a preterm or low birth weight infant. | 21 studies<br><br>Amorim 2019<br>Dorner 2020<br>Fernandez-Medina 2021<br>Franck 2017<br>Granrud 2014<br>Guillaume 2013<br>Ignell Modé 2014<br>Kim 2020<br>Leonard 2008<br>Liu 2019<br>Lorie 2021 | High confidence | "The Neonatology [unit] has a psychologist (...) but the nurses gave us a lot of support, every time we needed they gave us a lot of support." Amorim et al 2019 (Portugal, hospital)<br><br>"I am very satisfied with the time here . . . they make you feel special, that becoming a parent is something unique. Even though they see infants all the time, every day, the whole year, they succeed in making you feel quite special." Ignell Modé 2014 (Sweden, hospital)<br><br>"[The nurse] was cheering me up a bit because this time the [breastfeeding] didn't go as well as before. So, she was really supporting me, encouraging me not to give up" Liu et al 2019 (Canada, hospital) |

| | | | | |
|---|---|---|---|---|
| | | Mihae 2021<br>Ncube 2016<br>Orapiriyakul 2007<br>Petty 2018<br>Rossman 2011<br>Russell 2014<br>Sawyer 2013<br>Skene 2012<br>Veronez 2017<br>Yu 2020 | | "The nurse always welcomed me with a smile… I got positive energy from the nurse. Although, it was not always okay, the nurse kept saying positive words to me like 'your baby is holding up very well'." Mihae 2021 (South Korea, hospital)<br><br>"She then comforted me and told me to focus on the now and forget about the past. She said we should focus on the positive side and hope that the baby will be well. I felt better after talking to that nurse." Ncube 2016 (Botswana, hospital)<br><br>"I went into a bit of a depression… it was not postnatal depression as such, more the effect of everything… they were really supportive, got me some counselling, so that was good" Petty 2018 (UK, hospital and community)<br><br>"She counseled me, and she let me know 'if you want to stop, that's okay, there's no problem. You're the mom. You can do that.' But she was just letting me know that . . . 'you should just try to keep going for him at least while he's in the hospital,' and she just really talked to me about how good it is for him but also made me feel okay" Rossman et al 2011 (USA, hospital) |
| | **Support for fathers:** As above, but regarding emotional support specifically delivered to support the father of the infant. | 7 studies<br><br>Arnold 2013<br>Feeley 2013<br>Franck 2017<br>Leonard 2008<br>Lian 2020<br>Lundqvist 2019<br>Olsson 2017 | Moderate confidence | "I needed to be strong for her, because I knew that she was going through a hell of a lot. I was too … but … I think to balance all … the emotions … that I had to suppress it". Leonard 2008 (South Africa, hospital)<br><br>"Of course, I am very anxious, of course I am trying to think what can be done," Lian et al 2020 (Singapore, hospital) |
| | **Support from other parents in similar situation:** Carers wanted support from other parents of preterm or sick infants; to | 17 studies<br><br>Amorim 2019 | High confidence | "The other mothers know what we are feeling because they are feeling the same (...) and we are more comfortable to talk with them |

| | | | | |
|---|---|---|---|---|
| | develop their interactions and support the journey after a preterm or low birthweight birth. | Chang Lee 2009<br>Fernandez-Medina 2021<br>Franck 2017<br>Hägi-Pederson 2021a<br>Klawetter 2019<br>Leonard 2008<br>Lian 2020<br>Lomotey 2020<br>Ncube 2016<br>Neu 2020<br>Noren 2018<br>Petty 2018<br>Rossman 2011<br>Unsworth 2021<br>Villeneuve 2018<br>Yu 2020 | | [instead of a health professional].” Amorim et al 2019 (Portugal, hospital)<br><br>“Most people cannot understand our experience, only those who have watched their child in the NICU can understand what it’s like.. . they are the people who would understand fully.” Chang Lee et al 2019 (Taiwan, hospital)<br><br>“I went to a parents’ association meeting, and there was a little girl who is now nine years-old, an extremely preterm infant with 25 weeks of gestation and hydrocephalus. When I saw her I thought that my son had a chance, I was certain that my son’s health was going to get worse. This girl gave me faith and a strong sense of peace” Fernandez Medina et al 2021 (Spain, hospital and community)<br><br>“hope came from seeing and hearing other parents’ experiences.” Franck 2017 (UK, hospital and community)<br><br>“Often we would meet either one coming to the nursery and the other one leaving, in the bus sometimes, coming down in the taxi, we could enquire about each other’s children and that ... has also helped strengthen us as mothers”. Leonard 2008 (South Africa, hospital)<br><br>“It is not easy to position the baby for KMC but the nurses do not help us so I help one person to position her baby and she also helps with mine.” Lomotey et al 2020 (Ghana, hospital)<br><br>“Some mothers who were there are the ones that comforted me by saying, please touch him, kiss him. I started touching his legs and toes” Ncube 2016 (Botswana, hospital)<br><br>“One of the things that helped me was I joined a couple of NICU support groups (online) which kind of helped me know what to expect from other moms and help me understand some of the terms from a layman’s perspective. And then there are some moms in there |

| | | | | |
|---|---|---|---|---|
| | | | | whose other kids are 2 or 3 years old. So to kind of know that as far as milestones.” Neu 2020 (USA, hospital)<br><br>“The fact that they did walk in my shoes and they’ve been through some of the exact things that I was going through . . . made your experience of going through this a little easier to bear.” Rossman et al 2011 (USA, hospital)<br><br>“by the time I was leaving I had received a lot of support from the hospital and then there were also many mothers who had small babies, I was not alone. We were around five and we encouraged each other so that we could be strong for our babies that they may live and that whatever the people were saying could not happen.” Unsworth et al 2021 (Kenya, community) |

### Supplementary Table 14. Synthesis results with additional supporting data for analytical theme: “Healthcare environment”

| Analytical theme: Healthcare environment | Descriptive theme (review finding) | Studies contributing to review finding | CERQual grading | Supporting data |
|---|---|---|---|---|
| | **Access to babies:** Carers want mechanisms, or initiatives, to help them to visit and interact with their baby (including specific issues with the co-location of twins) | 15 studies<br><br>Abeasi 2020<br>Arnold 2013<br>Chang Lee 2009<br>Feeley 2013<br>Gallegos-Martinez 2013<br>Glazer 2021<br>Granrud 2014<br>Kim 2020<br>Leonard 2008<br>Lomotey 2020<br>Lundqvist 2019<br>Noren 2018<br>Orapiriyakul 2007<br>Russell 2014<br>Yu 2020 | High confidence | “You cannot just touch your baby when you want to, you have to be given the go ahead from the staff. It feels someway but I am helpless. You know you cannot go against the instructions of the staff. They are taking care of your baby so you have to respect them at all cost” Abeasi 2020 (Nigeria, hospital)<br><br>“they do not let me (enter the NU) [...] it doesn’t feel the same seeing her from a distance compared to up close [...] I wanted to hold her, but maybe it’s better like this, so she will get better soon, only her mom enters, maybe I don’t have to be there because she is more important for taking care of her because she is the mom [...] maybe it is because they put them in the incubators so they do not let us enter maybe because sometimes you can bring a cold or I don’t know; it complicates things” Gallegos-Martinez et al 2013 (Mexico, hospital)<br><br>“I was away from my baby immediately after I had the baby. It was too hard for me, as a mother. [...] Leaving her alone in the NICU, I felt too disheartened, and it was very, very hard.” Kim 2020 (South Korea, hospital)<br><br>“Every day, there are quarrels between some nurses and mothers. This happens when the babies are crying and their mothers want to attend to them but they are prevented by the nurses because the time is not due for them to enter the unit. Sometimes you can just feel your baby is crying because he/she is hungry or has soiled him/ herself but you will not be allowed to see the baby and those times are terrible.” Lomotey et al 2020 (Ghana, hospital)<br><br>“I saw him just a moment, how small he was. After delivering he was moved to the other room… Then I did not see him until next morning… I was anxious and worried about him.” Orapiriyakul et al 2007 (Thailand, hospital) |

| | | | | |
|---|---|---|---|---|
| | | | | "I had not seen him. He was born ten days ago. I had not seen him, it made me too sad" Yu et al 2020 (China, hospital) |
| | **Orientation and familiarity with NICU:** Carers want to learn about the healthcare setting (e.g. the noises, leads/equipment, and processes) in which they need to live and care for their infant (including the ability to tour the NICU before the birth if practical). | 19 studies<br><br>Abeasi 2020<br>Arnold 2013<br>Chang Lee 2009<br>Feeley 2013<br>Finlayson 2014<br>Glazer 2021<br>Guillaume 2013<br>Ignell Modé 2014<br>Klawetter 2019<br>Leonard 2008<br>Mihae 2021<br>Ncube 2016<br>Neu 2020<br>Noren 2018<br>Olsson 2017<br>Orapiriyakul 2007<br>Treherne 2017<br>Veronez 2017<br>Villeneuve 2018 | High confidence | "The whole set up appears stressful. We were asked to remove our slipper before we can enter the area were babies are kept, you need to wash your hands, generally there are a lot of rules" Abeasi 2020 (Nigeria, hospital)<br><br>"I'm glad they [showed me around], because it's quite daunting going into intensive care, NICU. I've never been in. All the, you know, computers, mechanical wombs basically for the premature babies. I'd never seen a premature baby previously. So it gave me an insight of what... it would freak me out if I'd just gone up there after having the babies. At least I knew where they were going." Arnold 2012 (UK, hospital)<br><br>"When I walked into this big room with all the incubators and all the other critically-ill little babies, I couldn't focus on just mine. There were so many machines sending out loud beeps. As I walked closer to the corner where they kept my baby, I nearly collapsed. He had so many lines and tubes attached to his tiny body...it was terrible...I just wanted to run away..." Chang Lee et al 2009 (Taiwan, hospital)<br><br>"You're sitting there and I feel for most of the time I'm looking at the monitors more than I'm looking at xxx [the baby]." Finlayson et al 2014 (UK, hospital)<br><br>"What was fantastic was that we could meet a physician and a nurse from here already at the delivery unit, before the infant was born. That information was nearly the most valuable of it all." Ignell Modé 2014 (Sweden, hospital)<br><br>"The nurse said, 'this is an oxygen giving tube'. 'This is a fluid tube to ensure that the baby gets enough nourishment'. 'This is an incubator with temperature and humidity control'. After receiving that |

| | | | | |
|---|---|---|---|---|
| | | | | explanation, I deeply understood my baby's situation.” Mihae 2021 (South Korea, hospital)<br><br>“And those machines sound the same for all the infants. As soon as an alarm goes off you want to check whether it’s our son who has a problem.” Olsson 2017 (Sweden, hospital)<br><br>“We could have been told that the people in the neonatal unit are different from those outside the unit. Do not be surprised by what you will see, because they do not breathe on their own, they are assisted to breathe.” Ncube 2016 (Botswana, hospital)<br><br>“My god … if the monitors could be wireless, it would change everything. The technology is there. It’s just somebody taking the time to do it!” Neu 2020 (USA, hospital)<br><br>“When I went to see him, he was in an incubator, with a device in the nose, CPAP, I was so scared, I didn’t know what was happening.” Veronez et al 2017 (Brazil, hospital) |
| | **Privacy vs monitoring:** Carers want privacy for them and their families for breastfeeding or everyday family activities (e.g. reading stories) in neonatal units. However, they also understand the need for medical observation and monitoring, and therefore struggle with the potential conflict between desire for privacy and desire for monitoring. This theme also included parental views on the structure and design of neonatal units (e.g. large wards versus individual rooms). | 6 studies<br><br>Feeley 2013<br>Klawetter 2019<br>Liu 2019<br>Neu 2020<br>Skene 2012<br>Treherne 2017 | Low confidence | “Even when we pumped milk, we didn’t need a curtain to hide behind, there aren’t many people, we are alone, and we are free to pump milk without embarrassment, without people seeing us” Liu et al 2019 (Canada, hospital)<br><br>“I like the open pod because I can see other mothers. It is easy to meet them and talk. The nurses can see the babies all the time.” Neu 2020 (USA, hospital)<br><br>“I saw people doing things and watched them. I don’t read but I watch so I’ve learned from watching, they have been very helpful.” Skene et al 2012 (UK, hospital)<br><br>“preoccupied with my surroundings [I] would lose the ability to focus on the time I was spending with my son.” Treherne et al 2017 (Canada, hospital) |

| | | | | |
|---|---|---|---|---|
| | **Staffing and equipment levels:** Carers are concerned about real, or perceived, issues with staffing levels, equipment availability, and seeing to the needs of the infant. | 10 studies<br><br>Amorim 2019<br>Arnold 2013<br>Chang Lee 2009<br>Franck 2017<br>Granrud 2014<br>Kim 2020<br>Liu 2019<br>Ncube 2016<br>Russell 2014<br>Unsworth 2021 | High confidence | "I often feel there is a shortage of medical staff working in the NICU. It may be or must be too hard for one nurse to care for several babies properly at once." Kim 2020 (South Korea, hospital)<br><br>"The nurse was [. . .] rushing. [. . .] It's not [an] ideal situation where nurses don't have enough time to take care of each [baby]" Liu et al 2019 (Canada, hospital)<br><br>"What made me sad is ... you will find your baby there ... not taken care of ... If you happen not to go to the unit because you were not feeling well, by the time you go there to check on her, you will find her in the same sheets and the nappy not changed." Ncube 2016 (Botswana, hospital)<br><br>"It was reassuring as well, because it was almost one-on-one care. So it was like she was being monitored the whole time. If she needed anything, there was somebody there straight away. Erm, so you felt that you could leave her, and there was nothing we could do, it was just the medical staff and they needed to do what they needed to do." Russell 2014 (UK, hospital)<br><br>"Long waiting time because if the mothers are so many and could be few staff" Unsworth et al 2021 (Kenya, community) |

### Supplementary Table 15. Synthesis results with additional supporting data for analytical theme: “Information needs met”

| | Descriptive theme (review finding) | Studies contributing to review finding | CERQual grading | Supporting data |
|---|---|---|---|---|
| **Analytical theme: Information needs met** | **Information about the baby:** Carers want information relating to the baby’s condition, prognosis, investigations/procedures performed or daily events. However, they also can feel overloaded and there is often a conflict between wanting to be told about all possible information and outcomes, and only wanting to know what they need to know at that particular point in time. | 17 studies<br><br>Amorim 2019<br>Brødsgaard 2015<br>Dorner 2020<br>Franck 2017<br>Gallegos 2013<br>Granrud 2014<br>Guillaume 2013<br>Hendriks 2017<br>Ignell Modé 2014<br>Lomotey 2020<br>Mihae 2021<br>Ncube 2016<br>Orapiriyakul 2007<br>Russell 2014<br>Veronez 2017<br>Villeneuve 2018<br>Yu 2020 | Moderate confidence | ‘‘Mother: One day, I arrived at NICU and I saw the incubator empty... I was in shock. I didn’t ask any question, I just started crying. When a nurse saw me [crying], she ran to tell me that my daughter was moved to be closer to her twin. She should have been more careful and call me (...) or to the father. This could seem the most insignificant thing in the world but it’s not, it’s very important [for us].” Amorim et al 2019 (Portugal, hospital)<br><br>“I’ve definitely been taking it day by day. I’m choosing not to stress myself out about something that I know nothing about, which is tomorrow; I have no idea what it’ll hold” Dorner et al 2020 (USA, hospital)<br><br>“I don't care what the doctor says about other children, I just care about my own child. I wanted to know what the doctor had to say. I wasn't allowed to and that was a pity.” Granrud et al 2014 (Norway, hospital)<br><br>“Sometimes when you are kangarooing and they pass during rounds . . . it is difficult to have a discussion when 80% of your brain is focusing on the infant, and you just try to engage in conversation with 20%.” Ignell Modé 2014 (Sweden, hospital)<br><br>“The additional things that the nurses explained to me helped a lot. What did this test do, and maybe the baby would react to it this way… Talking about the process helped me understand the baby's treatment.” Mihae 2021 (South Korea, hospital)<br><br>“They did the chest X-ray and the head X-ray… Then they told me that the X-ray revealed bleeding from the lungs. The way they told me the disadvantages of bleeding from the lungs … I ended up accepting the situation ….” Ncube 2016 (Botswana, hospital) |

| | | | | |
|---|---|---|---|---|
| | | | | “She explained every machine to me: ‘That’s to monitor her heart rate’, she goes ‘Your baby’s not on oxygen, she’s on air just to help her lungs, 'cos you do know baby’s really small. Baby’s not sick mummy.’ And she explained everything to me. The machines, how they incubate her.” Russell 2014 (UK, hospital)<br><br>“Another thing was that some doctors tended to [exaggerate]…the baby's disease and it really scared us. They would…inform us about all the possible complications. I felt unsure about those issues. I did not know how to describe the feelings, especially when they [complications] did not happen to my baby”. Yu et al 2020. (China, hospital) |
| | **Frequent updates:** Carers want frequent and regular updates from the clinical team, rather than just meetings when sentinel events occur, alongside open channels of communication. | 7 studies<br><br>Blomqvist 2012<br>Ignell Modé 2014<br>Leonard 2008<br>Neu 2020<br>Russell 2014<br>Veronez 2017<br>Villeneuve 2018 | Moderate confidence | “They do inform you every day ... they keep you up to date. ... inform you of the progress of the infant, if there is something wrong with him or things like that, the sisters are quite clued up so they keep you clued up as well”. Leonard 2008 (South Africa, hospital)<br><br>“They ask you if you want to be in for the rounds every time and I’m like, ‘of course I want to be in for the rounds.’ They’ll just open up the door and let you sit in.” Neu 2020 (USA, hospital) |
| | **How information is given:** Carers want HCPs to have good communication skills and to use a variety of information-giving methods. This relates to a need for immediacy, an appropriate pace and timing of information giving, and the opportunity for follow-up discussions. However, what constitutes good communication may vary by parent, culture and situation. | 9 studies<br><br>Abeasi 2020<br>Amorim 2019<br>Franck 2017<br>Guillaume 2013<br>Hendriks 2017<br>Lorie 2021<br>Veronez 2017<br>Villeneuve 2018<br>Yu 2020 | High confidence | “I try to read online, but most often because I am not a medical person, I do not understand and would have wished to get more information from the staff, especially the nurses. I believe the staff can explain things to me at my level by excluding all those big words” Abeasi 2020 (Nigeria, hospital)<br><br>"It's very good to have news by telephone... it takes 15 seconds but afterwards, you feel so much better... then pffff! I pump my milk and I fill the bottle" Guillaume et al 2013 (France, hospital)<br><br>“I mean, I knew it was busy but they still do not have the right to drop heavy news just like that and then say ‘oh, by the way, I don’t have any time right now to talk about it but we will do so |

| | | | | |
|---|---|---|---|---|
| | | | | tomorrow'. Well, you just cannot do that and, for me, that was really like . . . I don't want to talk to you anymore." Lorie et al 2021 Netherlands, hospital)<br><br>"[. . .] I think they were pussyfooting" Villeneuve et al 2018 (UK, hospital) |
| | **Matching needs with information.** Carers want their informational needs and expectations to be met, and to feel confident in obtaining that information (e.g. by being comfortable or enabled to ask question). They also want to supplement this information via other sources (e.g. leaflets, books and the internet). | 18 studies<br><br>Abeasi 2020<br>Brødsgaard 2015<br>Chang Lee 2009<br>Finlayson 2014<br>Frack 2017<br>Glazer 2021<br>Guillaume 2013<br>Hendriks 2017<br>Ignell Modé 2014<br>Kim 2020<br>Klawetter 2019<br>Lian 2020<br>Liu 2019<br>Lomotey 2020<br>Lorie 2021<br>Russell 2014<br>Villeneuve 2018<br>Yu 2020 | High confidence | "The nurses and doctors are seen going around and it's difficult to get a lot of information regarding what is really happening to your baby. You know you cannot blame them, they are busy but there should be a way around it… I sometimes wish I have more information, something like an update every now and then." Abeasi 2020 (Nigeria, hospital)<br><br>"And they said, 'you don't need to concern yourself with that, we need to concern ourselves with that'. And I was like, actually I don't agree, I think I need to know that as well cos I'm his mum." Finlayson et al 2014 (UK, hospital)<br><br>"no question was a stupid question." Franck 2017 (UK, hospital)<br><br>"They always told me everything that was going on, whether it was a bath, whether it was a line change. They always gave me all the information I needed, I never felt worried when went home. I always felt like my baby's safe, everything's fine." Glazer et al 2021 (USA, hospital)<br><br>"If there is no problem with the examinations, the doctors don't come to tell you the results. (..) If they tell us the results right away, whether they are good or bad, we know them and we can start to enjoy the child" Guillaume et al 2013 (France, hospital)<br><br>"One would inform us well and would start telling us everything on her own; with others you had to drag the information out of them." Hendriks 2017 (Switzerland, hospital) |

| | | | | |
|---|---|---|---|---|
| | | | | "I think it has been very clear . . . and good. There is a lot of information all the time, but not so much that I need to ask more questions afterwards. Most of what I am wondering about is covered." Ignell Modé et al 2014 (Sweden, hospital)<br><br>"Sometimes I feel like maybe nurses and doctors are so used to everything that's happening with most of these babies because they see it a lot, but to parents it's all new and it's still really scary. So, when they have to call in people to help them because the heart rate dropped or something, they don't think that they should let the parents know until they're coming in. To me, that's something that I want to know when that happens. I don't want to wait. I definitely think that they could communicate certain things a little bit more and involve us in it …" Klawetter 2019 (USA, hospital)<br><br>"[in relation to internet forums] I see the response from people and the amount of support coming right, I become more relaxed and I feel like… people can understand". Lian et al 2020 (Singapore, hospital)<br><br>"I had an answer and not only an answer, but I had an explanation and [the nurse] really ensured that I understood" Liu 2019 (Canada, hospital)<br><br>"They (health workers) usually inform us before performing procedures on our babies but they do not always explain the purpose to us." Lomotey 2020 (Ghana, hospital)<br><br>"the other doctors had decided, that it was too soon and they needed to wait a bit longer, but nobody had told us that, so we're expecting results, and we're not getting anything we haven't even had the test". Russell 2014 (UK, hospital) |

**Supplementary Table 16. Synthesis results with additional supporting data for analytical theme: "Logistical support"**

| Analytical theme: Logistical support | Descriptive theme (review finding) | Studies contributing to review finding | CERQual grading | Supporting data |
|---|---|---|---|---|
| | **Accommodation (comfort and facilities).** Carers need practical support in travelling to see their infants, and support to sleep and live near them during their initial inpatient stay. | 13 studies<br><br>Amorim 2019<br>Blomqvist 2012<br>Franck 2017<br>Glazer 2021<br>Granrud 2014<br>Klawetter 2019<br>Leonard 2008<br>Liu 2019<br>Lomotey 2020<br>Neu 2020<br>Noren 2018<br>Russell 2014<br>Villeneuve 2018 | High confidence | ''Mother: I think it's missing a room for parents. Father: Yes for, those who are there [NICU] all day, resting. The available room had only one chair, (...) without any furniture, only lockers. We need a coffee machine, a water machine (...) some chairs to talk to each other and get some rest." Amorim et al 2019 (Portugal, hospital)<br><br>"Those beds were anything but what you would call a bed, perhaps you know what they look like. They are like sofa beds they are hideous. I don't think I have ever had such a pain in my back after sleeping in a bed before, but, all you could do was to endure." Blomqvist et al 2012 (Sweden, hospital)<br><br>"I've found even if you have money to get up here, the food is really expensive and you only have $5 left. The [bus] fare is $5 minimum, so it's like should I eat or have money to get home? So, you're like okay I need to get home, so you don't end up eating and you're starving." Klawetter 2019 (USA, hospital)<br><br>"It would be cool to have two sofa beds. Like . . . [dad] is not obliged . . . he can stay with the mom, and then he does not have to stay in the same sofa." Liu 2019 (Canada, hospital)<br><br>"All the mothers cannot fit in the mothers' room here and even the beds are not enough so some of us sleep on the corridor." Lomotey et al 2020 (Ghana, hospital)<br><br>"It would be nice to have a dedicated space for NICU families. Since you're spending so much time here, it's not like you're just here for a couple of days. People are living here, spending weeks, if not months. Having something just for NICU families where you could get food sent to you if you wanted to buy it or whatnot, I think |

| | | | | |
|---|---|---|---|---|
| | | | | would be a lot. Again, just all those things that are extra to stress about, and worry about, and deal with." Neu 2020 (USA, hospital)<br><br>"[The neonatal staff] appreciate that there's a bond between parents and baby that needs to be maintained. The biggest plus was… the (neonatal) unit got us a room over the road". Russell 2014 (UK, hospital)<br><br>"Playrooms for other children, so that they could come to the unit too and not be left out of the picture." Villeneuve et al 2018 (UK, hospital) |
| | **Broader family support and impact:** Carers want support for the wider family (such as creche facilities for siblings) to allow the carers to engage in their 'normal' parenting and caring roles, before, and after, discharge | 13 studies<br><br>Feeley 2013<br>Gallegos-Martinez 2013<br>Hägi-Pederson 2021a<br>Klawetter 2019<br>Leonard 2008<br>Lundqvist 2019<br>Ncube 2016<br>Neu 2020<br>Nyondo-Mipando 2020<br>Olsson 2017<br>Unsworth 2021<br>Veronez 2017<br>Villeneuve 2018 | High confidence | "Trying to manage everything together has an impact. Work, come home, do laundry, clean the house, take care of my son, go to the internet, work, wake up. It's just a continuous cycle" (father) Feeley et al 2013 (Canada, hospital)<br><br>"(To come) [...] (I am supposed to) leave my children in the care of others, and right now my (other) girl is sick and (I am supposed) to risk that to come here" Gallegos-Martinez et al 2013 (Mexico, hospital)<br><br>"There were days when I couldn't get up here and it made me feel terrible. Like I was missing him [infant]. And then I felt like, 'Dang, I'm not able to see him every day.' I was feeling down on myself. I have these other children, so I didn't want to spend too much time here and neglect them and their needs. It was hard to juggle, me with four kids. So that's why I'm really excited about coming home. It will make it a lot easier, I won't have to come up here. I'm a single mom, so it's hard. It's really hard." Klawetter 2019 (USA, hospital)<br><br>"Mostly I was focused on my studies since I was still learning . . . I left him at home then I went back to school, when I went back to school, was given my mother's sister, she also then went back to school, so I left the baby to my neighbour". Unsworth et al 2021 (Kenya, community) |

| | | | | |
|---|---|---|---|---|
| | | | | "Every day I have more hope that he'll leave soon. His little sister is at home, she's anxious to meet the brother who was born, but has still not come home" Veronez et al 2017 (Brazil, hospital)<br><br>"[. . .] every day we ended up saying sorry [to each other] and starting each day [. . .]" Villeneuve et al 2018 (UK, hospital) |
| | **Costs of treatment:** Cares want additional support, or recognition, of the direct and indirect costs (e.g. travel) needed while the infant is an inpatient, and then afterwards for additional appointments and lost earnings because of the preterm birth. | 12 studies<br><br>Abeasi 2020<br>Amorim 2019<br>Fernandez Medina 2021<br>Franck 2017<br>Gallegos-Martinez 2013<br>Klawetter 2019<br>Lomotey 2020<br>Mihae 2021<br>Nyondo-Mipando 2020<br>Petty 2018<br>Unsworth 2021<br>Villeneuve 2018 | High confidence | "The doctors said my child has another condition, so I buy a lot of drugs which are very expensive. I have seen people lose their babies after all the money spent, I just pray mine gets well" Abeasi 2020 (Nigeria, hospital)<br><br>"I had to leave my job to take care of him, I couldn't miss work an average of twice a week for medical appointments. We had to tighten our belts a lot, if you don't have savings or family who can help you financially it is extremely complicated. We had to move to my parents' house because we had no money" Fernandez Medina 2021 (Spain, hospital and community)<br><br>"The NHIS [National Health Insurance Scheme] is supposed to be functioning but we buy some of the drugs…., they are quite expensive." Lomotey et al 2020 (Ghana, hospital) |
| | **Parental leave:** Parents want formal, protected leave to allow them to visit the baby whilst in hospital, and to support them to provide care for a prolonged period after birth. | 7 studies<br><br>Amorim 2019<br>Fernandez Medina 2021<br>Klawetter 2019<br>Lundqvist 2019<br>Nyondo-Mipando 2020<br>Olsson 2017<br>Villeneuve 2018 | Moderate confidence | ''Mother: I should have the right to have a bigger parental leave [100% instead of 65% of the salary], at least during the hospitalisation period. Mother stays there [NICU] for a lot of hours alone in a very difficult situation. At least during the hospitalisation in NICU, the father and the mother should have the right to stay both with a [full parental] leave (...)." Amorim et al 2019 (Portugal, hospital)<br><br>"The other frustrating thing for me is that I have been supposed to be going back to work. I had 8 weeks medical leave. I probably can extend it, but I have only been at this job for 3 months before I ended up in the hospital … So [I'm] trying to look into options so |

| | | | | |
|---|---|---|---|---|
| | | | | that I can start plugging back in to work. Remotely earlier, and then I can extend my leave so that I at least get a couple of weeks at home with him [infant] after I get discharged to try to settle into a routine." Klawetter 2019 (USA, hospital)<br><br>"They gave me only seven days as a holiday and I am worried they may terminate my contract should I stay here longer than expected." Nyondo-Mipando 2020 (Malawi, hospital) |

**Supplementary Table 17. Synthesis results with additional supporting data for analytical theme: "Positive relationships with staff"**

| | Descriptive theme (review finding) | Studies contributing to review finding | CERQual grading | Supporting data |
|---|---|---|---|---|
| **Analytical theme: Positive relationships with staff** | **Compassion and sensitivity.** Carers desire to develop a caring and sensitive relationship with HCPs caring for their baby. | 19 studies<br><br>Amorim 2019<br>Franck 2017<br>Glazer 2021<br>Guillaume 2013<br>Hendriks 2017<br>Kim 2020<br>Lian 2020<br>Liu 2019<br>Lomotey 2020<br>Lorie 2021<br>Lundqvist 2019<br>Mihae 2021<br>Nyondo 2020<br>Petty 2019b<br>Premji 2017<br>Russell 2014<br>Veronez 2017<br>Villeneuve 2018<br>Wernet 2015 | High confidence | "There are extraordinary nurses, they talk all the time to the baby, 'my sweetheart, I'm going to do this for you.' I love it that she is in such good hands" Guillaume et al 2013 (France, hospital)<br><br>"One of the nurses even wrote me an e-mail in the middle of the night to tell me that my baby was sleeping well. That touched me so because that was my first night at home. I found that amazing. What a calling, such a job." Hendriks 2017 (Switzerland, hospital)<br><br>"I was too awkward at feeding and burping my baby and even changing her diaper because it was the first baby in my life. The nurses taught me in a kind and gentle way; so I could rely on them." Kim 2020 (South Korea, hospital)<br><br>"You know they are dedicated professional individuals that care deeply about what they are doing and very compassionate….(that) allows you to focus on yourself, your daughter, and your wife." Lian 2020 (Singapore, hospital)<br><br>"We had one nurse, I recall, she was an incredibly sweet woman and she was also constantly concerned with how we were doing, and she also just said 'I understand that you are having a hard time', you know, and that really stayed with us" Lorie et al 2021 (Netherlands, hospital)<br><br>" 'Grandma, we want your grandson to survive.' So from that statement, I knew that every initiative that would be put in place would be for the survival of my grandson… You know when doctors or these nurses come to your bed you feel like your grandson is also considered important by them and gives a sense of hope. They send a message to us that they are hoping for life on our patient." Nyondo-Mipando et al 2020 (Malawi, hospital) |

| | | | | |
|---|---|---|---|---|
| | | | | "The small things make such a difference." Petty 2019b (UK, hospital and community)<br><br>"[The staff were] incredibly empathic and you know even, they were giving us the, the prognosis with Z, and they had tears in their eyes you know the way that they were saying it and you kind of, it makes it feel like you're a person who is experiencing a terrible thing rather than just another number going through the process." Russell 2014 (UK, hospital) |
| | **Consistency in care/communication:** Carers want consistent rules, advice, therapeutic plans and predictions of outcome, from, and between, different HCP. | 13 studies<br><br>Blomqvist 2012<br>Feeley 2013<br>Finlayson 2014<br>Franck 2017<br>Glazer 2021<br>Granrud 2014<br>Hendriks 2017<br>Ignell Modé 2014<br>Kim 2020<br>Liu 2019<br>Neu 2020<br>Premji 2017<br>Russell 2014 | Moderate confidence | "Some said 'you should touch him.' Some said 'you shouldn't touch him'." Feeley et al 2013 (Canada, hospital)<br><br>"To me it's just about consistency and every nurse does things differently. One nurse will tell you to do one thing and the next nurse will come in and criticise you 'cos they wind or feed a baby differently and it makes you feel like crap" Finlayson et al 2014 (UK, hospital)<br><br>"In the course of 16 days, I counted that I'd spoken to 16 different nurses, midwives and children's nurses about breastfeeding. And when I talked to the sixteenth person on the sixteenth day who gave me the sixteenth piece of advice about how to get started – well, I just lost it completely in the lactation room." Granrud et al 2014 (Norway, hospital)<br><br>"During every shift, there was someone who has participated since the first day. It is important in the beginning that one senses a connection between those who substitute each other . . . but I have felt a certain continuity on the whole.. . ." Ignell Modé et al 2014 (Sweden, hospital)<br><br>"in contact with five different public health nurses and heard five different sets of advice that were really contradictory; I had a really hard time trusting what I was being told." Premji et al 2017 (Canada, community) |

| | | | | |
|---|---|---|---|---|
| | **HCP expertise/care:** Carers want well-trained, competent, staff able to provide the specialist care needed by the infant. | 8 studies<br><br>Finlayson 2014<br>Kim 2020<br>Lomotey 2020<br>Premji 2017<br>Russell 2014/Sawyer 2013<br>Unsworth 2021<br>Veronez 2017<br>Villeneuve 2018 | High confidence | “It worries me and my partner this last week as we’ve noticed the different care you can get with nurses and the difference in attentiveness . . . we sometimes worry who’s looking after him.” Finlayson et al 2014 (UK, hospital)<br><br>“All the doctors that were there, as far as I’m concerned, were the experts. The doctors had, you know, been in this industry for like 15, 20, 25 years. They knew, they knew their stuff inside out you know, the information was never flaky…” Russell 2014 (UK, hospital)<br><br>“Absolute confidence in the staff. I didn’t feel like I needed to know every step of the way. I was able to just step back, realise that control was not mine. The control was where it should be, with professionals, and they would take good care of them [the babies]” Sawyer 2013 (UK, hospital)<br><br>‘I was attended to by trainees, so I did not know who was doing the right thing and (and who was doing) the wrong (thing). They (baby) were not handled in the right way.’ Unsworth 2021 (Kenya, community) |
| | **Respect, collaboration and trust:** Carers desire the development of collaborative relationships with the HCP; where the parent is identified as an important part of the care-team with unique skills and knowledge of the baby, and is heard and understood. | 14 studies<br><br>Amorim 2019<br>Finlayson 2014<br>Franck 2017<br>Glazer 2021<br>Klawetter 2019<br>Liu 2019<br>Lorie 2021<br>Ncube 2016<br>Petty 2019b<br>Russell 2014/Sawyer 2013<br>Skene 2012 | Moderate confidence | “Yes, they [doctors] said to us that he had two little points in the ultrasound (...) [and then] in the morning they said to me that it was a little haemorrhage, without importance (.. .) after that we couldn’t trust [the doctors] because they didn’t tell us the truth (...)” Amorim et al 2019 (Portugal, hospital)<br><br>“She [the nurse] was like, ‘Yes, I already started [the diaper change]. You know, I have a lot of things to do. I have a lot of babies.’…So, she can see my face that I’m really, really upset now. She’s like, ‘What’s wrong?’ I said, ‘Nothing.’ – my husband tells me, like in Creole, ‘I don’t know why you’re wasting your time, going back and forth with her.’ So, she goes, ‘Oh, don’t speak your language here.’ So, that was a whole ‘nother story. So, now I had to |

| | | | | |
|---|---|---|---|---|
| | | Treherne 2017<br>Villeneuve 2018<br>Wernet 2015 | | go and report her, because you can’t tell me, don’t speak my language here.” Glazer et al 2021 (USA, hospital)<br><br>“She [healthcare professional] just did not understand us, and she did not listen, and she was actually just working against us. So, we requested to have her near us as little as possible” Lorie et al 2021 (Netherlands, hospital)<br><br>“We could review the information at any moment we wanted without feeling like ‘helicopter parents’. That made me feel that the care team trusted us and it allowed me to trust them.” Treherne et al 2017 (Canada, hospital)<br><br>“Doctor [name removed] was very good and talked to my husband . . . until his questions finished and he had resolutions or some kind of answers . . . the fact that he had been heard was really, really important. You know, you can’t always give an answer or solve the problem but at least he’d been heard.” Villeneuve et al 2018 (UK, hospital)<br><br>“They [health professionals] don’t understand that. We are afraid they’ll die; we just want it to be all right. I’m the mother. (She thinks) They don’t see that; they just criticize us” Wernet 2015 (Brazil, hospital) |
